# Money, jobs or schooling? A model-based evaluation of economic strengthening in South Africa and its impact on HIV, sexually transmitted infections and teenage births

**DOI:** 10.1101/2025.05.25.25328300

**Authors:** Leigh F. Johnson, Lise Jamieson, Mmamapudi Kubjane, Gesine Meyer-Rath

## Abstract

**Background:** High incidence rates of HIV, sexually transmitted infections (STIs) and teenage pregnancy are major challenges facing South Africa. The role of socio-economic factors in driving these is complex, with high socio-economic status protecting against some risk behaviours (condomless sex, early sexual debut and casual/transactional sex in females) but increasing other risk behaviours (e.g. male engagement in casual and commercial sex). Consequently, the impact of different economic strengthening interventions is unclear.

**Methods:** We extended a previously-developed agent-based model of HIV, STIs and fertility in South Africa, to reflect effects of education, employment and per-capita household income on sexual behaviours. These effects were estimated from literature and from calibration of the model to African randomized controlled trials of economic strengthening interventions.

Population attributable fractions (PAFs) were calculated. We considered three intervention types, all targeting households with log per-capita income below the national average: school support to reduce school dropout; vocational training for unemployed adults; and unconditional cash transfers.

**Results:** Low socio-economic status is estimated to have accounted for 14% of new HIV infections, 10% of incident STIs (gonorrhoea, chlamydia and trichomoniasis) and 46% of teenage births in South Africa, over 2000-2020. However, because of uncertainties regarding effect sizes, confidence intervals around these PAFs are wide (3-32%, 0-21% and 12-89% respectively), with uncertainty in the effect of education on condom use being the most significant correlate of the HIV PAF (r=0.94) and the relative rate of female sexual debut while in school being the most significant correlate of the teenage birth PAF (r=-0.96). Over 2025-2040, none of the interventions are estimated to have impacts significantly different from zero, due to limited impact on secondary economic outcomes. However, school support would come close to significantly reducing teenage births (by 4%, 95% CI: -1-18%).

**Conclusions:** Although poverty is likely to be a significant driver of HIV, STIs and teenage pregnancy in South Africa, precise quantification is challenging. Recently trialled economic strengthening interventions have insufficient impact on socioeconomic status to reduce HIV and STIs significantly at a population level.

## Background

Countries in southern and eastern Africa have the highest levels of HIV prevalence globally [1], as well as high incidence of curable sexually transmitted infections (STIs) [2] and teenage fertility [3]. High levels of poverty in the region are often blamed for these reproductive health challenges [4, 5]. Teenage pregnancy in Africa is indeed associated with poverty, low female employment and incomplete schooling [3, 6–9]. However, the relationship between socio-economic status and the risk of HIV and STIs in Africa is more complex. For example, early reviews documented a *positive* relationship between socioeconomic status and HIV in Africa [10–12], but later reviews have found a more nuanced picture, with suggestions of a change in the relationship between HIV and socioeconomic status over time [13–15] and variation in the relationship across regions within Africa [15–20]. Similarly, STIs are often associated with employment [21, 22] and wealth [22], but have also been found to be associated with low educational attainment [22, 23].

The complexity of the relationship between socio-economic status and HIV/STI risk reflects a diversity of ways in which education, employment and wealth can both mitigate and potentiate sexual risk behaviour. Higher educational attainment is strongly associated with greater condom use in non-spousal relationships [11, 24–27], and schooling and household income both delay the timing of female sexual debut and entry into marriage [11, 26, 28–32]. In men, employment may be associated with engaging in commercial and transactional sex [33–35] as well as multiple partnerships [30, 36]. In women, on the other hand, lower income is associated with transactional sex [37–39]. Factors other than sexual behaviour may also affect the observed association between HIV/STIs and socioeconomic status. For example, voluntary medical male circumcision (VMMC), which protects against male HIV and STI acquisition, is more common in men with greater wealth [40, 41]. A higher HIV prevalence in higher socio-economic strata might be a reflection of longer survival [42], due to higher rates of HIV testing [43] and better access to treatment in wealthier individuals. The relationship between wealth and HIV might also be partly confounded by urbanicity, with urban areas having both higher levels of wealth and higher HIV prevalence [14, 17].

Economic strengthening interventions have been developed in an attempt to address some of the socioeconomic drivers of HIV, STIs and teenage pregnancy. The most commonly tested interventions include cash transfers (either unconditional or conditional upon achieving certain educational/health outcomes), educational support, vocational training, microcredit schemes and food assistance, among others [44]. Although reviews of the impacts of these interventions have noted some reductions in self-reported risk behaviours and improved contraceptive knowledge, there is relatively little evidence of changes in clinical outcomes (HIV and STI incidence and unintended pregnancy) [44–48]. Very few studies evaluated whether intervention impacts were sustained over the longer term [45] or whether the interventions themselves were sustainable [48]. There is a lack of consensus on which interventions are most appropriate and in what contexts.

Mathematical models are widely used to inform policy decisions around HIV and reproductive health [49–51], and play a particularly important role in evaluating the relative cost-effectiveness of different interventions and the optimal allocation of scarce healthcare resources. Yet there is a notable lack of mathematical modelling of economic strengthening interventions [52]. This is partly a reflection of the challenges associated with representing the complex causal pathways described previously, a challenge common to the modelling of structural interventions generally [53]. It is also partly due to the lack of consistent evidence of the impact of economic strengthening interventions on clinical outcomes. In this study we make a first attempt at modelling the impact of these interventions in South Africa, the country with the world’s largest HIV epidemic, and the highest level of income inequality globally [54].

## Methods

### Model structure

We adapted a previously-developed model of HIV and reproductive health in South Africa (MicroCOSM, or Microsimulation for the Control of South African Morbidity and Mortality). This is an agent-based model, which has previously been used to evaluate structural drivers of HIV and STIs [55–57]. Briefly, the model simulates a nationally-representative sample of the South African population, with each individual (‘agent’) being randomly assigned a set of characteristics: demographic (age, sex, race), socioeconomic (education, urban/rural location, migration and incarceration history), psychological (gender norms, conscientiousness), healthcare access (use of hormonal contraception, HIV prevention and HIV testing/treatment), and behavioural (alcohol consumption, propensity for concurrent partners, sexual experience, sexual preference, current relationship/marital status). Four types of sexual relationship are modelled: marital/cohabiting, short-term non-cohabiting, casual/once-off (characterized as ‘transactional sex’) and commercial sex (between sex workers and clients). When new partnerships are formed, individuals are linked to other individuals in the simulated population, and the transmission of HIV/STIs is modelled based on specified probabilities of transmission per condomless sex act (with multiplier adjustments to account for factors such as the HIV-positive partner’s HIV viral load and male circumcision). A woman’s monthly probability of conception is similarly based on the woman’s numbers of sexual partners, age and contraceptive use. The simulation begins in 1985, with HIV being introduced into the simulated population in 1990.

For the purpose of the current study, the model was extended to identify family links between individuals (parents, children and siblings), and based on family and marital relationships individuals are grouped into households. Household formation and dissolution is modelled dynamically, with the model being calibrated to national survey data on household composition (see section 1.2 of the supplementary materials).

The model was further extended to simulate employment in individuals aged 15-64 who are not currently in school/studying or incarcerated. Rates of entering employment are assumed to depend on age, sex, educational attainment, race, urban/rural location and parity (in women). In addition, people living with HIV are assumed to experience a reduced odds of employment if they are untreated and have low CD4 counts [58–60]. The model is calibrated to national survey data on employment levels by age, sex and race (see section 1.1 of the supplementary materials).

Four sources of household income are modelled: salaries/wages (for each employed member in the household), child support grant payments, the state old age pension and private pensions. A more detailed description of these four income sources is provided in section 1.3 of the supplementary materials. The adjusted per capita household income (APCHI) is calculated as

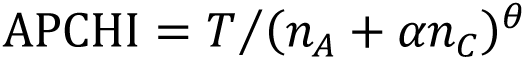

where *T* is the total household income, *n*_*A*_ and *n*_*C*_ are the numbers of adults (15+) and children respectively in the household, *α* is the relative cost of supporting children (relative to adults) and *θ* represents the economies of scale in meeting the needs of larger households. In line with previous South African studies [61], we set *α* = 0.5 and *θ* = 0.9.

Figure 1 illustrates the hypothesized causal pathways linking socio-economic status and HIV/reproductive health outcomes. Economic strengthening interventions are assumed to affect socio-economic variables (educational attainment, employment and household income), and these are in turn assumed to affect sexual behaviour; sexual behaviour then influences HIV, STIs and pregnancy (which can in turn influence socio-economic status, e.g. pregnancy causing school dropout). Table 1 summarizes the key parameters that relate socio-economic status to sexual risk behaviour and health seeking behaviour. Three intervention types are modelled, all targeting households with adjusted per capita income below the national average: (1) school support to reduce school dropout (the intervention is also assumed to increase the chance of re-enrolment in youth aged 13-22 who have dropped out of school); (2) vocational training (for ages 20-49); and (3) unconditional cash transfers. These interventions were selected from a more complete list of economic strengthening interventions [44], based on the availability of data regarding their impacts on HIV/STI/teenage pregnancy outcomes. The school support intervention is assumed to include both material support (e.g. school uniforms, cost of school transport) and non-material support (e.g. attendance monitoring and counselling). The effects of the two components are modelled separately, with half of the cash equivalent value of the material support being added to the household income, in the same way as for cash transfers.

**Figure 1:**
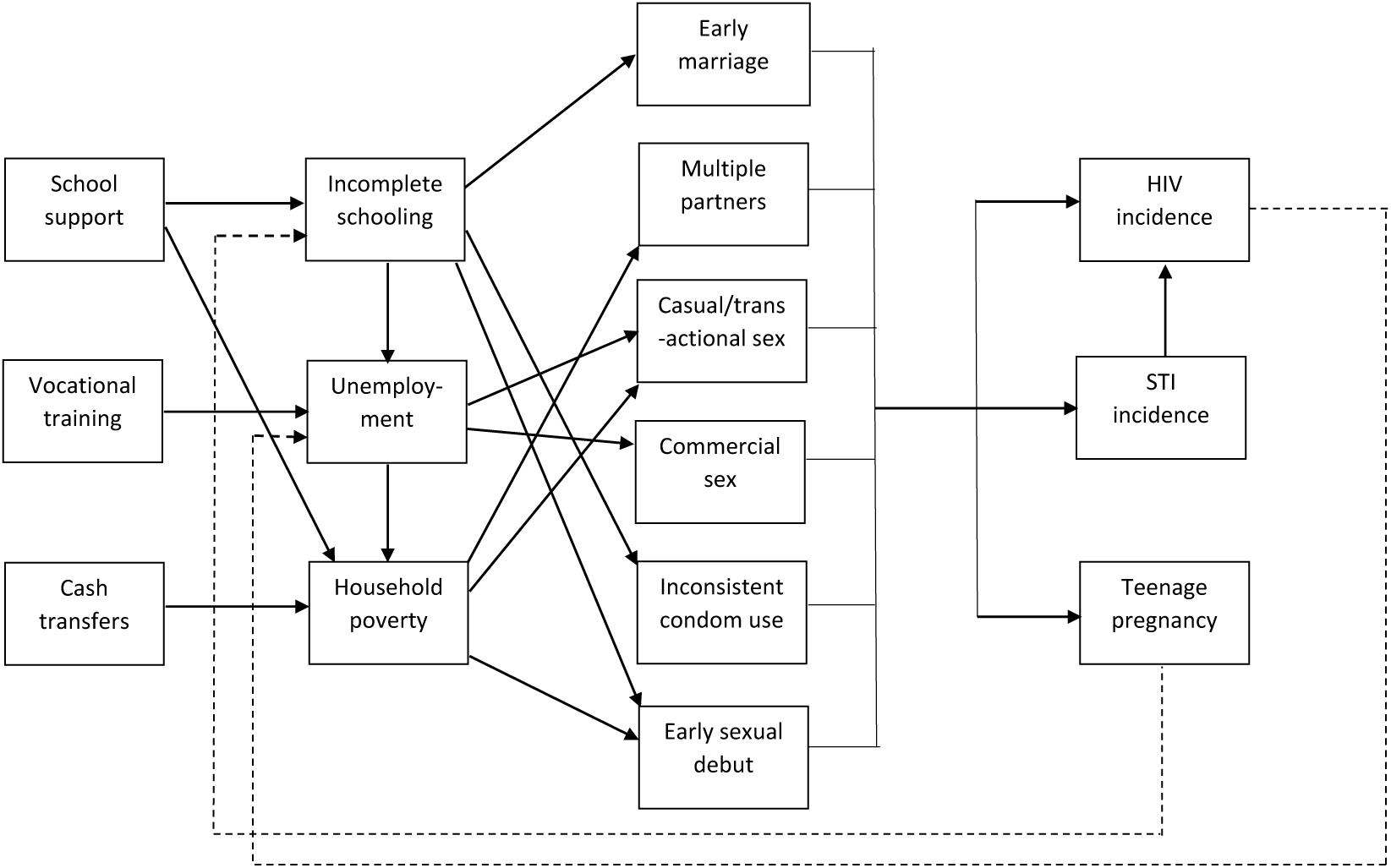
Model of economic determinants of sexual risk behaviour.

**Table 1:** Model parameters relating socio-economic status to sexual risk behaviour and health seeking behaviour.

| Model parameters | Mean | SD | Data sources |
| --- | --- | --- | --- |
| Sexual debut |  |  |  |
| Increase in rate of debut in females per log reduction in APCHI* | 0.23 | 0.28 | [30-32, 62-65] |
| RR of sexual debut in females if currently in school | 0.46 | 0.44 | [64, 66-72] |
| RR of sexual debut in males if currently in school | 0.80 | 0.45 | [66, 68] |
| School dropout |  |  |  |
| Probability of permanent dropout in the year of giving birth | 0.35 | - | [73] |
| Casual/transactional sex |  |  |  |
| Increase in casual sex in females, per log reduction in APCHI* | 0.43 | 0.66 | [37-39, 74] |
| Increase in casual sex in men who are employed (vs unemployed) | 0.25 | 0.40 | [31, 34, 74] |
| Commercial sex |  |  |  |
| RR of commercial sex in men who are employed | 1.40 | 0.63 | [35, 75, 76] |
| RR of commercial sex in women who are employed | 0.00 | - | [77] |
| Short-term, non-cohabiting relationships |  |  |  |
| RR of partner acquisition in men who are employed | 1.32 | 0.43 | [30, 36, 74] |
| Marital and cohabiting relationships |  |  |  |
| RR of marriage if currently in school | 0.37 | 0.34 | 1993 OHS |
| RR of male marriage if completed high school (no tertiary) | 1.50 | - | [78] |
| RR of male marriage if completed tertiary education | 3.50 | - | [78] |
| RR of female marriage if completed high school | 0.70 | - | [26, 78, 79] |
| Condom use |  |  |  |
| OR for consistent condom use per year of schooling | 1.05 | 0.08 | [80, 81] |
| Health seeking behaviour |  |  |  |
| RR of medical male circumcision per log increase in APCHI* | 1.15 | 0.19 | [40, 41] |
| RR of HIV testing per year of education | 1.12 | - | [82] |
| RR of hormonal contraceptive use per year of education | 1.15 | - | [80] |
| Intervention effectiveness |  |  |  |
| RR of school dropout if receiving school support (non-material) | 0.68 | 0.18 | [83] |
| RR of school dropout per R800 of school support | 0.86 | 0.10 | [84] |
| Probability of return to school if aged 13-22 and receiving school support | 0.34 | - | [64] |
| OR of unemployment if receiving vocational training/microfinance | 0.87 | 0.10 | [85] |

\* Per unit difference between the natural log of the APCHI and the log of the national average APCHI, for households that have a log APCHI below the national average (for those above the average, no income effect is modelled).
APCHI = adjusted per capita household income. OHS = October Household Survey. OR = odds ratio. RR = relative rate. SD = standard deviation.

### Model calibration

We adopted a Bayesian approach to model calibration [86], similar to that in a previous application of our model [55]. Prior distributions were specified to represent the uncertainty in key model parameters (Table 1). Hurdle distributions were specified to represent the uncertainty in the effect of socio-economic status on sexual risk behaviour, with the hurdle representing a non-zero probability of a null relationship. The non-zero probabilities were set based on a review of the strength of evidence from randomized controlled trials (RCTs); a more detailed description of the prior distributions is provided in section 2.1 of the supplementary materials.

The model was calibrated to the results of RCTs of economic strengthening interventions conducted in sub-Saharan Africa. RCTs were identified from recent reviews of economic strengthening interventions [44, 45, 47, 48]. Included trials were classified as being either ‘pure’ cash transfer interventions (including both conditional and unconditional cash transfers, but without any strong conditioning on school attendance), school support interventions (which typically aimed to promote school retention, often through the provision of financial support, or through cash transfers conditional on school attendance) and vocational training programmes (directed to individuals who were out of school, providing training to improve their employment prospects and/or credit to enable them to establish their own business). Intervention effects were calculated on a natural log scale, with associated standard errors (Supplementary Table S28).

A sample of 5000 parameter combinations was randomly drawn from the prior distributions in Table 1. For each parameter combination, the model was run eight times: twice in the ‘baseline’ scenario, twice in the cash transfer scenario, twice in the school support scenario and twice in the vocational training scenario (two simulations being necessary in order to quantify the extent of stochastic variation in model outputs; the average of the two results was calculated for the purpose of estimating intervention effects). For the purpose of calibration to historic RCT data, all interventions were assumed to start in 2005 (close to the average start year of the included RCTs), and the annual value of the cash transfer in 2005 was set to R800 (equivalent to $117 in 2005), the average value of the cash transfers in the included RCTs. The interventions were assumed to be directed to households with log APCHI below the national average (8.68 in 2005, equivalent to R5856 or $488 annually). The modelled intervention effect was calculated as the log of the odds ratio/relative risk for the outcome of interest (comparing the intervention and baseline scenarios). A likelihood value was calculated by comparing the modelled intervention effect and the reported intervention effect, assuming the log difference followed a normal distribution with zero mean. Log likelihood values were summed for all outcomes, across all RCTs, to calculate a total likelihood for each parameter combination; separate likelihoods were also calculated for each of the three intervention types.

A posterior sample of 50 parameter combinations was drawn from the initial set of 5000 parameter combinations, using the likelihood values as weights. This sample was used to calculate the posterior means and 95% confidence intervals (CIs), running the model five times for each parameter combination to reduce stochastic variation.

The model was validated by comparing the modelled odds ratios for the associations between socio-economic status and HIV/sexual risk behaviour, as observed in four nationally-representative household surveys in 2005, 2012, 2016 and 2017 [74, 87].

### Model scenarios

In all scenarios, STI transmission probabilities were set at the median of the distribution of best-fitting parameters identified in previous model calibrations [88]. HIV transmission probabilities per sex act were adjusted to yield plausible estimates of HIV incidence and prevalence (see section 3.2 of the supplementary materials). Teenage fertility rates, per year of sex with a single partner in the absence of contraception, were estimated from age- and race-specific fertility rates in the baseline scenario [89].

We calculated the proportion of incident HIV cases/STIs/teenage births that were attributable to low socio-economic status, over the 2000-2020 period, by running a counterfactual scenario in which the sexual behaviour and health seeking behaviour of all individuals are the same as might be expected (a) if they had completed tertiary education, (b) if they remained in education at least to age 21, (c) if they were employed, and (d) if their adjusted per capita household income were no lower than the national mean in the baseline scenario (with the changes in behaviour occurring from mid-2000). The population attributable fraction (PAF) was calculated as the proportionate difference in cumulative HIV cases/STIs/teenage births, over the 2000-2020 period, between this counterfactual scenario and the baseline scenario. Partial rank correlation coefficients (PRCCs) were calculated to assess associations between PAFs and each of the parameters that were changed in the counterfactual scenario [90].

Future intervention impacts were considered over the period from 2025-2040. Interventions were assumed to be limited to households with adjusted per capita household income below the national average in the baseline scenario. The annual value of the cash transfer was set to $227 per household in the case of cash transfer interventions and $114 per eligible youth in the case of school support (at 2023 exchange rates), increasing in line with inflation [91].

## Results

### Calibration to economic and reproductive health data

Our model was in reasonable agreement with observed HIV prevalence and incidence trends in South Africa, external estimates of the incidence of gonorrhoea and chlamydia [92] and data on teenage fertility (Figure 2a-d). The model was also in agreement with census and survey data showing rising levels of high school completion among youth, and stable employment levels (Figure 2e-f). The model also estimated rising income together with declining income inequality, the result of increasing levels of social welfare expenditure (Figures 2g-h).

**Figure 2:**
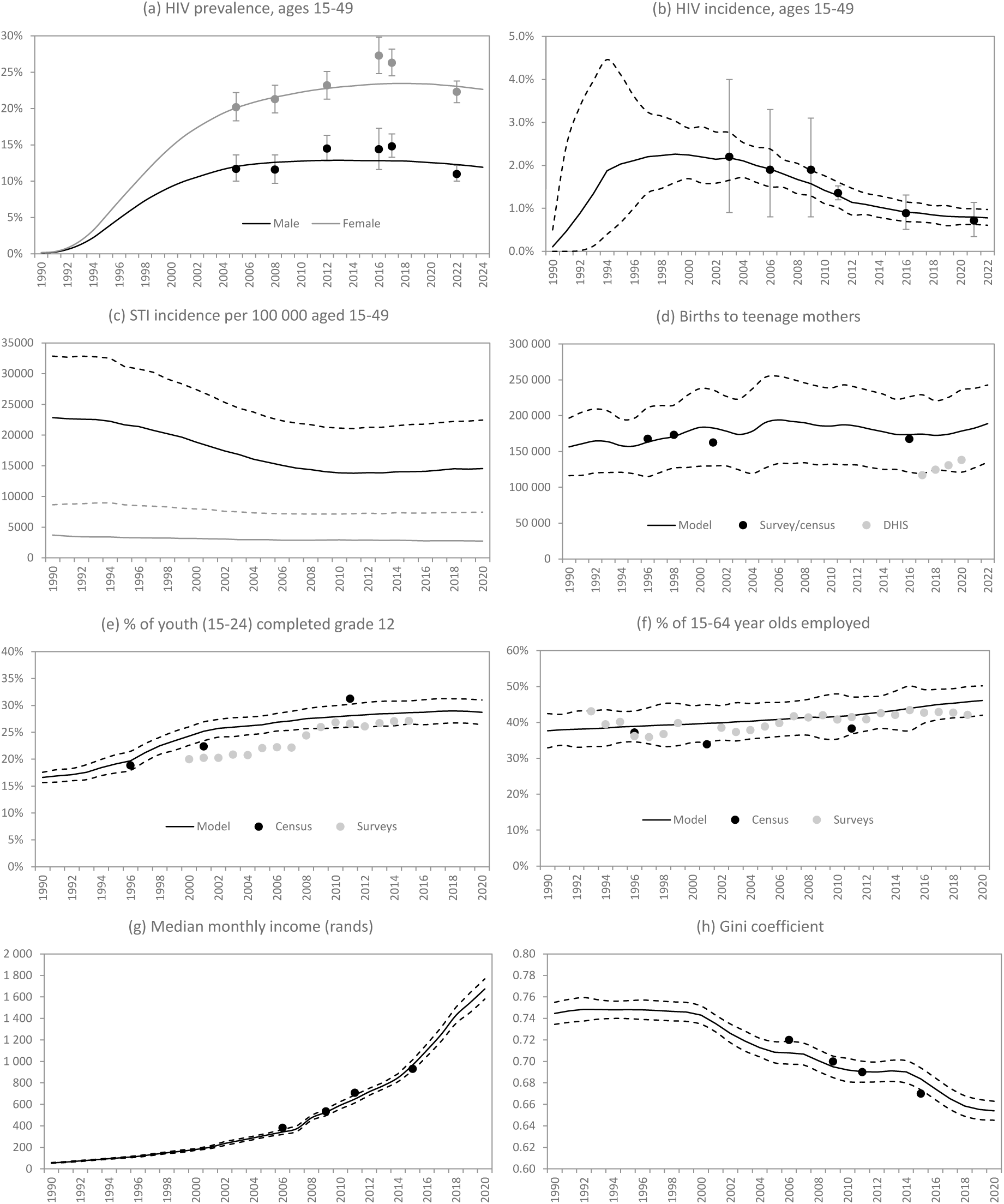
Model calibration. DHIS = District Health Information System, HSV-2 = herpes simplex virus type 2, STI = sexually transmitted infection. Except in panel c, solid lines represent posterior mean model estimates and dashed lines represent 95% confidence intervals. Calibration data points in panels a and b are from national household surveys [74, 87, 126], while data in panel c are from the DHIS [127], censuses and surveys [74, 128]. Data in panels e and f are from censuses, October Household Surveys, General Household Surveys and Labour Force Surveys. Data in panels g and h are derived from national surveys [129].

### Calibration to intervention effectiveness data

Posterior estimates of model parameters were largely consistent with prior distributions, although intervention effect parameters differed significantly: vocational training was estimated to be less effective in reducing unemployment (posterior OR 0.94, prior OR 0.87), while posterior estimates of school support impacts were mixed, with material support being more effective, compared to prior assumptions about effects on school dropout (Table S29). The posterior estimates of RCT effects were in good agreement with the data, although the model was unable to match the large reductions in HIV and herpes prevalence and pregnancy incidence in one trial [64] (Figures S13-S15). The model was also consistent with most of the validation data, although the model under-estimated the strength of the negative association between HIV prevalence and education in women (Figure S16).

### Population attributable fractions

Low socio-economic status accounted for 14% of new HIV infections, 10% of incident STIs (gonorrhoea, chlamydia and trichomoniasis) and 46% of teenage births in South Africa, over 2000-2020 (Table 2). However, because of uncertainties regarding effect sizes, confidence intervals around these PAFs were wide (3-32%, 0-21% and 12-89% respectively). PAF estimates were similar for men and women. Table 3 shows the socioeconomic effect parameters that account for the most uncertainty. The effect of education on condom use was the most significant correlate of the HIV PAF (PRCC=0.94) and the STI PAF (PRCC=0.93), but was less significant as a correlate of the teenage birth PAF (PRCC=0.49). The relative rate of female sexual debut while in school was the most significant correlate of the teenage birth PAF (PRCC=-0.96) and was also significantly correlated with the HIV and STI PAFs. The effects of male employment on casual sex and commercial sex were both significant determinants of the STI PAF, and the effect of household income on female entry into casual sex significantly influenced both the HIV and teenage birth PAFs. Other parameters were not statistically significant.

**Table 2:** Proportion of incident HIV cases, STIs and teenage births attributable to low socio-economic status, 2000-2020.

|  | HIV | STIs | Teenage births |
| --- | --- | --- | --- |
| Total | 14% (3 to 32%) | 10% (0 to 21%) | 46% (12 to 89%) |
| Males | 13% (-1 to 31%) | 9% (0 to 21%) | - |
| Females | 16% (2 to 33%) | 11% (0 to 23%) | 46% (12 to 89%) |

**Table 3:**
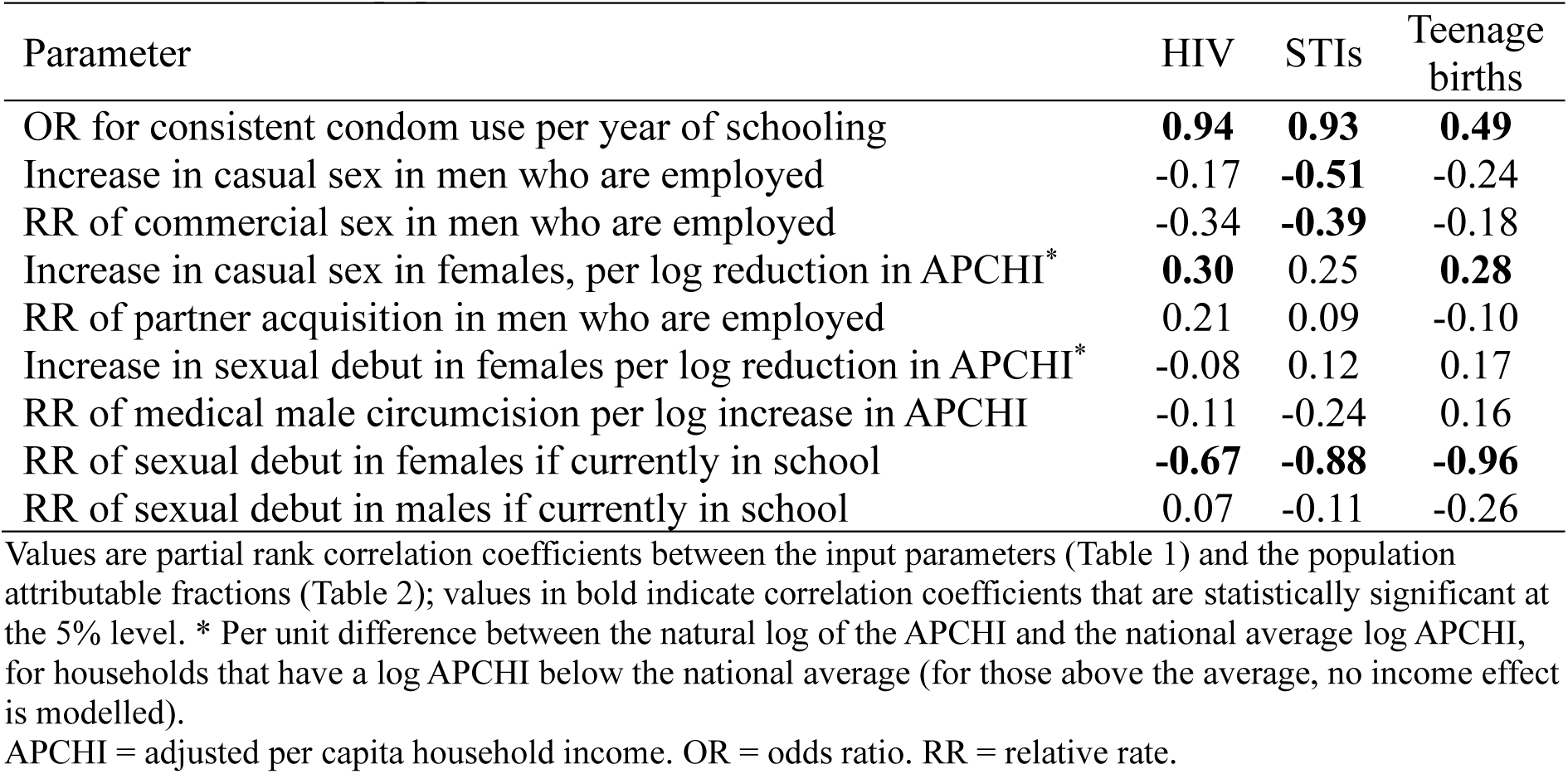
Correlates of population attributable fractions.

| Parameter | HIV | STIs | Teenage births |
| --- | --- | --- | --- |
| OR for consistent condom use per year of schooling | **0.94** | **0.93** | **0.49** |
| Increase in casual sex in men who are employed | -0.17 | **-0.51** | -0.24 |
| RR of commercial sex in men who are employed | -0.34 | **-0.39** | -0.18 |
| Increase in casual sex in females, per log reduction in APCHI* | **0.30** | 0.25 | **0.28** |
| RR of partner acquisition in men who are employed | 0.21 | 0.09 | -0.10 |
| Increase in sexual debut in females per log reduction in APCHI* | -0.08 | 0.12 | 0.17 |
| RR of medical male circumcision per log increase in APCHI | -0.11 | -0.24 | 0.16 |
| RR of sexual debut in females if currently in school | **-0.67** | **-0.88** | **-0.96** |
| RR of sexual debut in males if currently in school | 0.07 | -0.11 | -0.26 |

APCHI = adjusted per capita household income. OR = odds ratio. RR = relative rate.

### Intervention impacts

In the absence of any economic strengthening interventions, our model suggests there would be little change in income inequality and high school completion over the next 15 years, although employment is expected to increase modestly as a result of population aging (Figure 3). The cash transfer intervention would reduce income inequality, with the Palma ratio (the ratio of income in the top decile to that in the four lowest deciles) decreasing from 9.1 to 8.2 by 2030 (Figure 3a). The school support intervention would significantly increase the proportion of black South African youth who have completed high school, from 27% to 35% by 2040 (Figure 3b). Vocational training interventions would only slightly increase employment levels (Figure 3c).

**Figure 3:**
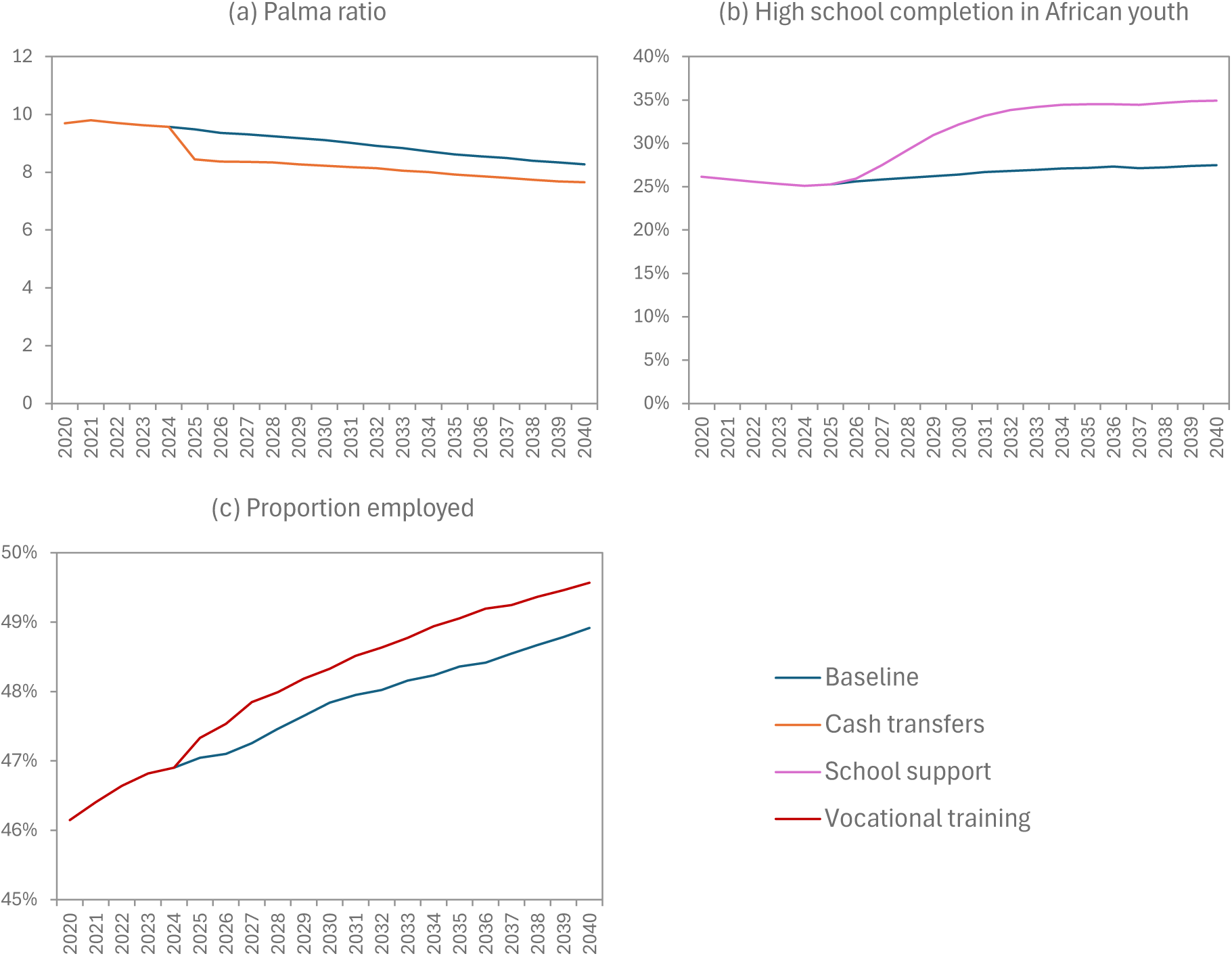
Intervention impacts on socio-economic outcomes. The Palma Ratio (panel A) is defined as the ratio of total income in the top income decile to the total income in the four lowest deciles. In panel (b), the high school completion fraction is the proportion of all 15-24 year old black South Africans who have successfully completed grade 12. In panel (c) the denominator is all individuals aged 15-64 (including those in school and those not actively seeking work).

Our model suggests that economic strengthening interventions would have no significant impact on most reproductive health outcomes (Table 4). Over 2025-2040, schooling support is the intervention that would come closest to achieving a significant impact, with an anticipated 4% reduction in teenage births (which is strongly correlated with the relative rate of sexual debut for girls who remain in school [PRCC=-0.73]) and a 1% reduction in STI incidence (which is strongly correlated with the relative rate of sexual debut among girls in school [PRCC=-0.55] and the increase in condom use per year of education [PRCC=0.50]). For all other intervention/outcome combinations, the expected impact is 0%, with lower confidence interval limits below zero (partly a reflection of residual stochastic variation).

**Table 4:** Reduction in key outcomes (2025-2040) as a result of different economic strengthening interventions.

|  | New HIV infections (15+) |  |  | New STIs (15+) |  |  | Births to teenagers |
| --- | --- | --- | --- | --- | --- | --- | --- |
|  | Total | Male | Female | Total | Male | Female |  |
| School support | 0% (-4 to 4) | 0% (-4 to 6) | 0% (-4 to 3) | 1% (0 to 1) | 0% (0 to 1) | 1% (0 to 2) | 4% (-1 to 15) |
| Vocational training | 0% (-4 to 3) | 0% (-5 to 4) | -1% (-4 to 3) | 0% (-1 to 1) | 0% (-1 to 1) | 0% (-1 to 1) | 0% (-5 to 6) |
| Cash transfers | 0% (-3 to 4) | 0% (-4 to 5) | 0% (-3 to 5) | 0% (-1 to 0) | 0% (-1 to 0) | 0% (-1 to 1) | 0% (-3 to 2) |

95% confidence intervals are in brackets.

Vocational training is unlikely to have a significant impact if it is limited to females (for example, a 0% reduction [95% CI: -1 to 1] in STIs). Intervention impacts are similar in males and females. Proportionate reductions in HIV, STIs and teenage pregnancies are also similar over the 2025-2040 period, i.e. there is no suggestion of the intervention impact waning or improving at longer durations (results not shown).

## Discussion

Although our model suggests a potentially significant contribution of low socio-economic status to the incidence of HIV, STIs and teenage pregnancy in South Africa, there is substantial uncertainty around the magnitude of this contribution. This uncertainty arises because socio-economic status affects sexual behaviour in diverse ways. On the one hand, our model identifies significant positive effects of the relationship between condom use and educational attainment, the relationship between schooling and sexual debut and the relationship between household income and female engagement in casual/transactional sex. On the other hand, we find that increases in male socioeconomic status can significantly increase STI incidence because of the effect of male employment on entry into casual and commercial sex relationships. Because of these offsetting effects, the net effects of economic strengthening on sexual and reproductive health outcomes can be modest.

One possible response to this complexity would be to target economic strengthening to those groups that are most likely to be positively affected, for example economically vulnerable adolescent girls and young women. While this may have merit, our simulations suggest that vocational training targeted to women (for example) would not significantly alter HIV and STI incidence. We also find that economic strengthening interventions have similar effects in men and women, despite very different socio-economic effects on sexual behaviour. This is largely because HIV and STI transmission in South Africa is predominantly heterosexual (i.e. any gain by one sex in the short term is likely to result in reduced secondary transmission to the other sex in the longer term).

The disappointing lack of impact of economic strengthening interventions on reproductive outcomes is to some extent a reflection of the difficulty in achieving significant changes in socioeconomic status, against a background of extreme economic inequality. For example, vocational training interventions would increase employment levels by only 1%, and cash transfers to the poorest South African household would only reduce the Palma ratio of inequality by 8% (Figure 3). More radical structural changes may be needed to achieve substantial gains in economic and reproductive health outcomes.

There have been few previous attempts to develop mathematical models of the structural drivers of sexual risk behaviour [52], and even fewer that have specifically assessed the role of socioeconomic status. A few studies have modelled the effects of poverty and social protection in Brazil [93, 94], but these are calibrated based on observed temporal associations between economic indicators and HIV indicators, not on the results of individual-level associations or RCTs of economic strengthening. A strength of our calibration approach is that we have used a Bayesian approach to combine both local observational data on the likely relationships between socio-economic status and sexual behaviour (through prior distributions) and RCT data on the impacts of economic strengthening interventions in African settings (through a likelihood function). It is perhaps disappointing that the resulting confidence intervals around the posterior model estimates remain wide, despite the systematic approach to including different types of evidence, but we believe this reflects real uncertainty around socio-economic relationships.

A limitation of this model is that we have focused mainly on the effects of socio-economic status on sexual behaviour and have ignored some of the other effects that are relevant to sexual and reproductive health outcomes. For example, we have not modelled effects of socio-economic status on mortality in people living with HIV [42]. We have also not modelled an effect of socio-economic status on STI health seeking [95], although in the South African setting higher socio-economic status is not necessarily associated with better STI treatment [96, 97]. We have nevertheless included socioeconomic effects on hormonal contraceptive use, male circumcision and rates of HIV testing, all of which are important. We have not modelled some of the more detailed causal pathways that link socio-economic status and sexual risk behaviour. For example, there is evidence of a relationship between food insecurity and sexual risk behaviour, particularly in women [38, 98–100]. In addition, poverty is associated with greater mental distress [101–103], which in turn may be linked to higher sexual risk behaviours [102, 104, 105]. Education is also associated with exposure to life skills programmes and HIV prevention messaging [106].

Another limitation is that we have not modelled the effects of COVID-related lockdowns on education [107], employment [108], ART uptake [109] and contraceptive use [110]. The adverse social effects of the COVID pandemic were to some extent mitigated by the introduction of a Social Relief of Distress grant, which could be accessed by adults who were unemployed and not receiving any other income [111], and this grant has been continued into the post-COVID period. There have been calls to make this grant permanent, and to increase the amount of the grant [111]. A detailed analysis of the impact of COVID on sexual and reproductive health is beyond the scope of this study, as is the analysis of the potential impact of the Social Relief of Distress grant and its continuation. However, it is worth noting that in our cash transfer scenario we have considered a grant that is worth $227 per annum in 2023, which is similar to the $223 per annum currently under the Social Relief of Distress grant.

Given the modest HIV and STI impacts estimated in our cash transfer scenario, it seems unlikely that the Social Relief of Distress grant has had much impact on sexual and reproductive health outcomes.

This study considers the role of socio-economic factors at the individual and household level, but socio-economic characteristics of the community may also be important in driving sexual risk behaviour, even when controlling for individual socio-economic status [112, 113]. In South Africa, income inequality at the municipal level significantly increases women’s risk of HIV [114], and income inequality has also been shown to explain much of the variation across countries in sexual risk behaviour [115], HIV prevalence [20, 116, 117] and teenage fertility [3]. Further work is required to understand (and potentially model) these community effects. Our results might not be generalizable to other African settings, where there is typically less income inequality but more absolute poverty.

Economic interventions can have important positive impacts beyond sexual and reproductive health, including on mental health [71, 118–120], tuberculosis [121] and child health [122]. Ideally, all of these health outcomes should be considered when evaluating the impact or cost-effectiveness of economic interventions, and we do not suggest that policy decisions should be based on sexual and reproductive health outcomes alone. There is a need for a more inter-sectoral ‘whole of government’ approach to addressing health challenges [123], and this is especially true when considering economic strengthening interventions. This necessitates more advanced analytic tools, both in cost-effectiveness evaluation [124] and in simulation of complex systems [125]. This study represents a first step towards capturing this inter-sectoral complexity in a model of a middle-income country with significant income inequality.

## Data Availability

This modelling study makes use of estimates from published studies, which are cited in the paper and supplementary materials. The code is available from the corresponding author on request.

