## supplementary for "Money, jobs or schooling? A model-based evaluation of economic strengthening in South Africa and its impact on HIV, sexually transmitted infections and teenage births"

### Supplementary materials

#### Contents

1. Extensions to the MicroCOSM model structure ... 3
1.1 Modelling employment status ... 3
1.1.1 Socio-demographic determinants of employment ... 3
1.1.2 HIV effects ... 7
1.1.3 Starting employment profile ... 7
1.1.4 Calibration ... 8
1.1.5 Limitations ... 9
1.2 Modelling household formation ... 10
1.2.1 Definitions ... 10
1.2.2 Initial assignment to households in 1985 ... 11
1.2.3 Modelling changes in household membership ... 14
1.2.4 Homelessness ... 20
1.2.5 Model calibration and validation ... 22
1.2.6 Limitations ... 23
1.3 Modelling household income ... 24
1.3.1 Salary/wage income ... 25
1.3.2 Child support grants ... 26
1.3.3 State old age pension ... 29
1.3.4 Private pensions ... 29
1.3.5 Model validation ... 30
1.3.6 Limitations ... 31
1.4 Effects of socio-economic status on sexual behaviour ... 32
1.4.1 Sexual debut ... 35
1.4.2 Casual/transactional sex ... 37
1.4.3 Commercial sex ... 40
1.4.4 Short-term non-cohabiting relationships ... 43
1.4.5 Condom use ... 47
1.4.6 Marriage/cohabitation ... 50
1.5 Other socio-economic effects ... 56
1.5.1 Effects on hormonal contraceptive use ... 56
1.5.2 Effect of socio-economic status on male circumcision ... 58

### 1. Extensions to the MicroCOSM model structure

The structure of the MicroCOSM model and its assumptions has been described in detail previously [1]. The sections that follow describe recent extensions to the model structure that are relevant to modelling the role of socio-economic factors in the epidemiology of HIV, STIs and teenage pregnancy in South Africa.

#### 1.1 Modelling employment status

In our model, all individuals aged 15-64 are classified as employed or unemployed. For simplicity, we assume that all individuals aged 65 and older are retired, and that individuals who are currently in school or attending a tertiary education institution are not employed. Our definition of 'employed' is the same as that used in the national Labour Force Surveys: individuals are classified as employed if they have done any work for pay in the last week, if they ran a business in the last week, or if they had a temporary absence from work (for example due to sick leave, maternity leave or vacation).

As in the previously described MicroCOSM model [2], we update employment status at annual time steps, using estimates of employment rates in the Labour Force Surveys. However, our approach differs from that adopted previously in that we assume the employment probability at each annual update depends on the employment status one year previously. In addition, we assume that employment probability depends on the individual's HIV status and (in the case of women) the degree of childcare responsibility at a household level. We define the latter as the number of children (under the age of 15) per woman aged 15 or older in the household, recognizing that current gender norms place the burden of childcare mostly on women.

##### 1.1.1 Socio-demographic determinants of employment

Multivariable logistic regression models were applied to the Quarter 3 Labour Force Surveys, for each year from 2008 to 2019. Quarter 3 was chosen as it corresponds most closely to the middle of each year, which is when our model updates employment status. Because the surveys do not include a direct question on employment status 12 months prior, we created a composite indicator based on other questions: in the case of individuals who were currently employed, we defined them as being unemployed one year previously if they reported having started their current job in the current year, and in the case of individuals who were currently unemployed, we defined them as being unemployed one year previously if they reported having been unemployed for 12 months or longer. This is not an accurate reflection of the individual's true employment status 12 months previously, but is intended to capture roughly the dependence of current employment on past employment history.

Table S1 shows the results of the multivariate logistic regression models. The regression coefficients are fairly similar across surveys; in every survey, the odds of employment are strongly associated with male sex, urban location, middle age (peaking in the 35-49 age group), higher education, white race, lower childcare responsibility (in women) and previous employment. The high odds ratios for previous employment indicate that current employment is strongly dependant on past employment, even after controlling for education, age and other socio-demographic factors. Results are consistent with previous South African longitudinal analyses of changes in employment status [3], which similarly found that being unemployed increased the odds of being unemployed one year later (aOR 13.9, 95% CI: 10.7-18.2).

In our model we assume that the effects of age, sex, urban location, education, race, childcare responsibility and previous employment on the odds of employment remain constant over time, and we use the average odds ratios over the 2008-2019 period (final column of Table S1) to set these assumed effects. However, in the white population we assume a stronger effect of female sex on employment (OR 0.46 instead of 0.72), in order to bring the model more in line with survey data. The baseline odds of employment (the odds of employment in a rural 15-19-year old black male with no education and no employment 12 months previously) is kept constant at 0.06, although it could be allowed to vary if employment trends changed substantially over time.

Table S1: Predictors of employment in the South African Labour Force Surveys

| | 2008 | 2009 | 2010 | 2011 | 2012 | 2013 | 2014 |
|---|---|---|---|---|---|---|---|
| Sex | | | | | | | |
| Male | 1 | 1 | 1 | 1 | 1 | 1 | 1 |
| Female | 0.63 (0.59-0.67) | 0.75 (0.70-0.80) | 0.69 (0.64-0.75) | 0.71 (0.65-0.76) | 0.70 (0.65-0.76) | 0.80 (0.74-0.86) | 0.73 (0.67-0.78) |
| Location | | | | | | | |
| Rural | 1 | 1 | 1 | 1 | 1 | 1 | 1 |
| Urban | 1.41 (1.32-1.50) | 1.33 (1.24-1.42) | 1.45 (1.35-1.57) | 1.44 (1.33-1.55) | 1.30 (1.21-1.40) | 1.13 (1.05-1.22) | 1.20 (1.12-1.29) |
| Age group | | | | | | | |
| 15-19 | 1 | 1 | 1 | 1 | 1 | 1 | 1 |
| 20-24 | 1.56 (1.29-1.89) | 1.97 (1.57-2.47) | 2.29 (1.74-3.01) | 2.32 (1.81-2.97) | 2.04 (1.56-2.67) | 1.88 (1.40-2.51) | 1.57 (1.19-2.06) |
| 25-29 | 2.45 (2.03-2.96) | 3.14 (2.51-3.92) | 3.59 (2.74-4.70) | 3.35 (2.63-4.26) | 3.38 (2.60-4.40) | 2.71 (2.04-3.61) | 2.68 (2.05-3.49) |
| 30-34 | 3.30 (2.73-3.99) | 3.88 (3.10-4.86) | 5.05 (3.85-6.62) | 4.63 (3.62-5.90) | 4.42 (3.39-5.75) | 3.99 (2.99-5.33) | 3.35 (2.56-4.38) |
| 35-39 | 3.63 (3.00-4.40) | 4.51 (3.60-5.64) | 5.24 (3.99-6.88) | 5.22 (4.09-6.66) | 4.95 (3.79-6.47) | 4.32 (3.24-5.77) | 3.86 (2.95-5.04) |
| 40-44 | 3.13 (2.58-3.78) | 5.32 (4.25-6.65) | 5.93 (4.52-7.78) | 5.08 (3.98-6.49) | 5.23 (4.00-6.82) | 4.50 (3.37-6.01) | 4.15 (3.18-5.43) |
| 45-49 | 3.36 (2.78-4.06) | 4.36 (3.48-5.47) | 4.78 (3.65-6.27) | 5.09 (3.99-6.50) | 4.61 (3.53-6.01) | 4.28 (3.19-5.73) | 3.68 (2.81-4.82) |
| 50-54 | 2.58 (2.13-3.13) | 3.73 (2.95-4.70) | 4.41 (3.37-5.78) | 4.20 (3.30-5.35) | 4.18 (3.19-5.47) | 3.67 (2.75-4.90) | 3.62 (2.77-4.73) |
| 55-59 | 1.84 (1.52-2.23) | 2.69 (2.14-3.39) | 3.26 (2.47-4.31) | 3.40 (2.66-4.34) | 3.15 (2.41-4.10) | 2.71 (2.02-3.64) | 2.82 (2.15-3.70) |
| 60-64 | 0.86 (0.70-1.05) | 1.32 (1.04-1.68) | 1.41 (1.06-1.86) | 1.20 (0.93-1.55) | 1.21 (0.92-1.60) | 1.18 (0.87-1.60) | 1.26 (0.96-1.66) |
| Education | | | | | | | |
| No education | 1 | 1 | 1 | 1 | 1 | 1 | 1 |
| Primary only | 0.96 (0.85-1.08) | 0.93 (0.81-1.06) | 1.00 (0.85-1.18) | 1.08 (0.92-1.28) | 1.18 (1.01-1.37) | 0.99 (0.84-1.17) | 0.97 (0.83-1.13) |
| Incomplete secondary | 1.20 (1.07-1.34) | 1.30 (1.15-1.47) | 1.46 (1.26-1.70) | 1.36 (1.18-1.57) | 1.32 (1.14-1.54) | 1.03 (0.87-1.21) | 1.12 (0.97-1.29) |
| Completed secondary | 1.26 (1.11-1.44) | 1.32 (1.14-1.52) | 1.35 (1.14-1.59) | 1.33 (1.13-1.56) | 1.97 (1.68-2.30) | 1.57 (1.33-1.86) | 1.78 (1.53-2.07) |
| Tertiary | 1.63 (1.32-2.03) | 2.22 (1.77-2.79) | 2.79 (2.17-3.59) | 2.40 (1.89-3.04) | 4.93 (3.81-6.39) | 4.21 (3.29-5.38) | 3.57 (2.85-4.48) |
| Race | | | | | | | |
| Black African | 1 | 1 | 1 | 1 | 1 | 1 | 1 |
| 'Coloured' | 1.08 (0.99-1.19) | 1.08 (0.98-1.19) | 1.15 (1.04-1.28) | 1.01 (0.91-1.11) | 1.06 (0.95-1.18) | 1.12 (1.02-1.24) | 1.27 (1.15-1.41) |
| Indian/Asian | 1.21 (1.02-1.42) | 1.13 (0.96-1.33) | 1.69 (1.41-2.03) | 1.14 (0.94-1.37) | 1.01 (0.83-1.22) | 1.01 (0.84-1.21) | 1.05 (0.89-1.25) |
| White | 2.19 (1.95-2.45) | 2.03 (1.78-2.32) | 2.14 (1.90-2.40) | 2.01 (1.78-2.28) | 1.58 (1.38-1.81) | 1.62 (1.41-1.87) | 1.51 (1.31-1.75) |
| Number of children per woman | 0.78 (0.75-0.81) | 0.80 (0.76-0.84) | 0.82 (0.78-0.86) | 0.80 (0.76-0.84) | 0.81 (0.77-0.85) | 0.79 (0.75-0.83) | 0.80 (0.76-0.84) |
| Recent employment | | | | | | | |
| Not employed a year ago | 1 | 1 | 1 | 1 | 1 | 1 | 1 |
| Employed a year ago | 13.9 (13.1-14. 7) | 16.17 (15.2-17.2) | 21.4 (20.0-22.8) | 26.2 (24.6-28.0) | 21.5 (20.1-23.0) | 19.4 (18.2-20.6) | 18.8 (17.6-20.1) |
| Baseline odds | 0.13 (0.10-0.15) | 0.06 (0.05-0.08) | 0.04 (0.03-0.05) | 0.04 (0.03-0.05) | 0.05 (0.03-0.06) | 0.07 (0.05-0.09) | 0.07 (0.05-0.09) |

Table S1: Predictors of employment in the South African Labour Force Surveys (continued)

| | 2015 | 2016 | 2017 | 2018 | 2019 | Average (min-max) |
|---|---|---|---|---|---|---|
| Sex | | | | | | |
| Male | 1 | 1 | 1 | 1 | 1 | 1 |
| Female | 0.69 (0.64-0.74) | 0.69 (0.64-0.74) | 0.71 (0.66-0.77) | 0.76 (0.70-0.82) | 0.79 (0.73-0.86) | 0.72 (0.63-0.80) |
| Location | | | | | | |
| Rural | 1 | 1 | 1 | 1 | 1 | 1 |
| Urban | 1.38 (1.29-1.48) | 1.37 (1.27-1.47) | 1.29 (1.20-1.39) | 1.42 (1.32-1.53) | 1.27 (1.18-1.37) | 1.33 (1.13-1.45) |
| Age group | | | | | | |
| 15-19 | 1 | 1 | 1 | 1 | 1 | 1 |
| 20-24 | 1.65 (1.26-2.17) | 2.05 (1.51-2.78) | 1.50 (1.13-1.98) | 2.37 (1.73-3.24) | 1.63 (1.13-2.33) | 1.90 (1.50-2.37) |
| 25-29 | 2.37 (1.81-3.10) | 3.09 (2.30-4.17) | 2.05 (1.55-2.71) | 3.20 (2.35-4.35) | 2.64 (1.86-3.75) | 2.89 (2.05-3.59) |
| 30-34 | 3.16 (2.42-4.13) | 3.99 (2.96-5.37) | 2.49 (1.89-3.28) | 4.32 (3.18-5.86) | 3.57 (2.52-5.06) | 3.85 (2.49-5.05) |
| 35-39 | 3.34 (2.55-4.37) | 4.65 (3.44-6.27) | 3.01 (2.28-3.97) | 5.00 (3.68-6.79) | 4.39 (3.09-6.22) | 4.34 (3.01-5.24) |
| 40-44 | 3.76 (2.87-4.93) | 4.87 (3.61-6.57) | 3.02 (2.28-3.98) | 5.43 (3.99-7.40) | 4.55 (3.20-6.48) | 4.58 (3.02-5.93) |
| 45-49 | 3.53 (2.69-4.63) | 4.90 (3.62-6.61) | 3.12 (2.36-4.12) | 5.07 (3.73-6.90) | 4.30 (3.02-6.11) | 4.26 (3.12-5.09) |
| 50-54 | 2.72 (2.07-3.57) | 4.30 (3.19-5.80) | 2.79 (2.11-3.69) | 4.46 (3.27-6.09) | 3.74 (2.63-5.32) | 3.70 (2.58-4.46) |
| 55-59 | 2.02 (1.54-2.65) | 2.93 (2.17-3.95) | 2.12 (1.60-2.81) | 3.54 (2.60-4.83) | 2.82 (1.98-4.00) | 2.77 (1.84-3.54) |
| 60-64 | 0.90 (0.68-1.19) | 1.35 (0.99-1.84) | 0.96 (0.72-1.28) | 1.49 (1.09-2.04) | 1.22 (0.86-1.75) | 1.20 (0.86-1.49) |
| Education | | | | | | |
| No education | 1 | 1 | 1 | 1 | 1 | 1 |
| Primary only | 1.18 (1.01-1.37) | 1.11 (0.94-1.31) | 0.91 (0.77-1.08) | 1.05 (0.89-1.25) | 1.15 (0.96-1.37) | 1.04 (0.91-1.18) |
| Incomplete secondary | 1.11 (0.96-1.28) | 1.14 (0.98-1.34) | 0.97 (0.83-1.14) | 1.18 (1.00-1.39) | 1.07 (0.91-1.26) | 1.19 (0.97-1.46) |
| Completed secondary | 1.64 (1.42-1.90) | 1.62 (1.38-1.91) | 1.30 (1.11-1.53) | 1.47 (1.25-1.74) | 1.47 (1.25-1.74) | 1.51 (1.26-1.97) |
| Tertiary | 4.28 (3.47-5.27) | 3.31 (2.72-4.04) | 2.73 (2.25-3.30) | 3.31 (2.74-4.01) | 3.12 (2.58-3.78) | 3.21 (1.63-4.93) |
| Race | | | | | | |
| Black African | 1 | 1 | 1 | 1 | 1 | 1 |
| ‘Coloured’ | 1.17 (1.05-1.32) | 1.11 (0.99-1.25) | 1.02 (0.91-1.14) | 1.10 (0.98-1.24) | 1.20 (1.06-1.35) | 1.12 (1.01-1.27) |
| Indian/Asian | 1.01 (0.86-1.19) | 0.99 (0.84-1.15) | 1.12 (0.93-1.36) | 1.07 (0.88-1.30) | 1.12 (0.92-1.38) | 1.13 (0.99-1.69) |
| White | 1.73 (1.54-1.96) | 1.48 (1.29-1.70) | 1.49 (1.29-1.72) | 1.50 (1.31-1.72) | 1.73 (1.49-2.01) | 1.75 (1.48-2.19) |
| Number of children per woman | 0.83 (0.79-0.86) | 0.82 (0.78-0.86) | 0.88 (0.84-0.92) | 0.84 (0.80-0.88) | 0.81 (0.77-0.86) | 0.81 (0.78-0.88) |
| Recent employment | | | | | | |
| Not employed a year ago | 1 | 1 | 1 | 1 | 1 | 1 |
| Employed a year ago | 16.3 (15.4-17.3) | 20.5 (19.3-21.9) | 20.74 (19.5-22.1) | 22.4 (21.1-23.9) | 26.5 (24.9-28.4) | 20.3 (13.9-26.5) |
| Baseline odds | 0.08 (0.06-0.1) | 0.05 (0.04-0.07) | 0.09 (0.06-0.12) | 0.04 (0.03-0.06) | 0.05 (0.03-0.07) | 0.06 (0.04-0.13) |

Although the baseline odds of employment (in black South Africans) are assumed to remain constant over time, there is some evidence of a slight reduction in employment rates in white and coloured[1] South Africans after 2008 (discussed below), and we therefore specify different race effects for the period up to and after 2008 (Table S2).

Table S2: Assumed racial difference in employment parameters

| | OR for effect of race on entry into employment | | OR for effect of female sex on entry into employment |
|---|---|---|---|
| | Up to 2008 | Post-2008 | |
| Black African | 1 | 1 | 0.72 |
| 'Coloured' | 1.32 | 1.12 | 0.72 |
| White | 1.75 | 1.49 | 0.46 |

##### 1.1.2 HIV effects

South African studies on the effect of HIV on employment are scarce; most studies focus instead on the effect of employment on HIV risk. Levinsohn *et al* [4] analysed the data from the national household survey conducted by the HSRC in 2005, at a time when ART availability was limited, and found that among black South Africans there was, overall, a significant increase in unemployment among HIV-positive individuals (OR 1.064). However, the effect of HIV appeared to be much greater in individuals with an educational attainment below grade 12 (OR 1.105) than in individuals who had completed grade 12 (OR 0.994). This suggests that the effects of HIV might be more significant in less highly skilled professions, which are typically more manual and physically demanding. Studies also suggest that employment prospects increase after starting ART. In a study of mineworkers in Botswana, absenteeism was found to dip substantially in the 12 months prior to ART enrolment, then return to normal levels within 12 months after ART initiation [5]. Absenteeism was also found to increase significantly at CD4 counts of 350 or lower. Rosen *et al* [6] found that in a South African cohort of ART patients, the proportion who reported being employed increased from 32% at ART enrolment (typically when they had advanced HIV) to 44% by four years after ART initiation (unadjusted OR 1.67). Data from randomized controlled trials are conflicting, with one trial suggesting that early ART initiation (prior to advanced disease) leads to increased employment [7], and another trial finding no evidence of any effect [8].

Based on these studies, we assume that the odds of employment reduce by 10% in untreated individuals with CD4 counts of 200-350 cells/µl and by 40% in untreated individuals with CD4 counts <200 cells/µl, if they have not completed high school (no reduction in employment is assumed for individuals who have completed high school, on the assumption that they would be working in more highly skilled positions). This is roughly equivalent to a 10% reduction in employment among HIV-positive adults who have not completed high school in 2005, when using the proportions of HIV-positive adults in different CD4 categories in 2005 [9], consistent with the results of Levinsohn *et al* [4]. No reduction in employment is assumed for individuals who are on ART.

##### 1.1.3 Starting employment profile

We lack data on the employment profile of the South African population in 1985, the start year of our simulation. Our approach is therefore to use the same regression model that we

[1] The term 'coloured' is widely used and accepted in South Africa. We use it here rather than the more internationally accepted term 'mixed race', which is less accepted in the local context.

previously used (based on the 2015 Labour Force Survey [2]) to assign employment status in 1985. The starting employment profile has relatively little impact on the results after the first few years of the simulation.

##### 1.1.4 Calibration

Figure S1a shows that the modelled proportion of the working age population (ages 15-64) that is employed has remained relatively stable over time, with a slight increase from 38% in 1990 to 44% in 2019. The model estimates are roughly consistent with data from the October Household Surveys (1993-1999), the General Household Surveys (2002-2008), the Labour Force Surveys (2008-2019) and the censuses in 1996, 2001 and 2011, although there are a few outlier observations (such as the 1993 OHS, the 2001 census and the 2008 LFS). The different surveys and censuses use slightly different questions to assess employment status, so the overall consistency between the model and the data appears reasonable. The model also appears consistent with the age pattern of employment in 2015, although the model slightly under-estimates levels of employment in the youngest age groups (Figure 1b).

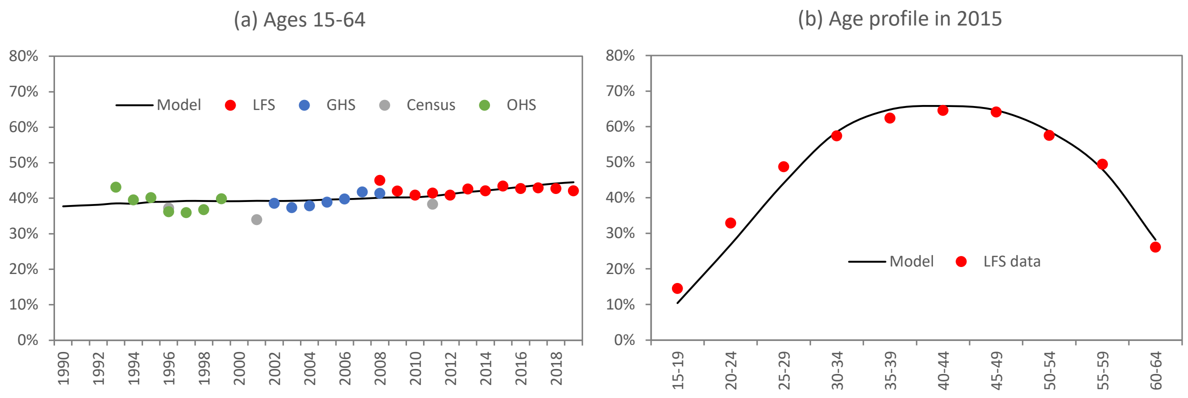

Figure S1: National levels of employment in South Africa, in the population aged 15-64
GHS = General Household Survey, LFS = Labour Force Survey, OHS = October Household Survey.

Figure S2 shows the model calibration to the same survey data, stratified by sex and race. In both men and women, employment levels are lowest in Africans, higher in coloureds and highest among whites. Employment levels are substantially lower in women than in men in all three population groups, although the differences are starkest in the white population. Overall the model appears roughly consistent with the data, although the model tends to slightly under-estimate levels of employment in coloured men in the 1990s. The assumed reduction in the relative odds of employment among whites and coloured, after 2008, appears roughly consistent with the observed trends in these population groups.

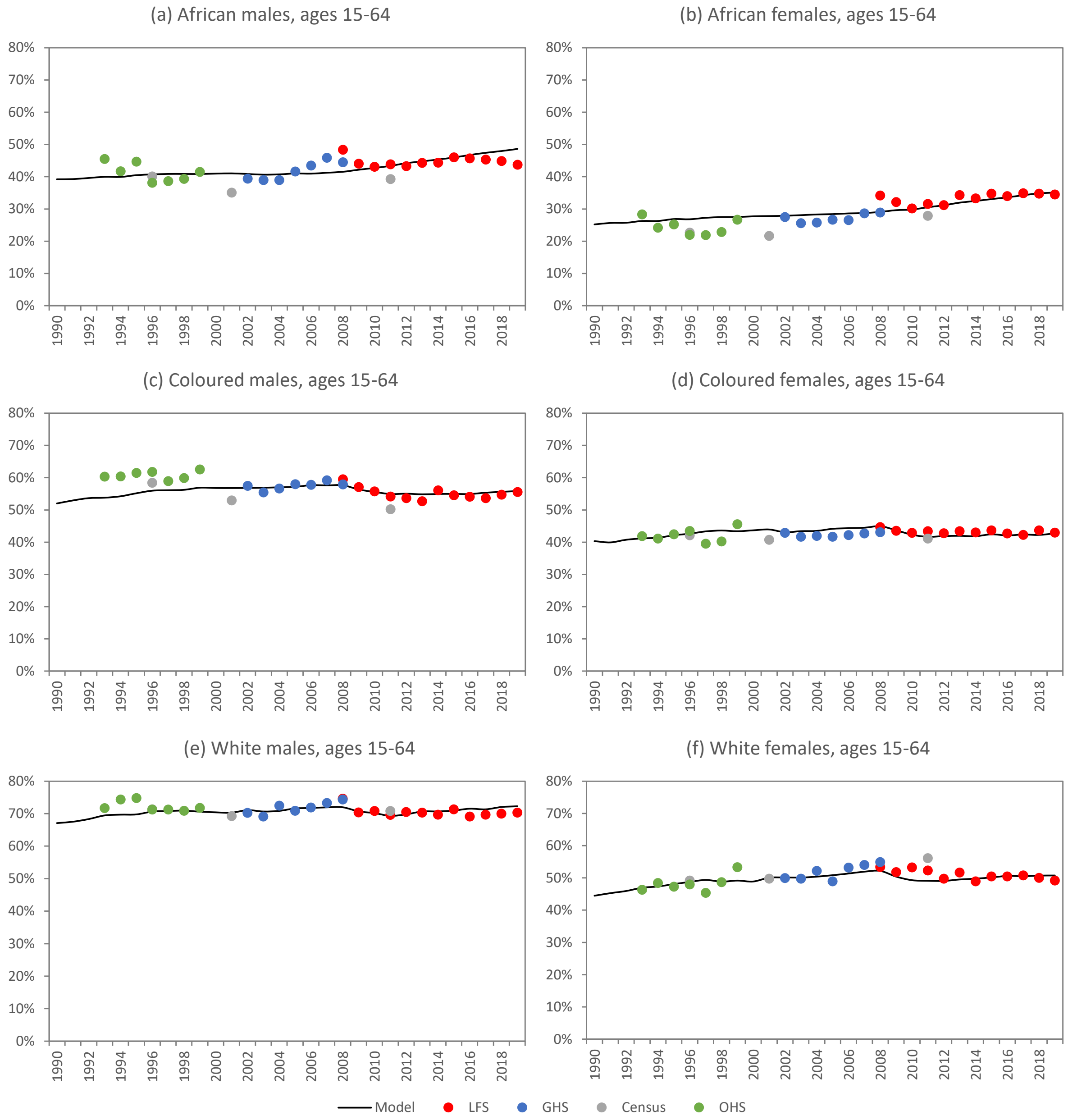

Figure S2: National trends in employment (ages 15-64), by sex and race
GHS = General Household Survey, LFS = Labour Force Survey, OHS = October Household Survey.

##### 1.1.5 Limitations

A limitation of this model is that it assumes the effects of sex on employment remain constant over time, although we do allow for a slight reduction in relative rates of employment among white and coloured South Africans. The introduction of affirmative action policies since the mid-1990s might have had led to a change in the effects of race and sex over time. Fredericks and Yu [10] found that although there was a substantial increase in female labour force participation and employment over the period from 1997 to 2016, there has been relatively little change in the difference in employment rates between black and white South Africans, when controlling for other predictors of employment. Some of the increase in women's employment may be attributable to the introduction of the Child Support Grant in 1998, which has been shown to be associated with increased female labour force participation [11, 12]. Our model has also been calibrated using data up to 2019, and does not consider the changes in employment that have occurred as a result of COVID-19 lockdowns [13].

Another limitation is that our model assumes individuals who are currently in school are not employed. In reality this may be untrue, especially in older youth, and this might account for some of the discrepancy between the model and the survey data in Figure S1b. However, when analysing the 2015 Labour Force Survey, we found that the fraction of individuals attending an educational institution who are also employed was only 0.2% and 3.6% among 15-19 and 20-24 year olds, respectively. This means that our model estimate of the total number of employed individuals in South Africa would not be much altered if we were also to allow for employment among individuals who are currently attending an educational institution.

Our model does not distinguish between unemployed individuals in the labour market and outside the labour market (i.e. we do not differentiate unemployed individuals according to whether they are actively seeking work). Other analyses have shown that most adults who are not unemployed are ‘not economically active’ (i.e. not seeking work) [14]. Our model also considers only paid work, and does not consider unpaid work. However, in the 2015 Labour Force Survey, only 0.2% of respondents reported unpaid work in the last week (as compared to 43.4% who reported either paid work, owning a business or a temporary absence from work). Because the model only updates employment status at one-year intervals, it implicitly ignores short-term changes in employment, for example due to seasonal work.

#### 1.2 Modelling household formation

##### 1.2.1 Definitions

A household is defined here as a group of individuals living in the same dwelling. For the sake of consistency with most of the household surveys conducted in South Africa, we define an individual to be a member of a household if they spend most nights in that household (in the General Household Surveys, for example, individuals are only included as members of a household if they have spent at least four nights per week in the house, on average over the last four weeks). This differs from the definition used in some surveys (for example the National Income Dynamics Survey), which includes members who are mostly resident outside of the household [15]. We make an exception in the case of people who are temporarily staying in institutions (e.g. hospitals, university residences, prisons, military barracks), who are considered to remain part of the household while in the institution. We only consider households in which there is at least one individual aged 15 or older, as households consisting only of younger children are extremely rare (i.e. we assume a household can only be formed by an individual aged 15 or older, and that a household dissolves if there are no remaining members aged 15 or older). In line with most microsimulation models of households [16], we further assume that households consist of individuals who are related by blood or by marriage. In the model, every household is assigned an ID, and every individual is assigned a single household ID, indicating the household in which they spend most nights. An individual who is assigned a household ID of zero is considered homeless.

For the sake of defining relationships between individuals within households, we also assign a ‘head’ to each household, i.e. an individual aged 15 or older who is considered ‘in charge’ of the household. Ziehl [17] notes that the notion of headship is problematic, as there is no clear criterion for determining which household member is head, and there may not be consensus among household members as to which member is the head. We do not attempt a formal definition of headship, but note that in most households the head is the most senior member of the household.

##### 1.2.2 Initial assignment to households in 1985

At the start of the simulation, in 1985, all individuals are assigned to a household. This involves proceeding through three steps:

1. For each individual aged 15 or older at the start of the simulation, randomly assign them to be either a household head or a non-head member.
2. For each individual aged 15 or older who is not a head, find them a family member who they co-reside with.
3. For each child aged younger than 15, assign them to the household of a parent or grandparent.

In each of these three steps, the probabilities of assignment to headship (or relationship with the head) are estimated from multivariable logistic regression models that have been applied to the 1993 October Household Survey (OHS) data. (The 1993 OHS was chosen as it was the earliest nationally representative survey after 1985 for which it was possible to obtain information on household structure.) The sections that follow describe the three steps and the regression models in more detail.

*Assignment of household heads in 1985*

Table S3 summarizes the results of the first logistic regression model, which determines the probability of being household head. The regression model is fitted separately for males and females, as some of the factors have very different effects in men and women. For example, men who are married are substantially more likely to be household heads than men who are unmarried, but the opposite is the case for women, and current employment is much more strongly associated with being head in men than in women. In both sexes, age is strongly positively related to the probability of being head. The results in Table S3 are used in our model to calculate the probability that an adult is a household head. For example, the odds of being a household head in an African man aged 25-29, who is unmarried and unemployed, with incomplete secondary education, is $1.34 \times 0.08 \times 0.63 = 0.067$, and the associated probability of being head is 0.06 (0.067 / (1 + 0.067)). We randomly assign headship based on this calculated probability. If a married/cohabiting individual is assigned to headship, their partner is automatically assigned to the same household (as a non-head member).

Table S3: Predictors of household headship in men and women

| | Males (aOR, 95% CI) | Females (aOR, 95% CI) |
|---|---|---|
| Married/cohabiting | 17.86 (16.63-19.19) | 0.10 (0.09-0.11) |
| Age group | | |
| 15-19 | 0.01 (0.01-0.02) | 0.02 (0.01-0.02) |
| 20-24 | 0.04 (0.04-0.05) | 0.05 (0.04-0.06) |
| 25-29 | 0.08 (0.06-0.09) | 0.11 (0.09-0.12) |
| 30-34 | 0.14 (0.11-0.16) | 0.23 (0.20-0.26) |
| 35-39 | 0.23 (0.20-0.28) | 0.39 (0.35-0.45) |
| 40-44 | 0.35 (0.29-0.42) | 0.57 (0.51-0.65) |
| 45-49 | 0.55 (0.45-0.67) | 0.75 (0.66-0.85) |
| 50-54 | 0.60 (0.48-0.73) | 0.79 (0.69-0.89) |
| 55-59 | 0.76 (0.60-0.96) | 0.87 (0.76-1.00) |
| 60-64 | 0.94 (0.74-1.20) | 1.00 (0.88-1.14) |
| 65+ | 1 | 1 |
| Population group | | |
| African | 1 | 1 |
| ‘Coloured’ | 0.80 (0.73-0.88) | 0.74 (0.68-0.81) |
| White and Asian | 1.20 (1.09-1.32) | 0.54 (0.49-0.58) |
| Currently employed | 3.35 (3.09-3.64) | 1.33 (1.24-1.43) |
| Currently attending school/ higher education | 0.49 (0.39-0.60) | 0.48 (0.39-0.61) |
| Highest educational attainment | | |
| None | 1 | 1 |
| Primary only | 0.74 (0.66-0.83) | 1.02 (0.94-1.11) |
| Incomplete secondary | 0.63 (0.55-0.71) | 0.84 (0.76-0.92) |
| Completed secondary | 0.73 (0.63-0.84) | 0.96 (0.86-1.08) |
| Tertiary | 1.05 (0.84-1.31) | 1.65 (1.35-2.03) |
| Ever given birth | - | 3.12 (2.79-3.48) |
| Constant | 1.34 (1.15-1.55) | 0.73 (0.64-0.84) |

Source: 1993 October Household Survey (author’s own calculations)

*Assignment of adults who are not heads to households*

In the second step, we assign non-head adults (who aren’t married to a household head) to other households. We start by assessing whether these individuals are still living with their parents. This is based on sex-specific logistic regression models that predict the probability of being the child of a household head. Table S4 summarizes the results of the models fitted to the 1993 OHS data. It is very uncommon for a married woman to be living in the household of her parents, but it is relatively common for a married man to be living in the same household as his parents, a pattern consistent with patrilocal kinship systems in South Africa [17, 18]. As might be expected, the probability of living with parents is strongly negatively related to the individual’s age. As before, the results from these logistic regression models are used to determine the probability that an individual lives in the same house as their parent if they are not a head. For example, the odds of being the child of a household head, for a coloured woman aged 30-34 who is unmarried and employed, with incomplete secondary education, is 0.051 × 26.9 × 1.19 × 0.80 × 1.76 = 2.30. The associated probability of being the child of the household head is 0.70 (2.30 / (1 + 2.30)). The individual is randomly assigned to living with their father (if their father is alive and is a household head) or living with their mother (in the event that their father is not alive or is not a household head, and the mother is alive and is a household head).

Table S4: Predictors of being the child of a household head, in non-head adults

| | Males (aOR, 95% CI) | Females (aOR, 95% CI) |
|---|---|---|
| Married/cohabiting | 0.45 (0.40-0.50) | 0.03 (0.02-0.03) |
| Age group | | |
| 15-19 | 28.64 (22.22-36.91) | 44.36 (35.93-54.77) |
| 20-24 | 26.86 (20.98-34.40) | 39.99 (32.58-49.09) |
| 25-29 | 26.16 (20.35-33.63) | 36.35 (29.49-44.81) |
| 30-34 | 22.71 (17.53-29.43) | 26.90 (21.69-33.36) |
| 35-39 | 17.02 (12.97-22.32) | 16.84 (13.43-21.11) |
| 40-44 | 11.28 (8.41-15.12) | 10.41 (8.16-13.28) |
| 45-49 | 6.06 (4.33-8.48) | 7.67 (5.83-10.09) |
| 50+ | 1 | 1 |
| Population group | | |
| African | 1 | 1 |
| 'Coloured' | 0.96 (0.87-1.05) | 1.19 (1.08-1.30) |
| White and Asian | 1.55 (1.40-1.72) | 1.06 (0.97-1.17) |
| Currently employed | 0.87 (0.79-0.95) | 0.80 (0.74-0.86) |
| Currently attending school/ higher education | 1.14 (1.02-1.27) | 1.38 (1.25-1.52) |
| Highest educational attainment | | |
| None | 1 | 1 |
| Primary only | 1.34 (1.15-1.56) | 1.29 (1.13-1.47) |
| Incomplete secondary | 1.60 (1.38-1.86) | 1.76 (1.55-2.00) |
| Completed secondary | 1.91 (1.61-2.26) | 2.05 (1.77-2.37) |
| Tertiary | 1.81 (1.32-2.48) | 1.48 (1.13-1.94) |
| Constant | 0.087 (0.067-0.112) | 0.051 (0.042-0.063) |

Source: 1993 October Household Survey (author's own calculations)

If the individual is not randomly assigned to live with their parent (or if neither parent is a household head), we instead randomly assign them to live with a maternal sibling who is alive and is a household head. If there are no surviving maternal siblings who are household heads, we instead randomly assign them to live with a paternal sibling who is a household head. If there are also no surviving paternal siblings who are household heads, we randomly assign them to live with any children who are aged 25 or older, who are household heads (the age 25 condition is included on the assumption that it would be very unlikely for a parent to be living with a child younger than 25 if the latter was considered the household head). If there are no children who are household heads, we instead assign the individual to live with their surviving mother or (if the mother is no longer alive) surviving father, even though their parent is not a household head (this assignment can only occur if the relevant parent already has been assigned to a household in a non-head capacity). Finally, if the individual has still not been assigned to a household, we re-assign them to be the head of a household.

*Assignment of children to households in 1985*

In the third and final step, we assign each child under the age of 15 to live with a parent or grandparent. Here we use the 1995 OHS rather than the 1993 OHS, as the 1995 survey included questions about whether children's parents were alive and whether children were living with their parents (questions that were not asked in the 1993 or 1994 OHSs). Table S5 shows the predictors of living in the same household as one's mother, if one's mother is alive, as well as the predictors of living in the same household as one's father, if one's father is alive and the child is not living with its mother.

Table S5: Predictors of children living with parents (if parents are alive)

| | Living with mother (aOR, 95% CI) | Living with father, if not with mother (aOR, 95% CI) |
|---|---|---|
| Age group | | |
| 0-4 | 1 | 1 |
| 5-9 | 0.69 (0.63-0.76) | 1.23 (0.93-1.62) |
| 10-14 | 0.62 (0.55-0.70) | 1.37 (0.99-1.90) |
| Race | | |
| African | 1 | 1 |
| 'Coloured' | 1.41 (1.30-1.53) | 0.82 (0.64-1.06) |
| White and Asian | 3.98 (3.49-4.53) | 2.11 (1.57-2.82) |
| Currently in school | 0.99 (0.90-1.09) | 0.85 (0.66-1.10) |
| Constant | 6.48 (6.13-6.86) | 0.13 (0.11-0.16) |

Source: 1995 October Household Survey (author's own calculations)

In our model we first determine if the child is living with their mother (if the mother is alive), and if they are not, determine whether the child is staying with their father (if the father is alive). In the event that the child is not living with a biological father or mother, we assign the child to live with their maternal grandmother, maternal grandfather, paternal grandmother or paternal grandfather (in descending order of preference, depending on which grandparent is alive).

##### 1.2.3 Modelling changes in household membership

The sections that follow describe the procedures that are followed when modelling different types of event *after* the start of the simulation in 1985. In this section, we rely on data from the General Household Surveys (GHSs), conducted between 2002 and 2018, to determine realistic assumptions about movements between households.

*Birth of a child or movement of a child out of its current household*

Similar to the procedure described in the previous section, we calculate the probability that a child is living with its mother (if the mother is alive) and the probability that the child is living with its father (if the father is alive and the child is not living with its mother). Table S6 summarizes the results of the multivariable logistic regressions applied to the 2002, 2006, 2010, 2014 and 2018 GHSs. Results are similar to those in Table S5: although the age effect is modelled differently, there is the same trend towards lower odds of living with one's mother in older children, and the inter-racial differences are similar to those estimated previously. We use the average of the odds ratios estimated from the five GHSs (final column) in setting the model assumptions. However, it is important to note that the logistic regression assesses the cross-sectional association between age and household membership, whereas our model assesses *change* in household membership. For example, the negative association between age and living with one's mother could be due to the higher cumulative likelihood of the mother having moved out of the household in which the child was born; it does not necessarily mean that a mother who moves out of a household is less likely to take her child with her if the child is older than if the child is younger. We therefore do not include an age effect in our model. For example, the odds of an African child being assigned to the same household as their mother immediately after birth is 7.88, and the associated probability is 0.89 (7.88 / (1 + 7.88)); if the mother subsequently moves out of the household in which the child was born, the same probability of 0.89 is used to determine whether the child moves with their mother. This is consistent with high rates of circulation of children between households in African kinship networks [15, 18].

Table S6: Predictors of children living with parents (if parents are alive)

| | 2002 | 2006 | 2010 | 2014 | 2018 | Average |
|---|---|---|---|---|---|---|
| *Odds of living with mother* | | | | | | |
| Age | 0.73 | 0.79 | 0.77 | 0.76 | 0.79 | 0.77 |
| Age squared | 1.015 | 1.011 | 1.014 | 1.014 | 1.012 | 1.013 |
| Race | | | | | | |
| African | 1 | 1 | 1 | 1 | 1 | 1 |
| ‘Coloured’ | 2.19 | 1.88 | 2.05 | 2.65 | 2.12 | 2.18 |
| White and Asian | 9.12 | 7.39 | 4.62 | 6.98 | 5.04 | 6.63 |
| Currently in school | 1.21 | 1.00 | 1.22 | 1.19 | 1.03 | 1.13 |
| Constant | 11.17 | 8.14 | 6.53 | 6.49 | 7.09 | 7.88 |
| *Odds of living with father, if child is not with mother* | | | | | | |
| Age | 0.99 | 0.97 | 0.94 | 0.93 | 0.94 | 0.95 |
| Age squared | 1.003 | 1.003 | 1.007 | 1.008 | 1.006 | 1.005 |
| Race | | | | | | |
| African | 1 | 1 | 1 | 1 | 1 | 1 |
| ‘Coloured’ | 1.29 | 1.25 | 1.71 | 2.42 | 2.54 | 1.84 |
| White and Asian | 4.53 | 2.38 | 4.54 | 8.93 | 8.86 | 5.85 |
| Currently in school | 0.87 | 0.86 | 0.91 | 0.88 | 0.95 | 0.90 |
| Constant | 0.15 | 0.17 | 0.16 | 0.16 | 0.16 | 0.16 |

Source: General Household Surveys (author’s own calculations). Results presented are adjusted odds ratios (95% confidence intervals not shown).

If the child is not living with their biological mother or father, we determine whether they are living with a grandparent. Preference is given to grandparents who are household heads (i.e. a child is less likely to be assigned to the household of a grandparent who is not a head), and after accounting for headship, preference is given to maternal grandmothers, maternal grandfathers, paternal grandmothers and paternal grandfathers (in that order). Only if there is no surviving mother or grandparent do we assign the child to the household of an aunt or uncle. Finally, in the event that there is also no surviving aunt or uncle, we assume the child enters an orphanage, and is classified as being part of the ‘homeless’ population.

*Death of a household head*

In the event that the household head dies, one of two things can happen. Firstly, if there are other household members who are aged 15 or older, headship is assumed to switch to the individual with the highest odds of being a household head (where the odds are calculated using the same coefficients that are used to assign headship at baseline, i.e. the coefficients in Table S1). Alternatively, if the head was the only individual in the household aged 15 or older, it is assumed that the household dissolves and any remaining children move to the households of other relatives, using the same algorithm as described in the previous section (i.e. first testing to see if the child stays with their mother, then testing to see if the child stays with their father, etc.).

*Household changes associated with marriage/cohabitation*

Most microsimulation models of household formation assume that when a couple marries or starts cohabiting, they establish a new household [16]. Although this assumption is reasonable in a Western setting, where households tend to be nuclear in structure, the assumption may be less appropriate in an African setting, where it is common for men to remain in their parents' household after marriage and for women to join the households of their husbands [17, 18]. Table S7 shows an analysis of 2018 GHS data on the living arrangements of married men aged 20-49. Multivariable logistic regression was used to assess the factors associated with living with one's parents (in a non-head capacity) if one's parents are still alive. The results imply, contrary to suggestions from the literature, that co-residence of married men with their parents is not an exclusively African household arrangement, and that in fact the odds of co-residence are slightly higher for non-African races than for Africans. Secondly, although there is a high probability of co-residence with parents among married men who are young and unemployed, this probability reduces rapidly as men age and enter into employment. For example, the odds of co-residence with parents in married African men aged 25 who are unemployed and living in urban areas is 0.39 ($13.1 \times 0.909^{25} \times 0.324$), but this reduces to 0.016 in married African men aged 45 who are employed and living in urban areas ($13.1 \times 0.909^{45} \times 0.324 \times 0.284$). Considering that rates of marriage are low among young men and men who are unemployed, the latter is likely to be more representative. Although newly married couples may initially live with their parents, these arrangements appear temporary in most circumstances, in contrast to the permanent attachment to paternal households described in the literature. Similar results are obtained when the logistic regression model is repeated with data from the 2010 GHS (final column of Table S7).

Table S7: Predictors of married men living with their parents in a non-head capacity, if parents are still alive

| | 2018 | 2010 |
|---|---|---|
| Per year increase in age | 0.909 (0.893-0.925) | 0.900 (0.888-0.913) |
| Race | | |
| African | 1 | 1 |
| 'Coloured' | 1.26 (0.83-1.91) | 1.16 (0.86-1.56) |
| White and Asian | 2.10 (1.48-2.97) | 1.61 (1.20-2.15) |
| Employment | 0.28 (0.22-0.37) | 0.31 (0.26-0.39) |
| Urban location | 0.32 (0.25-0.42) | 0.33 (0.27-0.40) |
| Constant | 13.13 (6.97-24.74) | 20.48 (12.38-33.86) |

Source: General Household Surveys (author's own calculations). Results presented are adjusted odds ratios (with 95% confidence intervals in brackets). Analysis is limited to married/cohabiting males aged 20-49, with one or both parents still alive.

In the interests of simplicity, we assume that newly married/cohabiting couples form a new household unless either is already a household head (in which case the non-head partner joins the household of the head partner). In the event that both partners are already heads, or neither partner is a head, we assign headship to whichever partner has the highest headship probability (calculated from the regression coefficients in Table S3). Any biological children under the age of 18 who were previously living with either partner get assigned to the new household or a different household, using the same algorithm as described before (i.e. using the coefficients from Table S6).

*Household changes associated with divorce*

We assume that in the event of a divorce (or dissolution of a cohabiting union), the partner who is not the household head leaves the household (or in the rare situations where neither partner is head, we assume that the partner who is not related to the household head leaves the household). If the male partner is the partner who leaves the household, we assume that he establishes a new household. If the female partner is the partner who leaves the household, we calculate her probability of establishing a new household based on a logistic regression model fitted to data from the 2018 GHS (Table S8). As might be expected, divorced/separated women are more likely to be heads if they are older and are employed. Even after controlling for age and other variables, divorced women whose parents are alive are less likely to be household heads, suggesting that such women are likely to return to their parent's home after a divorce.

Table S8: Predictors of household headship in women who are divorced/separated

| | Adjusted odds ratio (95% CI) |
|---|---|
| Per year increase in age | 1.25 (1.17-1.32) |
| Per unit increase in age squared | 0.9983 (0.9977-0.9989) |
| One or both parents alive | 0.34 (0.24-0.49) |
| Race | |
| African | 1 |
| 'Coloured' | 0.69 (0.46-1.03) |
| White and Asian | 0.45 (0.3-0.67) |
| Employment | 2.37 (1.75-3.22) |
| Constant | 0.0076 (0.0017-0.0346) |

Source: 2018 General Household Survey (author's own calculations). Urban location was omitted as it was not significant.

We assume that women who do not form new households after leaving the household in which they were cohabiting move to the household of a relative in the same urban/rural location. Preference is given to the household of a parent, a sibling, a child, or a grandparent (in that order).

*Household changes associated with urban-rural migration*

Although migration is often a household event (the whole household moves), we model migration between urban and rural areas as being (primarily) an individual-level event. Only in the event that a woman moves, and she has children who were previously living with her, do we allow for the possibility that the children move with their mother. All other household members are assumed to remain (at least initially) in the household from which the departing member migrated. If the individual was temporarily separated from their partner (e.g. a wife living in rural area and her husband living in urban area), they are automatically assumed to move into the same household as their partner when they migrate, if that partner is a household head. Otherwise we assume that an individual who migrates has a certain probability of forming a new household when they migrate to their new destination, and we use this probability to randomly assign them to either a new household or an existing household. Those who do not establish a new household are assumed to join the household of a relative, with preference being given to the household of a spouse, a parent, a sibling, a child, or a grandparent (in that order).

In order to determine the probability of forming a new household, we apply a logistic regression model to 2011 census data, which aims to assess the factors associated with establishing a new household versus joining an existing household, in individuals who have recently migrated (Table S9). The analysis is limited to individuals aged 15 and older who reported having moved from another municipality into the current municipality in 2011 (i.e. less than 12 months ago). It is difficult to use the census data to determine whether individuals moved from an urban area to a rural area (or vice versa), but we use 'moving between municipalities' as a rough proxy for migration between urban and rural areas. The outcome in the logistic regression is being a household head, which is a reasonable proxy for having established the present household (rather than having joined an existing household). As might be expected, the factors associated most strongly with establishing a new household are older age and employment. Married men have a higher probability of forming a new household than unmarried men, but married women are less likely than unmarried women to form a new household.

Table S9: Predictors of being a household head among recent migrants

| | Males (aOR, 95% CI) | Females (aOR, 95% CI) |
|---|---|---|
| Married/cohabiting | 2.45 (2.36-2.55) | 0.17 (0.16-0.17) |
| Age group | | |
| 15-19 | 1 | 1 |
| 20-24 | 2.66 (2.47-2.86) | 2.51 (2.33-2.70) |
| 25-29 | 4.33 (4.01-4.68) | 3.82 (3.52-4.13) |
| 30-34 | 5.73 (5.28-6.23) | 5.31 (4.88-5.79) |
| 35-39 | 6.50 (5.94-7.12) | 6.94 (6.33-7.61) |
| 40-44 | 6.62 (5.98-7.33) | 8.25 (7.45-9.13) |
| 45+ | 7.33 (6.70-8.02) | 7.50 (6.87-8.18) |
| Population group | | |
| African | 1 | 1 |
| 'Coloured' | 0.47 (0.44-0.51) | 0.52 (0.49-0.57) |
| White and Asian | 0.63 (0.6-0.66) | 0.51 (0.48-0.53) |
| Currently employed | 2.17 (2.09-2.25) | 2.04 (1.96-2.12) |
| Currently attending school/ higher education | 1.32 (1.25-1.40) | 1.50 (1.43-1.58) |
| Highest educational attainment | | |
| None | 1 | 1 |
| Primary only | 0.87 (0.80-0.95) | 0.97 (0.89-1.07) |
| Incomplete secondary | 0.91 (0.84-0.97) | 0.95 (0.88-1.02) |
| Completed secondary | 1.10 (1.02-1.18) | 1.10 (1.02-1.18) |
| Tertiary | 1.61 (1.47-1.76) | 1.39 (1.28-1.52) |
| Urban location | 0.83 (0.79-0.87) | 0.93 (0.88-0.98) |
| Constant | 0.22 (0.20-0.25) | 0.22 (0.20-0.24) |

Source: 2011 census (author's own calculations)

*'Leaving the nest' due to employment and tertiary education*

In the previous sections we allowed for adults to 'leave the nest' if they became orphaned, got married or moved between urban and rural areas. However, adults may eventually leave home for other reasons, typically because they have become financially independent or need to move for the purposes of tertiary education.

Table S10 shows the results of two multivariable logistic regression models fitted to data from the 2018 General Household Survey. In both regressions we include only data for the 15-49 age group (the age group in which individuals are most likely to 'leave the nest'), and include only those who report never being married/cohabiting with a partner (to exclude the effect of household movement associated with marriage, since that is accounted for separately in our model). We also include only those who report that their parents are still alive (to exclude the effect of household movements associated with orphanhood, which are also accounted for separately in our model). The first regression assesses factors associated with having left home (defined here as not being the child/stepchild or grandchild of the household head). As might be expected, this is strongly associated with age and employment status. There is also a higher probability of having left home among individuals who are currently studying (controlling for age), suggesting that tertiary education is often associated with leaving home.

Table S10: Factors associated with having left home, and with being a household head among those who have left home

| | Odds ratio for having left home (aOR, 95% CI) | Odds ratio for being household head (aOR, 95% CI) |
|---|---|---|
| Female sex | 0.85 (0.80-0.91) | 1.19 (1.06-1.34) |
| Per year increase in age | 1.15 (1.12-1.19) | 1.37 (1.30-1.46) |
| Per unit increase in age squared | 0.9986 (0.9981-0.9991) | 0.9965 (0.9956-0.9974) |
| Population group | | |
| African | 1 | 1 |
| 'Coloured' | 0.38 (0.33-0.44) | 0.35 (0.26-0.46) |
| White and Asian | 0.48 (0.41-0.57) | 0.63 (0.45-0.87) |
| Currently attending school/ higher education | 1.33 (1.20-1.48) | 1.71 (1.41-2.09) |
| Currently employed | 2.89 (2.68-3.11) | 3.03 (2.67-3.45) |
| Constant | 0.023 (0.014-0.037) | 0.0016 (0.0006-0.0040) |

Source: 2018 General Household Survey (author's own calculations). Analysis is limited to never-married adults aged 15-49 who have at least one surviving parent (and second analysis is further limited to those who have left home).

The second regression model is further limited to individuals who have left home (i.e. who are not reported to be children or grandchildren of the household head), and assesses factors associated with being head. As with the first model, the factors associated with being a head are older age, employment and tertiary education.

In MicroCOSM, we use the results from the first regression model to calculate age-specific probabilities of having left home due to employment or tertiary education, and then calculate the annual probability of leaving home from the difference in age-specific cumulative probabilities of having left home. In other words, if $S_0(x)$ is the probability of having left home for an individual aged $x$ in the baseline covariate group, and $p_0(x)$ is the annual probability of leaving home, for an individual aged $x$ in the baseline covariate group, we calculate

$$p_0(x) = (S_0(x + 1) - S_0(x)) / (1 - S_0(x)).$$

We define the baseline covariate group as black males who are employed and not studying. For the purpose of calculating the baseline covariate probability $S_0(x)$ we also reduce the constant base odds (0.023) to 0.005; this is because the regression model does not account for movements out of the household that are related to urban-rural migration, even though these are accounted for separately in MicroCOSM. Thus, for example, the baseline odds of having left home at age 30 is $0.005 \times 1.15^{30} \times 0.9986^{30\times30} \times 2.89 = 0.27$, which means $S_0(30) = 0.21$ (0.27/1.27).

We then multiply the $p_0(x)$ values by the odds ratios in Table S10 to account for the effects of sex, race and current studying. Although it is not strictly correct to use odds ratios as if they are risk ratios, the relation is approximately correct because the annual probabilities are low. We apply the same probabilities to married and unmarried individuals, which may also introduce bias (although it should be noted that the model also makes provision for a once-off probability of forming a new household at the time marriage occurs, as described previously). We also multiply the annual probability of leaving by an arbitrary factor of 2 if there are 10 or more individuals in the parental household (to take into account that people living in overcrowded households might feel more pressure to leave the nest).

If the individual leaves the home of their parent/grandparent in a given year, we calculate the probability that they form a new household from the second regression model in Table S10. If they do not form a new household, we assume they instead join the household of a sibling, assuming there is a sibling living in the same urban/rural area.

*Returning to the nest if unemployed*

We assume that if an unmarried individual loses their job (becomes unemployed), and is aged less than 40, there is a possibility that they return to the home of a parent. We assume that this only occurs if the parent is alive and is either a household head or is the spouse of a household head. We also assume that the return to the nest only occurs if parent and child both currently live in urban areas or both currently live in rural areas.

##### 1.2.4 Homelessness

We define individuals as homeless if they are living on the streets, with no permanent home. This differs from the broader definition of homelessness used in some studies, which include individuals living in informal settlements, individuals in temporary housing and individuals whose homes are insecure [19]. A major challenge in setting assumptions about homelessness is that there are no nationally representative data on homelessness. Although the censuses are supposed to collect data on homelessness, implausible discrepancies between successive censuses have been noted [20], and we are therefore forced to rely on limited local surveys.

The size of the homeless population is particularly difficult to estimate. A survey conducted by the HSRC in 2007 suggested a national homeless population of 100 000 to 200 000, although the methods used to obtain this estimate were not clearly described [20, 21]. This is equivalent to 0.2-0.4% of the South African population at the time. A census of the homeless population in Durban estimated the number of homeless people to be around 4 000 [22], equivalent to 0.13% of the Durban population at the time. This could be an under-estimate, as the census only covered the central business district and immediate surrounds (although it has been noted that homeless individuals tend to live in inner-city and surrounding neighbourhoods [21]). An earlier census of homeless people in the greater Cape Town area estimated the homeless population to be 4 133 in 1999 [23], equivalent to 0.15% of the population at the time. Thus most estimates of the homeless population are around 0.2% of the corresponding total population, with a range of 0.1-0.4%.

Homeless populations are predominantly male. Studies in Cape Town have estimated the male fraction to be around 60-75% [23-25], but in the rest of the country the proportion is much higher, typically about 85% [20, 22]. Homeless adults are mostly young, with the 25-34 age group being the age group that accounts for the largest proportion of homeless adults, and with median ages of around 30-35 years [20, 22, 26]. However, the relative size of the homeless child population is difficult to determine, as most studies sample only adults or only children. The only two studies that included adults and children in the same sampling frame estimated the fraction of homeless individuals who were children to be 2% in Durban [22] and 19% in Cape Town [23]. Durations of homelessness are variable; in a survey of homeless adults in four provinces in northern South Africa, the average duration of homelessness was 6.4 years [20], while median durations of 3 years and more than 5 years have been estimated in Durban [22] and Cape Town [23] respectively.

The most commonly reported reasons for homelessness include unemployment, substance use, crime/incarceration, divorce, abuse, death of a parent/family member, mental health and disability [22-25, 27, 28]. We attempt to allow for some of these factors by allowing for the possibility that people become homeless at various points in their life course: on migrating between urban and rural areas (if they cannot find a relative to live with and lack the means to pay for their own accommodation), on divorce/separation, on release from prison, and on death of a parent/guardian (in the case of children). We also allow for people to leave the homeless population and either establish their own household or join an existing household, with the rate of leaving the homeless population depending on their sex and employment status.

As described in previous sections, when individuals leave their home due to divorce/separation or urban-rural migration, an algorithm is used to determine whether they establish a new household or join the household of a relative. If neither event is assigned, and the individual is unemployed, we assume they become homeless with probability $h$. The same probability of homelessness is applied to men on release from prison, if they are unemployed. The value of $h$ has been set to 0.1 in order to produce a homeless adult population that is approximately 0.2% of the total adult population. Those individuals who are not assigned to homelessness instead form a new household.

At the start of each year we test whether homeless individuals return to households. The odds of return is $r_{g,i}$ for people of sex $g$ (0 for males, 1 for females) and employment status $i$ (0 for unemployed, 1 for employed). The base odds (for unemployed males) is set to 0.15, this value being chosen so that the average duration of homelessness is approximately equal to 6 years. The base odds is multiplied by a factor of 2 for females, this value being chosen such that approximately 20% of the homeless population is female. Similarly the base odds is multiplied by a factor of 1.6 for employed individuals, this value being chosen such that approximately 27% of homeless adults are employed [20].

Although individuals in institutions remain classified as being in households while they are (temporarily) institutionalized, the same is not true for children who are in orphanages. This means that some children are classified as ‘homeless’ in the model even though they are actually in orphanages. It is difficult to estimate the size of the population in orphanages, as no official estimates have been published, but according to the 2011 census 60 000 children under the age of 15 were in institutions (approximately 0.4% of the child population). We would therefore expect the modelled proportion of children who are homeless or institutionalized to be at least 0.4%.

##### 1.2.5 Model calibration and validation

Figure S3a compares the modelled proportion of household members who are household heads with the proportions reported to be household heads in the 2018 GHS. Although the modelled proportions appear slightly too high relative to the data in the 15-29 age group, the overall age pattern is roughly consistent with the data. Similarly Figures S3b-d compare the modelled and reported proportions of household members who are spouses of a household head, children of a household head and grandchildren of a household head. The modelled proportions who are partners of a household head are slightly higher than the reported proportions over the 40-64 age range, but again the overall pattern is consistent with the data (Figure S3b). The model under-estimates the proportion of individuals aged 10-29 who are children of household heads (Figure S3c) and under-estimates the proportion of children living with grandparents (Figure S3d). Although attempts were made to improve the model fit to the data in Figures S3c and S3d, it proved difficult to accomplish this. It is possible that the reported proportions of children/youth who are children/grandchildren of the household heads might be over-estimates if caregivers who are not biological parents/grandparents of the child in question tend to report the child as their own [29].

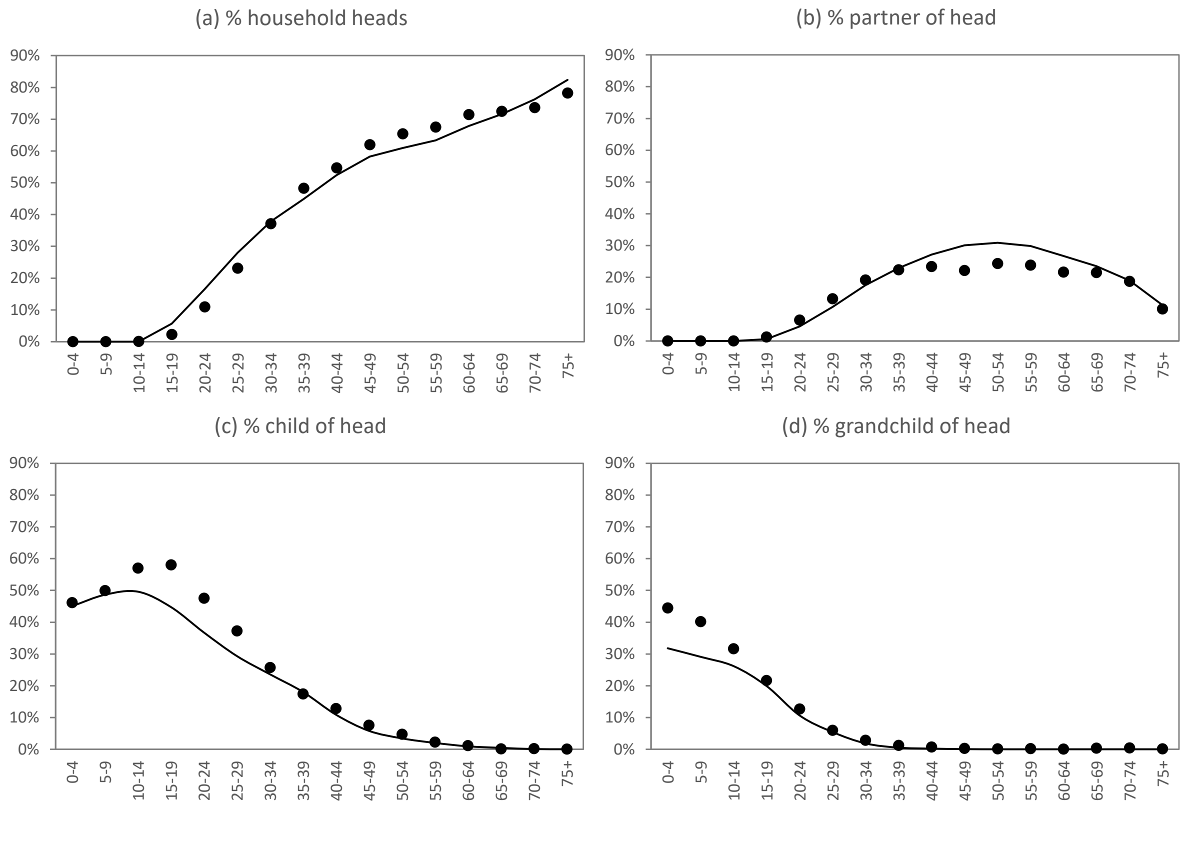

Figure S3: Proportion of household members with different relationships to the household head in 2018
Data are from the 2018 General Household Survey.

Figure S4 shows the modelled distribution of household sizes in three years (1998, 2008 and 2018) compared against the observed distribution in nationally representative household surveys. Although the modelled distribution is roughly consistent with the distributions in the household surveys, the model consistently under-estimates the proportion of the population in households of 5-6 individuals and over-estimates the proportion in households of 10 or more individuals.

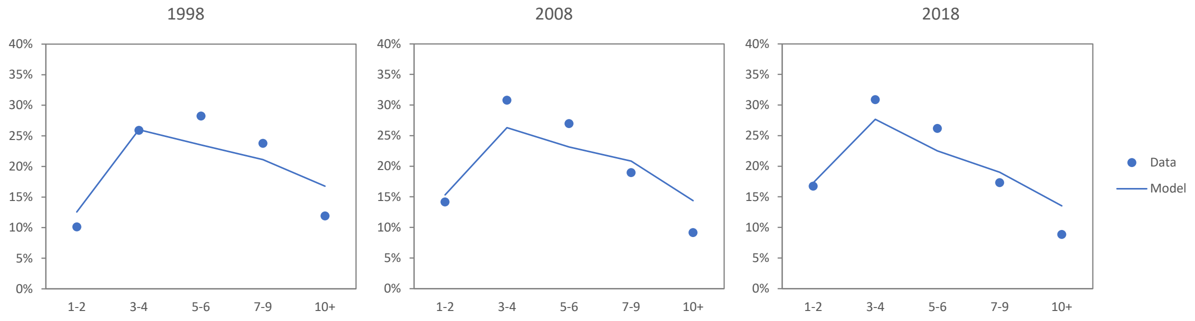

Figure S4: Proportion of household members in households of different size
Data are from the 1998 October Household Survey and the 2008 and 2018 General Household Surveys.

Table S11 compares the modelled characteristics of the homeless population and the calibration targets described previously. The model results are roughly consistent with the available survey data for the homeless population. Although we have not made any assumptions about the effect of alcohol consumption on either the rate of entry into homelessness or the rate of leaving the homeless population, the modelled level of drinking is roughly consistent with the level that would be expected based on reported levels of drinking in surveys of the homeless population.

Table S11: Characteristics of the homeless population

| | Model average (2010-18) | Target (range) |
|---|---|---|
| % of adult population (18+) that is homeless | 0.19% | 0.20% (0.1-0.4%) |
| % of child population that is homeless or in institutions | 0.59% | >0.4% |
| % of homeless adults who are female | 22% | 20% (15-40%) |
| % of homeless adults who are employed | 24% | 27% |
| Average duration of homelessness (adults) | 6.2 years | 6 years |
| % of homeless adults who drink more than two times per week | 29% | 33%* |

* The 33% is based on a survey of homeless adults in four provinces, of whom 21.4% reported drinking at least 2 times per week [24], adjusting for an assumed 35% under-reporting of the frequency of drinking (see [30]).

##### 1.2.6 Limitations

A disadvantage of using the 1993 OHS data in assigning the initial distribution of the population across households is that this survey was done 8 years after the start of our simulation, in 1985. The survey might not realistically reflect the actual household structures in 1985, particularly given the scrapping of the pass laws in 1986. Another disadvantage of using the 1993 OHS is that, unlike most later surveys, it did not include questions on whether individuals' parents were alive, which thus makes it difficult to determine whether people who are living without their parents are doing so because their parents are no longer alive or because they have moved out of their parent's home. The 1993 OHS also differs from later surveys in that it included domestic workers as household members; in the censuses and surveys after 1994, domestic workers who lived on the same property as their employer were instead considered separate households [17].

Our model does not account for all the factors that may cause household dissolution or reconfiguration. In South Africa, an important factor in the contexts of urban informal settlements is shack fires, which frequently cause the destruction of homes [15].

Another limitation is that our model only considers the formation of households that are defined by kinship (i.e. people who are related by blood or marriage). Although this may mean excluding certain household arrangements (e.g. roommates sharing a flat, or individuals temporarily living with friends), such arrangements are relatively uncommon. In the 1995-1999 OHSs and the 2002-2018 GHSs, the average proportion of household members who were reported as being unrelated to the household head was only 0.9%. This suggests that any bias from excluding such individuals from households defined by kinship is likely to be small.

Further refinement of the model is required in order to achieve better consistency with household survey data. In particular, confidence in the model would be improved if we could increase the proportion of children who are either children or grandchildren of household heads, and if we could reduce the proportion of the population of the population living in large households (10 or more individuals).

#### 1.3 Modelling household income

For the purpose of evaluating an individual's socio-economic status, it is generally preferable to consider household per capita income rather than individual income, as the former reflects the level of resource sharing that typically takes place within a household [31]. Household per capita income is also the basis for most metrics of inequality [14]. Household per capita income can be calculated in different ways. The simplest approach, which we use for the purpose of comparing to estimates published by Statistics South Africa [14], is to divide the total income of the household by the number of members of the household. However, this might not reflect the true level of financial distress experienced by a household, as children generally cost less to support than adults, and there are economies of scale in meeting the needs of larger households compared to single-person households. For the purpose of measuring the effect of financial distress on sexual behaviour, we therefore use an 'equivalence scale' that has been widely used in South Africa [32], when calculating the denominator in the adjusted per capita household income (as described in the main text).

In South Africa around 90% of all household income is from wages/salaries and social grants; the balance comes from remittances and other income (e.g. from investment income and private pensions) [33]. Of the social grants, the two most significant are the old age pension and the child support grant (CSG), accounting for 44% and 37% respectively of the total grant expenditure in 2019-20 [34]. Although private pension coverage is only about a third of the state old age pension coverage (in the population aged 60 and older), the average private pension is around five times the amount of the state old age pension [35], and private pensions are thus likely to contribute as much as (if not more than) the state old age pension to total income.

In MicroCOSM we model household income as the sum of four sources of income: salary/wage income, CSG payments, state old age pensions and private pensions. The sections that follow describe our approach to modelling each of these.

##### 1.3.1 Salary/wage income

The monthly salary/wage income for each employed individual is modelled based on regression models fitted to data from the General Household Surveys in 2002, 2006, 2010, 2014 and 2018 (Table S12). Individuals who did not report an income are excluded from the regression models, and only positive values were reported. In each survey the monthly income is log-transformed in order to reduce the influence of outliers [31] and to ensure that only positive incomes can be predicted by the model. The results show that higher incomes are associated with male sex, higher educational attainment, older age and white/Asian race, although it appears that the age and sex differentials in earnings have declined over time. These are consistent with declining sex differentials in employment levels over time [10].

Table S12: Factors affecting monthly income in employed South Africans (on a natural logarithm scale)

| | 2002 | 2006 | 2010 | 2014 | 2018 |
|---|---|---|---|---|---|
| Educational attainment (ref. none) | | | | | |
| Primary only | 0.33 | 0.25 | 0.27 | 0.18 | 0.21 |
| Incomplete secondary | 0.79 | 0.67 | 0.69 | 0.57 | 0.50 |
| Completed secondary | 1.44 | 1.30 | 1.40 | 1.21 | 0.99 |
| Tertiary | 2.17 | 2.20 | 2.27 | 2.23 | 1.94 |
| Female sex (ref. male) | -0.53 | -0.46 | -0.41 | -0.41 | -0.42 |
| Race (ref. black African) | | | | | |
| ‘Coloured’ | 0.27 | 0.23 | 0.30 | 0.22 | 0.37 |
| White/Asian | 0.89 | 0.77 | 0.80 | 0.83 | 0.62 |
| Age group (ref. <25) | | | | | |
| 25-29 | 0.26 | 0.22 | 0.22 | 0.20 | 0.11 |
| 30-34 | 0.52 | 0.35 | 0.34 | 0.32 | 0.27 |
| 35-39 | 0.64 | 0.50 | 0.41 | 0.33 | 0.32 |
| 40-44 | 0.77 | 0.63 | 0.48 | 0.39 | 0.36 |
| 45-49 | 0.84 | 0.64 | 0.59 | 0.46 | 0.33 |
| 50-54 | 0.78 | 0.67 | 0.64 | 0.53 | 0.33 |
| 55-59 | 0.71 | 0.64 | 0.70 | 0.65 | 0.38 |
| 60+ | 0.54 | 0.52 | 0.55 | 0.41 | 0.27 |
| Constant | 5.74 | 6.21 | 6.46 | 6.90 | 7.55 |
| Standard deviation of unexplained variance | 0.83 | 0.80 | 0.89 | 1.07 | 1.01 |

Source: General Household Surveys (author’s own calculations)

In our model we also allow for variation in income between employed individuals, even after accounting for the effect of age, sex, race and educational attainment. We do this by estimating the unexplained variance in each regression model and calculating the standard deviation. The results in Table S12 show that this standard deviation is consistently between 0.80 and 1.07. We therefore assume a standard deviation of 0.92 (the average across the five surveys), and randomly assign to each individual an income deviation score by sampling from the normal (0, $0.92^2$) distribution. For example, an employed African male aged 30, who has incomplete high school education, and who has been assigned an income deviation score of 1.00, would have a modelled income in 2018 of exp(7.55 + 0.50 + 0.27 + 1.00) = R11 159 per month. (Examples for other individual characteristics, assuming an income deviation score of 0, are summarized in Table S13.) The income deviation score that is assigned to each individual is assumed to be fixed over time, although it only applies in the period when the individual is employed.

Monthly incomes for each of the years 2002, 2006, 2010, 2014 and 2018 are calculated directly from the coefficients in Table S12. For the other years over the 2002-2018 period, we use linear interpolation to calculate the modelled monthly incomes. In the period before 2002, we estimate monthly incomes from the regression model for 2002, applying a Consumer Price Index (CPI) adjustment and assuming a 2% per annum real growth in incomes. Similarly, in the period after 2018 we estimate monthly incomes using the regression model for 2018, applying a 2% annual real growth rate and an annual inflationary increase of 5.5% (i.e. a total increase in earnings of 7.5% per annum). The 5.5% assumed inflationary increase is based on the average growth in CPI over the 2000-2020 period, and the 2% is based on average real growth in earnings over the 2002-2018 period (Table S13).

Table S13: Examples of modelled average monthly income levels for employed individuals with different characteristics (in South African rands)

| | 2002 | 2006 | 2010 | 2014 | 2018 |
|---|---|---|---|---|---|
| Expected average income | | | | | |
| African men with incomplete 2ndary education, age 30 | 1142 | 1386 | 1789 | 2425 | 4120 |
| African women with complete 2ndary education, age 40 | 1670 | 2156 | 2793 | 3303 | 4801 |
| Coloured men with primary education, age 55 | 1149 | 1544 | 2291 | 2861 | 4963 |
| White men with tertiary education, age 50 | 14406 | 18998 | 27074 | 36327 | 34389 |
| Average annual growth in earnings over the last 4 years | | | | | |
| African men with incomplete 2ndary education, age 30 | - | 4.9% | 6.6% | 7.9% | 14.2% |
| African women with complete 2ndary education, age 40 | - | 6.6% | 6.7% | 4.3% | 9.8% |
| Coloured men with primary education, age 55 | - | 7.7% | 10.4% | 5.7% | 14.8% |
| White men with tertiary education, age 50 | - | 7.2% | 8.2% | 8.6% | -1.4% |
| Average annual growth across examples | - | 6.6% | 8.0% | 6.6% | 9.3% |
| Annual CPI growth over the last 4 years | - | 3.7% | 7.3% | 5.7% | 5.2% |
| Annual real earnings growth over the last 4 years | - | 2.9% | 0.7% | 0.9% | 4.1% |

Evidence suggests that salary is strongly associated with an individual's conscientiousness, as well as other personality traits [36, 37], although South African data are unfortunately lacking. We assume that for every standard deviation increase in conscientiousness (relative to the population average) the income deviation score mentioned previously increases by a factor of 0.13, in line with data from a large nationally-representative US study [36]. This assumption is also consistent with a meta-analysis of longitudinal studies, which found the correlation between conscientiousness scores and income to be 0.14 [37].

##### 1.3.2 Child support grants

The CSG was introduced in 1998, and the conditions for eligibility have changed over time. At first the grant was only provided to caregivers of children under the age of 7 years, but over time the age limit increased to 18 years. In addition there has been a means test, which has changed over time: in the period before 2008 the means test referred to the household income, but thereafter the means test referred to the individual caregiver's income (or the couple's income if the caregiver was married, with the means test threshold for couples being twice that for single caregivers). In our model we assume, for the purpose of determining CSG eligibility, that from 2008 onward the caregiver is the biological mother, or if she is not present in the household, the youngest woman (15 or older) in the household, or if there is no woman in the household, the youngest man in the household. Table S14 shows the assumed annual amount of the CSG, the age threshold for eligibility and the means test for a single caregiver (note that we assume an age of eligibility less than the actual age of eligibility in the first 4 years of the CSG introduction, in order to approximate the effect of low CSG uptake when it was initially introduced [33]). We assume that in the period after 2021 both the CSG amount and the means test threshold for the CSG increase by 5.6% per annum, the average annual growth in the amount of the CSG over the 2016-2021 period.

Table S14: Assumptions regarding grant amounts and eligibility criteria

| Year | Child support grant | | | Old age pension |
|---|---|---|---|---|
| | Monthly amount | Age of eligibility | Means test (monthly income) | Monthly amount |
| 1985-86 | - | - | - | 213* |
| 1986-87 | - | - | - | 226* |
| 1987-88 | - | - | - | 239* |
| 1988-89 | - | - | - | 253* |
| 1989-90 | - | - | - | 269* |
| 1990-91 | - | - | - | 285* |
| 1991-92 | - | - | - | 302* |
| 1992-93 | - | - | - | 320 |
| 1993-94 | - | - | - | 370 |
| 1994-95 | - | - | - | 390 |
| 1995-96 | - | - | - | 410 |
| 1996-97 | - | - | - | 430 |
| 1997-98 | - | - | - | 470 |
| 1998-99 | 100 | -‡ | 1000† | 500 |
| 1999-00 | 100 | -‡ | 1000† | 520 |
| 2000-01 | 100 | -‡ | 1000† | 540 |
| 2001-02 | 110 | <4‡ | 1000† | 580 |
| 2002-03 | 140 | <7 | 1000† | 640 |
| 2003-04 | 160 | <9 | 1000† | 700 |
| 2004-05 | 170 | <11 | 1000† | 740 |
| 2005-06 | 180 | <14 | 1000† | 780 |
| 2006-07 | 190 | <14 | 1000† | 820 |
| 2007-08 | 200 | <14 | 1000† | 870 |
| 2008-09 | 210 | <14 | 2300 | 950 |
| 2009-10 | 240 | <15 | 2400 | 1010 |
| 2010-11 | 250 | <16 | 2500 | 1080 |
| 2011-12 | 260 | <17 | 2600 | 1140* |
| 2012-13 | 280 | <18 | 2800 | 1200 |
| 2013-14 | 290 | <18 | 2900 | 1270 |
| 2014-15 | 310 | <18 | 3100 | 1380* |
| 2015-16 | 320 | <18 | 3200 | 1500 |
| 2016-17 | 350* | <18 | 3300* | 1550* |
| 2017-18 | 380 | <18 | 3400* | 1600 |
| 2018-19 | 405 | <18 | 3600* | 1690* |
| 2019-20 | 425 | <18 | 3800* | 1780 |
| 2020-21 | 450 | <18 | 4000 | 1860 |
| 2021-22 | 460 | <18 | 4200* | 1890 |

All amounts (except for age limits) are in South African rands. Means tests are specified for single individuals; for married individuals the means test is two times the specified threshold and applies to the couple's combined income (except in the case of the CSG in the period before 2008). * Missing data – the assumed value is based on amounts in other years and trends in rand amounts. † The actual means test was specified separately for rural households (R800) and urban households (R1100), but we assume the same amount for all households in the interests of simplicity. Data are from a variety of published [12, 38, 39] and unpublished sources. ‡ The actual age of eligibility was less than 7 in these years, but because of extremely low uptake in the early years of the CSG we have modelled the effect of low uptake by assuming lower eligibility thresholds.

In the interests of simplicity, we assume that for every child that meets both the age and means test criteria, the CSG is paid to the household in which the child lives. In reality there is a significant minority of children who meet the eligibility criteria but do not receive the grant. For example, Woolard and Leibbrandt [33] found that although 70% of age-eligible children are eligible to receive the CSG, only 60% actually receive the CSG. The 10% gap is likely to be due to delays in accessing the grant, uncertainty about eligibility criteria and difficulties in determining eligibility, particularly for maternal orphans [33].

##### 1.3.3 State old age pension

The state age old pension has been provided since 1928, but the amount of the pension historically differed by race, and it was only in 1993 that the amount was equalized across race groups [33, 39]. The age of eligibility also differed historically between men and women: until 2008 the age of eligibility was 60 in women and 65 in men, but by 2010 the age of eligibility had changed to 60 for both men and women [39]. As with the CSG, a means test is applied, which has changed over time, and which differs according to whether the individual is single or married.

Our assumptions about the monthly amount of the state pension are summarized in Table S14. In the interests of simplicity we assume the same pension amount for all race groups before 1993, as data before 1993 are lacking. Although means test thresholds are applied in practice, these are not used in our model because we assume that people over the age of 65 are not employed, and that people automatically qualify for the state pension if they do not have a private pension. As in the previous section, we assume that all individuals who are eligible to receive the state pension actually receive the pension, although in reality some 20% of those eligible might in fact not receive the state pension [39]. We assume that in the period after 2021 the pension amount increases by 4.0% per annum, the average annual growth in the amount of the state pension over the 2016-2021 period.

##### 1.3.4 Private pensions

In our model we assign to each individual over the age of 60 an indicator: 0 if they receive no private pension, and 1 if they do. This indicator is randomly assigned at the point they reach age 60 and is assumed to remain fixed thereafter. The probability of being assigned a private pension is calculated from a multivariable logistic regression model that is calculated from the 2018 GHS data (Table S15). A limitation of this dataset is that the question about income from private pensions is asked at the household level rather than the individual level. It is therefore not possible to tell if multiple people in the household are receiving a private pension, or even if the identity of the pensioner matches that of the household head. However, as noted previously, in most households the oldest member is reported to be the household head, and this is especially likely if they are receiving a pension. We therefore assume for the sake of this analysis that if a household receives a private pension it is the household head that receives the pension. Table S15 thus shows the relationship between the characteristics of the household head and the household receipt of a private pension. As might be expected, private pensions are most frequently reported in individuals with higher levels of educational attainment and (given historically higher savings) whites and Asians. The effects of sex and urban/rural location were not significant. The odds of the household receiving a private pension increase by a factor of 1.76 for each additional member aged 60 or older, suggesting that individuals other than the household head might also be receiving a pension.

Table S15: Factors associated with receiving a private pension

| | aOR (95% CI) |
|---|---|
| Female sex (ref. male) | 0.93 (0.75-1.15) |
| Educational attainment | |
| None | 1 |
| Primary | 1.35 (0.85-2.16) |
| Incomplete secondary | 4.91 (3.19-7.55) |
| Completed secondary | 14.2 (9.1-22.2) |
| Tertiary | 14.0 (8.6-22.8) |
| Race | |
| Black African | 1 |
| 'Coloured' | 1.37 (0.99-1.88) |
| White/Asian | 2.54 (2.00-3.23) |
| Urban location (ref. rural) | 0.97 (0.76-1.26) |
| Number of household members aged 60 or older | 1.76 (1.44-2.15) |
| Constant (base odds) | 0.011 (0.007-0.019) |

Source: 2018 General Household Survey (author's own calculations). The analysis is limited to households in which there is at least one household member aged 60 or older.

We calculate the probability of receiving a private pension from the constant term and the education and race effects in the above table. For example, the odds of receiving a private pension for a 'coloured' South African who has completed high school is 0.011 × 14.2 × 1.37 = 0.21, and the associated probability is 0.18 (0.21/1.21).

The assumed amount of the private pension is R8 239 per month in 2019-20, based on records of 649 000 banked private pensions in South Africa [35]. We assume that this amount increases by 3% per annum in real terms, based on the same data source, and further adjust the pension amount for annual changes in the South African Consumer Price Index. Although one could use the pension data in the General Household Surveys to estimate the annual average private pension, this may be less reliable than using the data from the banking industry, as the survey may be affected by a number of sampling biases, and the mean income is particularly susceptible to outliers in the survey sample. We do not attempt to model the variation in the amount of the private pension. This simplifies the model as the amount of the private pension is always above the income threshold for the state old age pension, and thus we can assume for simplicity that people who receive private pensions do not qualify for state pensions.

##### 1.3.5 Model validation

Although this is not intended to be an accurate model of income in South Africa, it is nevertheless useful to assess the extent to which the model is consistent with published income statistics (Figure S5). The model is roughly consistent with median income levels, though slightly lower than published estimates, which might be because our model does not consider all sources of income. Our model estimates of the Palma ratio (the ratio of incomes in the top decile to that in the four lowest deciles) appear slightly too high, but our estimate of the Gini coefficient (the more commonly quoted metric of income inequality) is in good agreement with published estimates. Notably, the Palma ratio suggests a steep decline in income inequality after the rollout of the CSG in the early 2000s, whereas the Gini coefficient has declined more gradually. The model slightly under-estimates the extent of income inequality among black South Africans in recent years, but slightly overstates the level of income inequality among white and Asian South Africans.

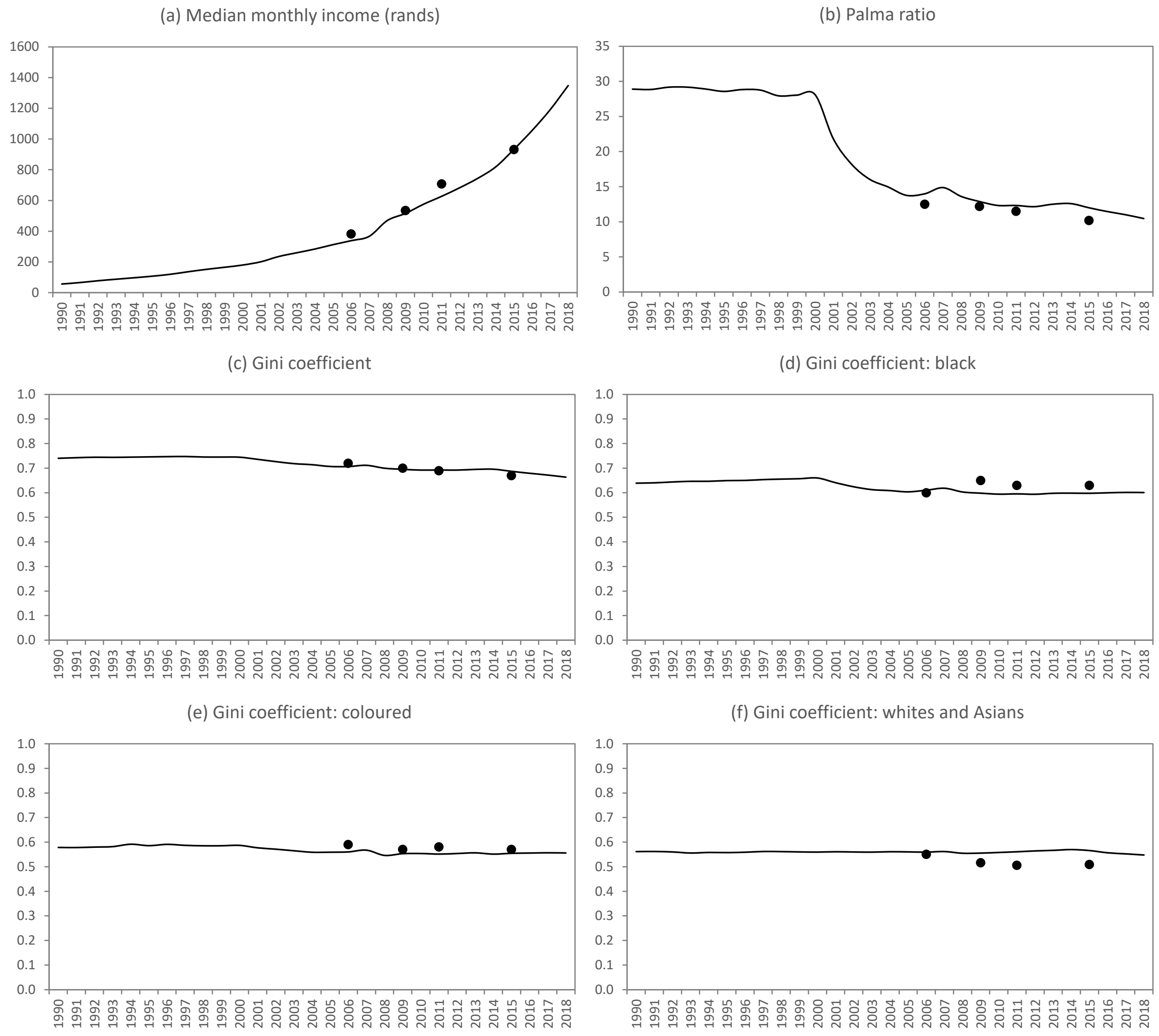

Figure S5: Income and income inequality trends in South Africa

Solid lines represent averages from model simulations; dots represent data from national surveys [14]. Monthly income levels are nominal (not adjusted for inflation). Gini coefficients have been approximated for whites and Asians combined, because MicroCOSM aggregates the white and Asian populations.

##### 1.3.6 Limitations

A limitation of our model of household income is that the four sources of income that we consider together account for only about 90% of total household income. The most significant omission from this model is likely to be investment income, which unfortunately is not quantified in the GHS, and remittances (which account for around 10% of household income in poorer households, on average [33]). We have also not considered the value of agricultural goods produced for self-consumption. A 1993 survey estimated that this accounted for only 4% of income, both in poor and non-poor households [32], and more recent analyses of NIDS data from 2008 and 2017 suggest proportions of 0.2-0.5% (Julius Ohrnberger, personal communication), consistent with significant 'deagrarianization' in recent decades [40].

Our model of salary income is also simplistic. Although we account for inter-individual variation in income among employed individuals (even after controlling for differences in age, sex, education and race), the income deviation score assigned to each individual is assumed to remain fixed over time. In reality, this income deviation score is likely to vary over time, but we have relied here on GHS data, which are cross-sectional; a longitudinal dataset such as the NIDS would be required to determine the extent of the temporal stability. We have also not attempted to model the sources of this inter-individual variation; although we model the contribution of conscientiousness, important contributors such as cognitive ability [36] and health [41] are not included. Another limitation is that we lack reliable income data from the period before the introduction of the GHS in 2002, and have therefore had to use simple extrapolation for the pre-2002 period. Although attempts were made to analyse earlier income data from the October Household Surveys (1993-1999), results were inconsistent with the later GHSs, and it appeared that there were problems with the reported frequencies of incomes (weekly/monthly/annual). We therefore chose not to use the data from the earlier household surveys. Income surveys before 1993 excluded the so-called 'homeland states' and were therefore not nationally representative [32]. The finding of a steep decline in the Palma ratio after 2000 needs to be treated with caution, as this reflects the assumption of stable salary distributions in the period before 2002. Others have argued that inequality in non-grant income increased substantially between 1993 and 2008 [31], which would suggest that salary distributions have not remained stable.

A more realistic model of private pensions would allow for individual variation in the amount of the pension received, not just the probability of receiving a private pension. A more realistic model would also allow for the possibility that people with private pensions might qualify for state pensions (if their private pension and savings fall below the means test threshold for the state pension). It would also be more realistic to link the probability of having a private pension (and the amount of that pension) to the individual's employment history over their working life. However, as private pensions account for only about 2% of all household income, and as relatively few South Africans have private pensions, this additional detail is unlikely to change our conclusions about the variation in household incomes across the South African population.

Despite its simplicity, our model is roughly consistent with reported median income levels and measures of income inequality in South Africa, making it an appropriate tool for simulating the impact of poverty and income inequality on HIV risk behaviours.

#### 1.4 Effects of socio-economic status on sexual behaviour

When modelling the effects of socio-economic status on sexual behaviour, a number of practical and theoretical challenges arise. The first (practical) challenge is that there are several sexual behaviour parameters, a number of different socio-economic status variables, and the effects of these variables on sexual behaviour may differ by sex – thus there is potentially a large number of parameters to be modelled. For example, if there are six sexual behaviour parameters and three socio-economic variables, this would potentially imply 18 effect parameters to estimate for each sex (36 parameters in total). This presents a challenge as we lack data on many of these effects, and even when data might exist on one socio-economic effect, it is often unclear how much of the observed effect is explained by other (unobserved) socio-economic effects. (For example, a study might show an association between education and condom use but not control for either employment or income, making it difficult to assess how much of the 'effect' of education is independent of employment and income.) In the interests of ensuring a parsimonious model, we limit ourselves to one socio-economic effect per sexual risk behaviour – the other socio-economic variables are assumed to have no effect (see Table S16). In deciding which socio-economic variable to link to each risk behaviour, we are guided by the following principles:

- According to the Health Belief Model, an individual's perceived susceptibility to disease, perceived barriers to action, exposure to factors that prompt action and self-efficacy all determine the extent to which they change their risk behaviour [42]. These factors are all likely to be linked to their educational attainment, and thus the Health Belief model posits that people with higher educational attainment are more likely to change their risk behaviour in respond to the threat of HIV. In South Africa, we have previously noted that the only significant change in sexual behaviour that can be clearly linked to the HIV response is increased condom use [43]. We thus assume levels of condom use to be linked to educational attainment.
- When considering sexual risk behaviours that have not changed in response to HIV, we assume the primary determinant of sexual risk behaviour is either income or employment status, as these are more proximal measures of economic deprivation than educational attainment. Literature suggests that in African settings, household income is allocated differently depending on whether it is earned by male or female household members, with more income being spent on alcohol and cigarettes when it is earned by male members and proportionately more income being spent on food and child health when income is earned by female household members, even when controlling for the level of household income [44, 45]. This suggests that men's ability to engage in risk behaviours (alcohol, smoking, extramarital partners) is determined principally by the income that they directly earn, rather than their household income; for women, on the other hand, the average per capita household income may be a better predictor of socio-economic status, as any individual income they earn is more likely to be shared with other household members. We therefore rely primarily on employment status to model the effect of male socio-economic status on sexual risk behaviour, and adjusted per capita household income (APCHI, as defined in the main text) to model the effect of female socio-economic status on sexual risk behaviour.
- After controlling for educational attainment, being in school is not a socio-economic effect but rather a network effect: being in school constrains youth contact with older out-of-school individuals and potential sexual contacts. We therefore model effects of being in school on early sexual risk behaviour (specifically sexual debut and marriage), but do not consider these to be socio-economic effects.
- Entry into marriage is technically not a 'sexual risk behaviour', and indeed African literature is conflicting as to whether marriage increases or reduces HIV risk [46-49]. Rates of marriage are strongly associated with socio-economic status, but as discussed below (in section 1.4.6) it is difficult to determine which measure of socio-economic status most directly determines marriage rates. We have assumed a dependence on educational attainment, as this is the socio-economic variable for which we have the strongest evidence of an effect on marriage.

Table S16: Overview of modelled socio-economic effects on sexual behaviour

| Sexual behaviour outcome | Current schooling M | Current schooling F | Educational attainment M | Educational attainment F | Employment status M | Employment status F | Household income M | Household income F |
|---|---|---|---|---|---|---|---|---|
| Sexual debut | ✓ | ✓ | | | | | | ✓ |
| Casual sex | | | | | ✓ | | | ✓ |
| Commercial sex | | | | | ✓ | ✓ | | |
| Short-term non-cohabiting relationships | | | | | ✓ | | | ✓ |
| Condom use | | | ✓ | ✓ | | | | |
| Marriage/cohabitation | ✓ | ✓ | ✓ | ✓ | | | | |

Another challenge in developing our model is that it is difficult to infer causality from observed associations between socio-economic status and sexual risk behaviour. For example, there is heterogeneity between individuals in their personal discount rates (the extent to which they are willing to trade off immediate gains and larger long-term gains), and individuals with higher discount rates may be less likely to invest in their education and career, as well as more likely to engage in health risk behaviours [50]. This could explain some of the observed associations between sexual risk behaviour and low socio-economic status. To some extent, our model does capture this source of confounding, as we model heterogeneity between individuals in levels of conscientiousness, which are strongly correlated with personal discount rates [51], and conscientiousness is assumed to be correlated with both higher rates of grade completion, higher income (see section 1.3.1) and lower rates of sexual partner concurrency [30]. However, the critical point is that published analyses of associations between socio-economic status and sexual risk behaviour are seldom able to control for these confounding psychological/ personality factors. We therefore aim to represent the uncertainty regarding socio-economic effects using 'hurdle distributions', with discrete probabilities attached to a zero (null) effect. This is described more fully in section 2.1.

Finally, there is uncertainty as to whether it is *relative* deprivation or deprivation in an 'absolute' sense that drives sexual risk behaviour. According to the relative deprivation hypothesis, relative socio-economic position is critical to determining one's health status and willingness to take risks [52-54]. This implies that it is levels of income inequality in a society that are critical to determining levels of risk behaviour, rather than absolute measures of wealth (such as per capita GDP). In our model we rely on a combination of absolute and relative measures of socio-economic status when modelling effects on sexual risk behaviour: educational attainment and employment status are absolute measures, whereas per capita household income is a relative measure (expressed relative to the national mean, on a log scale). This means that an intervention that raises incomes by the same proportion across all income groups will have little impact on sexual risk behaviour, because the poorest individuals will maintain the same relative level of deprivation relative to the national average income level. However, interventions that raise levels of education or employment will be expected to influence sexual behaviour more materially, regardless of whether they apply uniformly across the population. Because randomized trials of economic interventions tend to target the poorest individuals, it is difficult to elucidate whether the effects they observe conform to absolute or relative impacts of socio-economic status. To mitigate the model uncertainty, we similarly focus here on interventions that are targeted to households with income levels below the national mean (on the log scale).

##### 1.4.1 Sexual debut

The hazard function for the rate of sexual debut in the baseline category (black youth in the high-risk group, who are out of school and with household income at/above the national mean) is assumed to be log-logistic in form, with an offset of 10 to prevent sexual debut at ages below 10. The baseline hazard is set separately for males and females. Mathematically, the probability that an individual in the baseline category, aged *x* and of sex *g*, is sexually experienced is

$$F_g(x) = \frac{1}{1 + \left((x-10)/(m_g - 10)\right)^{-\beta_g}}$$

for *x* > 10. In this equation, $m_g$ is the median age at sexual debut, and $\beta_g$ is the shape parameter that determines the extent to which the rate of sexual debut changes in relation to age. The annual rate of sexual debut in the baseline category, at exact age *x*, is then

$$h_g(x) = \frac{\beta_g (x-10)^{-1}}{1 + \left((x-10)/(m_g - 10)\right)^{-\beta_g}}$$

The annual rate of sexual debut at age *x*, for an individual of sex *g*, in risk group *i* and population group *r*, with per capita household income *c* and schooling status *s* (0 if out of school, 1 if in school) is then

$$D_{g,i,r,s,c}(x) = h_g(x) R_i P_r E_g^s H_g(c)$$

where $R_2$ is the relative rate of sexual debut in the low-risk group (relative to the high-risk group), $P_r$ is the relative rate of sexual debut in race group *r* (relative to black African), $E_g$ is the relative rate of sexual debut in youth who are in school and

$$H_g(c) = \begin{cases} 1 & \text{if } g = 0 \text{ or } c \geq \bar{c} \\ (1+K)^{\log(\bar{c}) - \log(c)} & \text{if } g = 1 \text{ and } 0 < c < \bar{c} \\ (1+K)^{\log(\bar{c})} & \text{if } g = 1 \text{ and } c = 0 \end{cases}$$

where *K* is the proportional increase in the rate of sexual debut in girls per log reduction in per capita household income below the national mean ($\bar{c}$). Table S17 summarizes the values assigned to the different parameters. In most cases, these are set to be the same as in the original specification of the MicroCOSM model [1], but the median and shape parameters have been adjusted to ensure consistency with the calibration data is preserved after incorporating the income and schooling effects described below.

Table S17: Sexual debut parameters

| Symbol | Description | Value | Source |
|---|---|---|---|
| $m_0$ | Baseline group median age at sexual debut: males | 16.5 | Calibrated |
| $m_1$ | Baseline group median age at sexual debut: females | 15.5 | Calibrated |
| $\beta_0$ | Shape parameter: males | 7.5 | Calibrated |
| $\beta_1$ | Shape parameter: females | 11 | Calibrated |
| $R_1$ | Relative rate of debut: high-risk group | 1 | - |
| $R_2$ | Relative rate of debut: low-risk group | 0.58 | [1] |
| $P_0$ | Relative rate of debut: black African | 1 | - |
| $P_1$ | Relative rate of debut: 'coloured'/mixed race | 0.75 | [1] |
| $P_2$ | Relative rate of debut: white | 0.47 | [1] |
| $E_0$ | Relative rate of debut: boys in school | 0.80* | [55-57] |
| $E_1$ | Relative rate of debut: girls in school | 0.46* | [55-57] |
| $K$ | Proportional increase in rate of debut in girls, per log reduction in per capita household income | 0.23* | [58-60] |

* Allowed to vary in the model calibration process.

The $m_g$ and $\beta_g$ parameters have been chosen in such a way that the modelled levels of sexual experience at each age are consistent with the results of various national surveys [61-63], as shown in Figure S6. For the purpose of this comparison, we have calculated the average age-specific prevalence of sexual experience across the three surveys; however, for women we have adjusted the reported rates by an odds ratio of 2, to reflect likely under-reporting of sexual experience in young women [64, 65]. The model results show the prevalence of sexual experience in 2005, averaged across 50 simulations.

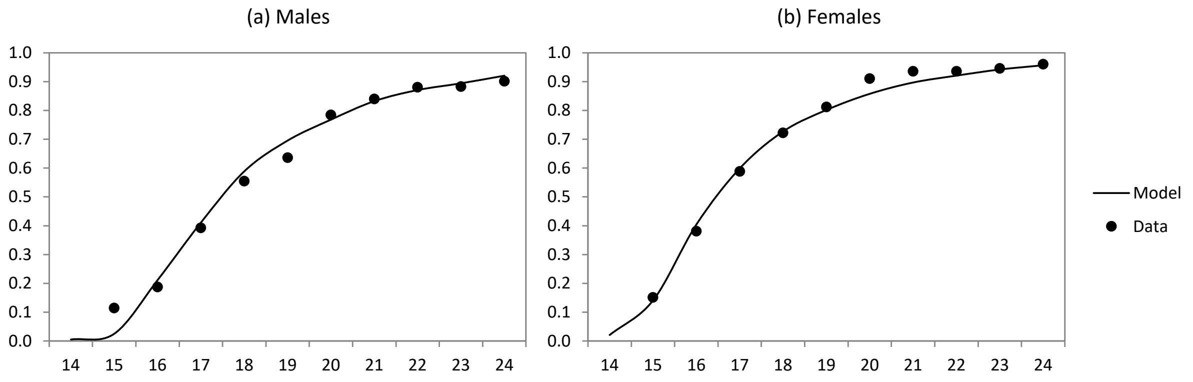

Figure S6: Fraction of youth who are sexually experienced, by age

Longitudinal studies in South Africa suggest that being in school is associated with a reduced rate of sexual debut, both in boys and girls [55-57], and reduced incidence of pregnancy [66]. However, these studies may be affected by confounding (for example, if common risk factors like poverty and sensation-seeking personality influence both school dropout and sexual debut), or may be biased by differential social desirability bias (for example, greater social desirability bias among girls in school [65]). Randomized trials of school support interventions have found reduced rates of sexual debut, which are likely to be mediated by reductions in school dropout [67-70], although the only trial to report results for boys and girls separately found that the intervention reduced sexual debut in girls only [67]. In a Malawian RCT, the relative rate of debut among girls who were in school at baseline to that in girls out of school was around 0.3 [71], similar to the hazard ratio of 0.24 in a longitudinal study in South African girls [55], but lower than the odds ratio of 0.67 in another South African cohort study [57]. To represent the uncertainty in the $E_1$ parameter, we use a beta hurdle distribution, with a weight of 0.1 given to a value of 1 (no effect) and a weight of 0.9 given to a beta distribution with a mean of 0.4 (the average of the values in the three previously cited studies) and standard deviation of 0.23.

The two previously-cited South African studies suggest relative rates of sexual debut for boys in school of 0.69 [55] and 0.52 [57]. To represent the uncertainty in the $E_0$ parameter, we use a beta hurdle distribution, with a weight of 0.5 given to a value of 1 (based on the lack of evidence from RCTs) and a weight of 0.5 given to a beta distribution with a mean of 0.6 (the average of the values in the two previously cited studies) and standard deviation of 0.23.

Southern African studies also suggest an effect of household income on the rate of sexual debut in girls, but not in boys [58-60]. In a longitudinal study in Cape Town communities, it was found that each log reduction in per capita household income was associated with a roughly 1.09-fold increase in the odds of sexual debut in girls (with an upper bound of 1.19) [58]. In a similar study in Zimbabwe, each standard deviation decrease in household wealth was associated with a 1.25-fold increase in the odds of sexual debut in girls (95% CI: 1.06-1.46) [59]. Household wealth is probably a good proxy for per capita household income, and in the Cape Town study the standard deviation of per capita household income (on a log scale) was 1.00 [58], suggesting approximate equivalence in the 'per log' and the 'per standard deviation' scales. RCTs also suggest that cash transfer interventions delay sexual debut in girls [71-74], but RCTs that report results for boys find little evidence of positive effects [72, 74, 75]. Based on these studies, we represent the uncertainty in the $K$ parameter using a gamma hurdle distribution with a weight of 0.1 given to the value of 0 (no effect) and a weight of 0.9 given to gamma distribution with mean 0.25 and standard deviation of 0.15.

##### 1.4.2 Casual/transactional sex

The model of casual sex is the same as described in the supplementary materials of our previous publication [30]. Briefly, individuals are assumed to move in and out of casual sex 'phases' depending on their age, risk group, relationship status, socio-economic status and level of binge drinking. We define casual sex relationships as relationships that last less than a week (mostly involving a single sex act), and assume that 'phases' of casual sex activity last for a year on average. A 'base rate' of entry into the casual sex state is specified for single high-risk individuals aged 17.5 who never engage in binge drinking. In men this base rate is further defined as applying to unemployed men with an average inequitable gender norm score (0.2), and in women the base rate is further defined as applying to women whose adjusted per capita household income is at or above the national average. A series of multipliers are applied to the base rate of casual sex to represent the effects of risk group, relationship status, age, binge drinking, inequitable gender norms and socio-economic status (detailed in Table S18). These are the same as assumed previously [30]. The age adjustments (not shown in Table S18) are the same as for short-term relationships (described in section 1.4.4).

Table S18: Model assumptions about heterosexual rates of entry into the casual sex state

| Parameter | Male | Female |
|---|---|---|
| Annual rate of entry in high-risk unpartnered individuals aged 17.5 | 0.28* | 0.20 |
| Relative rate of entry in unpartnered low-risk individuals | 0.40 | 0.12 |
| Relative rate of entry in partnered high-risk individuals | 0.80 | 0.80 |
| Relative rate of entry if binge drinking at least once a month | 1.39 | 1.46 |
| Relative rate of entry per 0.1 decrease in inequitable gender norm score | 0.81 | - |
| Increase in rate of entry if employed (RR – 1) | 0.25 | 0 |
| Increase in rate of entry per log reduction in per capita household income† | 0 | 0.425 |

* In men who have a propensity for same-sex relationships, this rate is reduced in proportion to their preference for male partners. † For women in households with per capita household income below the national average. RR = relative rate.

Although our model considers only 'casual' sex, we rely on data regarding both transactional and casual sex when parameterizing our model, given the lack of data on casual sex specifically, and given that there is often significant overlap between casual and transactional sex in the local context [76, 77]. Cross-sectional South African studies suggest an association between male engagement in transactional/casual sex and employment status. In a study of young men in the Eastern Cape, reporting of transactional sex with casual partners was associated with having ever earned money (aOR 1.92, 95% CI: 1.37-2.69), and was more weakly associated with household socio-economic status [78]. In our analysis of the 2016 South African DHS data [79], employment was moderately associated with male reporting of transactional sex, although this was not statistically significant (aOR 1.33, Table S19), and household wealth appeared to be a more significant predictor of male engagement in transactional sex (results not shown). Evidence from Zimbabwe also suggests a strong positive association between household wealth and male reporting of casual sex, but this study did not assess the effect of male employment on casual sex [59]. Studies have generally not found male education to be strongly associated with reporting of transactional or casual sex [78, 80, 81]. Given that male employment and household socio-economic status are strongly correlated, we have chosen to model only the effect of male employment on casual sex. We represent the uncertainty in this effect using a gamma hurdle distribution, with a weight of 0.5 given to the null effect (because there is no RCT evidence to suggest an effect of male socio-economic status on casual sex) and a weight of 0.5 given to a gamma distribution with a mean of 0.5 and standard deviation of 0.25. The latter gamma mean corresponds to a relative risk of 1.5, in between the estimates of 1.33 and 1.92 from the previously cited South African sources [78, 79].

Table S19: Predictors of transactional sex in South Africa (adjusted odds ratios, 95% CI)

| | Male | Female* |
|---|---|---|
| Age | 1.34 (1.18-1.52) | 15.8 (1.91-130.7) |
| Age-squared | 0.996 (0.994-0.998) | 0.935 (0.890-0.982) |
| Married | 0.44 (0.28-0.68) | - |
| Condone wife beating | 1.73 (1.04-2.88) | - |
| Employed | 1.33 (0.91-1.95) | - |
| Lowest wealth quintile | - | 2.07 (0.94-4.57) |

Source: 2016 DHS (author's own calculations)
* Limited to unmarried women, as there were no married women who reported transactional sex.

Studies in other African countries have found that women's reporting of transactional sex is negatively associated with their educational attainment [80], employment status [82] and household wealth [59]. In a South African study, women's reporting of transactional sex was negatively related to educational attainment and standard of housing (aOR of 1.72 for substandard housing), but paradoxically it was positively related to employment [83]. Another South African study found that women's reporting of recent transactional sex was not significantly related to their education or employment, but was significantly higher in poor households (specifically households without electricity and/or piped water – aORs were 3.6 and 2.0 respectively) [84]. A further South African study found that women's reporting of transactional sex was positively related to household food insecurity (OR 1.9), but did not assess the role of education or employment status [85]. We assessed predictors of transactional sex in the 2016 South African DHS, but due to the small number of women reporting transactional sex (27), it was difficult to identify significant correlates. Women's reporting of transactional sex was significantly negatively associated with their educational attainment but was not associated with their employment status (results not shown). The effect of household wealth was non-linear, with a substantially higher (and nearly significant) level of transactional sex in the lowest wealth quintile, but no significant difference between the other four wealth quintiles (Table S1.19). The observational evidence is therefore not completely consistent, but measures of household poverty appear to be most consistently associated with transactional/casual sex. The effect of household wealth appears to be non-linear, and we therefore model the effect of per capita household income by assuming an increase in the rate of entry into casual sex per log increase in per capita household income below the national average (while the rate is the same for all per capita income levels at or above the national average).

Although observational studies suggest an effect of socio-economic status on women's entry into transactional/casual sex, RCTs have found only very modest (non-significant) effects of economic strengthening on women's reporting of transactional sex [73, 86]. This makes it difficult to exclude the possibility that the 'effects' seen in observational studies are due to confounding factors. We therefore represent the uncertainty in the proportional increase in female entry into casual sex, per log reduction in per capita household income, using a gamma hurdle distribution with a weight of 0.5 attached to a zero value (no effect) and the remaining 0.5 weight given to a gamma distribution with a mean of 0.85 and standard deviation of 0.40. With the mean of 0.85, the ratio of the rate of casual sex entry at the $10^{th}$ decile of household incomes to that at the $50^{th}$ decile is 2.2 ($(1 + 0.85)^{1.3}$, assuming a roughly 1.3 log difference in per capita household income between the $10^{th}$ and $50^{th}$ percentiles [58]). This is roughly consistent with the average of the odds ratios from the South African observational studies comparing the poorest households to 'normal' households.

Table S20 shows that when taking the average results from 50 simulations, the model estimates in 2005 are roughly consistent with casual sex calibration targets [77, 83]. These calibration targets are explained in more detail in the supplementary materials of our previous work [30]. In addition, the model estimates levels of employment in men with multiple recent partners consistent with those in the study of Townsend *et al* [77], which validates the model assumptions about the effects of employment on casual sex and short-term relationships.

Table S20: Heterosexual casual sex model outputs in 2005

| Model output | Value | Calibration target |
|---|---|---|
| % of 15-49 year old men engaging in heterosexual casual sex | 14% | - |
| % of 15-49 year old women engaging in casual sex | 8% | - |
| % of men with multiple partners who have had casual sex in the last 3 months | 72% | 71% |
| Average # partners in the last 3 months, for men who report multiple partners in the last 3 months | 5 | 5 |
| % of men with multiple partners in the last 3 months who are 'high-risk' | 95% | 94% |
| % of men with multiple recent partners who are employed | 62% | 59%* |
| % of pregnant women who have ever had casual sex | 28% | 29% |
| % of pregnant women reporting a history of casual sex who also report a history of concurrent partnerships | 72% | 66% |

* This is a validation rather than a calibration target, relevant in the context of the current study.

##### 1.4.3 Commercial sex

*Male demand for commercial sex*

For a sexually experienced man with ID $u$, the annual rate of contact with female sex workers, $w(u)$, is assumed to depend on his age ($x_u$), race ($r_u$), urban/rural location ($b_u$), same-sex preference ($Y_u$), risk group ($i_u$), marital status ($l_u$), number of current partners ($j_u$), HIV status/stage ($s_u$) and employment status ($E_u$). A gamma probability density function is used to represent the age differences in rates of male contact with sex workers. The following formula is used to calculate $w(u)$:

$$w(u) = w\lambda^{\alpha}(x_u - 10)^{\alpha-1}\exp\left(-\lambda(x_u - 10)\right)T_{i_u,j_u,l_u}(1 - Y_u)J(b_u)\Psi_{j_u}(r_u)\Phi(s_u)\eta(E_u)$$

where $\lambda$ and $\alpha$ are the parameters of the gamma probability density function, $T_{i,j,l}$ is an adjustment factor to represent the man's risk group and relationship status, $J(b_u)$ is an adjustment factor to represent the effect of urban/rural location, $\Psi_{j_u}(r_u)$ allows for racial differences in the incidence of concurrency, and $\eta(E_u)$ represents the effect of employment status. The assumed values of the parameters are summarized in Table S21. Incarcerated men and virgins are assumed to have no contact with sex workers.

The base rate of sex worker contact ($w$, which applies to high risk men who currently have no partner and are unemployed) has been set in such a way that the demand for commercial sex is sufficient to match the estimated size of the South African sex worker population in a 2011 study [87], when it is assumed that sex workers have 750 clients per annum on average [88-95]. Using this sex worker population size estimate and the assumed number of 750 clients per annum implies roughly 5 sex worker contacts per annum for men aged 15-49. Hence the $w$ parameter has been calculated so that the average number of sex worker contacts per year, averaged across all men aged 15 to 49 at the start of the simulation, is 5, i.e.

$$w \equiv \frac{N(R_1)}{\sum_{u\in R_1}\lambda^{\alpha}(x_u - 10)^{\alpha-1}\exp\left(-\lambda(x_u - 10)\right)T_{i_u,j_u,l_u}(1 - Y_u)J(b_u)\Psi_{j_u}(r_u)\Phi(s_u)\eta(E_u)}$$

where $R_1$ is the set of all men aged 15-49 who are alive, and $N(R_1)$ represents the number of men in this set.

Table S21: Assumed male rates of sex worker contact

| Parameter | Value | Source/explanation |
|---|---|---|
| $\lambda$ | 0.264 | Based on fitting age pattern of male |
| $\alpha$ | 8.46 | contact with SW in 2016 DHS |
| $T_{i,j,l}$ for $i$=1 and $j$=0 | 1 | Base rate (unpartnered men) |
| $T_{i,j,l}$ for $i$=1, $l$=0 and $j$=1 | 0.5 | Assumption |
| $T_{i,j,l}$ for $i$=1, $l$=1 and $j$=1 | 0.3 | Assumption |
| $T_{i,j,l}$ for $i$=1, $l$=0 and $j$=2 | 0.2 | Assumption |
| $T_{i,j,l}$ for $i$=1, $l$=1 and $j$=2 | 0.1 | Assumption |
| $T_{i,j,l}$ for $i$=2 | 0 | Definition of low-risk group |
| $J(0)$ (urban effect) | 1.5 | [96, 97] |
| $J(1)$ (rural effect) | 0.5 | [96, 97] |
| $\Psi_j(r)$ for $r$=0 and for $j$=0 | 1 | Base rate (unpartnered men) |
| $\Psi_j(1)$ for $j\neq0$ | 0.09 | Calibration to concurrency data [1] |
| $\Psi_j(2)$ for $j\neq0$ | 0.02 | Calibration to concurrency data [1] |
| $\Phi(s)$ for $s$ = 0, 1 or 2* | 1 | |
| $\Phi(s)$ for $s$ = 3* | 0.65 | Same assumptions as for short-term |
| $\Phi(s)$ for $s$ = 4* | 0.25 | partnerships (see section 1.4.4) |
| $\Phi(s)$ for $s$ = 5 or 6* | 0.8 | |
| $\eta(1)$ | 1.4 | [96, 98, 99] |

* HIV stages are defined as acute HIV ($s$ = 1), ART-naïve with CD4 ≥ 350 ($s$ = 2), ART-naïve with CD4 200-349 ($s$ = 3), ART-naïve with CD4 < 200 ($s$ = 4), on ART ($s$ = 5) or interrupting ART ($s$ = 6). DHS = Demographic and Health Survey, SW = sex worker.

Most of the parameters in Table S1.21 are the same as assumed in earlier versions of MicroCOSM, but the model has been extended to allow for an effect of HIV status/disease stage on sex worker contact (consistent with that assumed for short-term relationships) and the age-related parameters have been updated to ensure the age pattern of male contact with sex workers is consistent with that reported in the 2016 DHS (Figure S7).

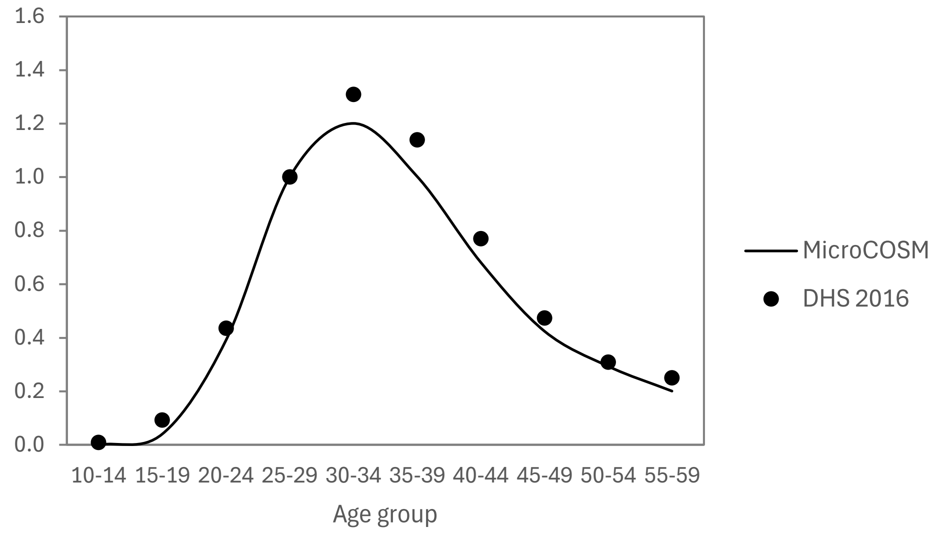

Figure S7: Rate of male contact with sex workers relative to age 25-29
DHS 2016 estimates are obtained by fitting a logistic regression model to the DHS male reports of sex with a sex worker in the last year, using a cubic polynomial to represent the age relationship. MicroCOSM estimates are obtained by dividing the total number of male contacts with sex workers in each age group by the modelled population size in that age group. Both sets of estimates are expressed as relative rates (dividing by the rate at age 25-29); we do not attempt to match absolute levels of male reporting of sex worker contact due to concerns about under-reporting of sex worker contact.

There is mixed evidence on the effect of men's socio-economic status on their rate of contact with sex workers. Leclerc and Garenne [96] found that among Zambian men, higher educational attainment and greater wealth were both associated with lower reporting of contact with commercial sex workers, but the effect of employment was not significant. A more recent analysis of DHS data from across sub-Saharan Africa also found higher wealth and tertiary education to both be moderately associated with lower rates of male reporting of recently paying for sex, but being employed was strongly associated with paying for sex (aOR 1.73, 95% CI: 1.58-1.90) [99]. A South Africa study found that higher income (but not education) predicted male sex with a sex worker [100], and in another analysis of the same data, male contact with sex workers was found to be significantly associated with recent employment (OR 1.86, 95% CI: 1.42-2.46) [98]. However, a limitation of the South African study is that it asked about having *ever* had sex with a sex worker (unlike the other two studies, which asked about contact with a sex worker in the last 12 months), and men's past employment status/income may have differed from that at the time of the survey. We assume that household wealth and male employment are strongly correlated, and that it is the latter that more directly determines their rate of contact with sex workers. To represent the uncertainty around the effect of employment on male rate of sex worker contact, we use a gamma hurdle distribution with a weight of 0.5 given to the zero value (no effect) and a weight of 0.5 given to a gamma distribution with a mean of 0.8 and standard deviation of 0.4. The mean of 0.8 corresponds to a relative risk of 1.8, which is consistent with both the South African study [98] and the analysis of African DHS data [99].

*Female entry into commercial sex*

Rates of female entry into commercial sex are set such that there is sufficient ‘supply’ of commercial sex to meet male ‘demand’, based on the previously stated assumption that sex workers have 750 clients per annum. The number of new sex workers required over the period $[t, t + d)$ in order to satisfy male demand, $\Delta(t, t + d)$, is calculated as

$$\Delta(t, t + d) = \frac{1}{C}\sum_{u \in R_2} w(u) - N(R_3)$$

where $C$ is the assumed annual number of clients per sex worker (750), $R_2$ is the set of all men who are sexually experienced and not incarcerated at time $t$, and $R_3$ is the set of women engaged in sex work at time $t$. It is assumed that women enter into commercial sex only if they are in the high-risk group, single and unemployed (represented by the set $R_4$). The probability that a woman in the high-risk group, who has no partners and is aged $x$, and who is in HIV disease state $s$ with diagnosis history $v$ (0 if undiagnosed or HIV-negative, 1 if diagnosed), becomes a sex worker over the period $[t, t + d)$ is calculated as

$$\frac{\Delta(t, t + d)W(x, t)\Phi(s)K(v)}{\sum_{u \in R_4} W(x_u, t)\Phi(s_u)K(v_u)}$$

where $W(x, t)$ is the factor by which the rate of recruitment into the ‘sex worker’ group is multiplied when the woman is of age $x$ in order to match the target sex worker age profile, and $\Phi(s)$ is the factor by which the rate of recruitment into commercial sex is adjusted in HIV disease stage $s$ and $K(v)$ is the effect of HIV diagnosis ($K(0)$ is 1 and $K(1)$ is 0.63, consistent with assumptions in the Thembisa model [101]). The age distribution of women starting commercial sex is set to match a target age distribution, which changes over time, in line with a recent review of South African sex worker studies [102]. The same review found that the average duration of sex work in South Africa has increased over time, and based on that we assume a declining rate of exit from commercial sex, over time. Both sets of assumptions about changes in sex worker age and duration of sex work are the same as in the Thembisa model [101].

In earlier versions of MicroCOSM [1], entry into commercial sex was not assumed to be limited to unemployed women. In the updated model, we assume only unemployed women enter sex work, because few South African sex workers report other sources of employment [103-106]. We similarly assume that sex workers who acquire other work discontinue sex work. The model does not consider male sex workers.

##### 1.4.4 Short-term non-cohabiting relationships

Sexually experienced individuals are assumed to acquire new short-term partners at a rate that depends on their age, risk group, race, current relationship status and (in the case of men) employment status. The parameter $c_{g,i,j,l,r,m}^{s}(x)$ is defined as the annual rate at which a sexually-experienced individual of sex $g$ wishes to form new short-term partnerships if they are in risk group $i$ (1 for high risk, 2 for low risk), aged $x$, in race group $r$, in HIV disease state $s$, with $j$ current partners, marital status $l$ (0 for unmarried, 1 for married) and employment status $m$. A gamma probability density function is used to represent age differences in rates of partnership formation. The rate at which individuals wish to form new partnerships is calculated as

$$c_{g,i,j,l,r,m}^{s}(x) = c_g(x - 17.5)^{\alpha_g - 1}\exp\left(-\lambda_g(x - 17.5)\right)\Omega_{g,i,j,l}\Phi(s)\Psi_j(r)\Theta_g(m)$$

where $c_g$ is the desired rate in the baseline group (single, HIV-negative Africans in the high risk group who are aged 17.5 and unemployed), $\lambda_g$ and $\alpha_g$ are the parameters of the gamma probability density function, $\Omega_{g,i,j,l}$ is an adjustment factor taking into account the individual's risk group and current relationship status, $\Phi(s)$ is an adjustment factor that takes into account the individual's HIV status/disease stage, $\Psi_j(r)$ represents an effect of race on acquisition of secondary partners, and $\Theta_g(m)$ represents the effect of employment. The values assumed in the model are summarized in Table S22. Some of these parameter values were previously estimated by fitting a similarly-structured deterministic model to data on numbers of current sexual partners and HIV prevalence data by age and sex [107], but the first three parameters ($c_g$, $\lambda_g$ and $\alpha_g$) have been updated to preserve approximate consistency with the same calibration data (Figure S8).

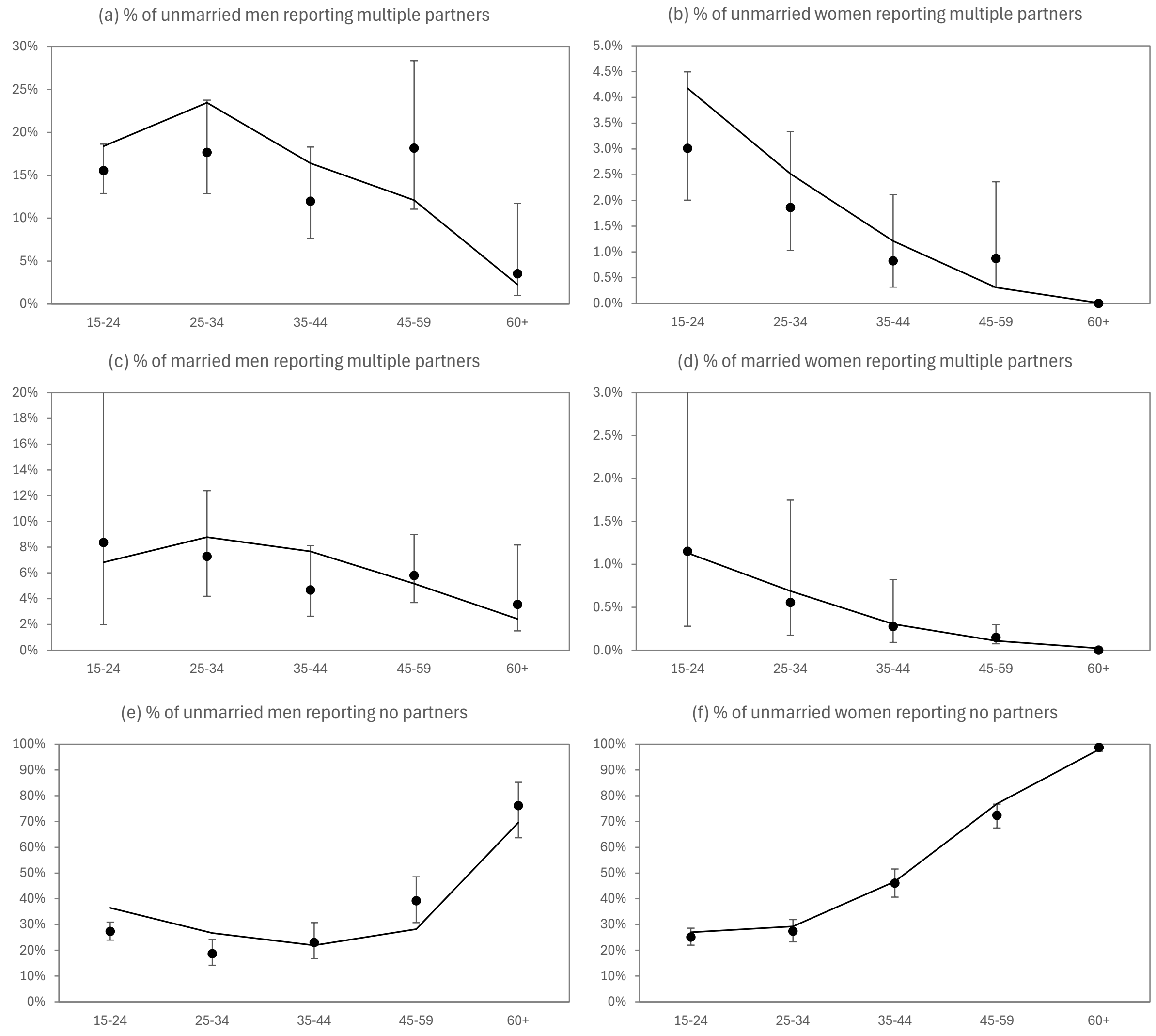

Figure S8: Sexual behaviour of sexually experienced individuals in 2005

Proportions of individuals reporting different numbers of current partners are represented by closed circles (with 95% confidence intervals), based on data from a 2005 national household survey [61]. The mean of the posterior model predictions, after adjustment for estimated reporting bias, is represented by the solid black line. The data and the method for adjusting for reporting bias are described more fully elsewhere [107].

In the present study, the model has been extended to represent the effect of employment, using the equation

$$\Theta_g(m) = \begin{cases} 1 & \text{if } g = 1 \\ (1+Z)^m & \text{if } g = 0 \end{cases}$$

where $Z$ is the proportion by which men's rate of acquiring short-term partners increases if they are employed ($m = 1$).

Table S22: Parameters determining rates of short-term partnership formation, in sexually-experienced adults

| Parameter | Assumed value Males | Females | Source/explanation |
|---|---|---|---|
| $c_g$ | 2.2 | 2.2 | Calibrated |
| $\alpha_g$ | 3.48 | 5.98 | Calibrated |
| $\lambda_g$ | 0.1244 | 0.2716 | Calibrated |
| $\Omega_{g,i,j,l}$ for $i$=1, if $j$=0 | 1 | 1 | - |
| $\Omega_{g,i,j,l}$ for $i$=1, if $l$=0 and $j$=1 | 0.64 | 0.54 | Calibrated [107] |
| $\Omega_{g,i,j,l}$ for $i$=1, if $l$=1 and $j$=1 | 0.41 | 0.17 | Calibrated [107] |
| $\Omega_{g,i,j,l}$ for $i$=1, if $j$=2 | 0 | 0 | Maximum of 2 current partners |
| $\Omega_{g,i,j,l}$ for $i$=2, if $j\neq0$ | 0 | 0 | Definition of low risk |
| $\Omega_{g,i,j,l}$ for $i$=2, if $j$=0 | 0.19 | 0.60 | Calibrated [107] |
| $\Omega_{g,i,j,l}$ for $i$=3 (sex workers) | 0 | 0 | No regular partners assumed for sex workers |
| $\Phi(0)$ | 1 | 1 | - |
| $\Phi(1)^*$ | 1 | 1 | No change in behaviour |
| $\Phi(2)^*$ | 1 | 1 | assumed during early disease |
| $\Phi(3)^*$ | 0.65 | 0.65 | [108-111] |
| $\Phi(4)^*$ | 0.25 | 0.25 | [108-111] |
| $\Phi(5)$, $\Phi(6)^*$ | 0.80 | 0.80 | [112, 113] |
| $\Psi_j(r)$ for $r$=0 and for $j$=0 | 1 | 1 | - |
| $\Psi_j(1)$ for $j\neq0$ | 0.09 | 0.09 | Calibrated [1] |
| $\Psi_j(2)$ for $j\neq0$ | 0.02 | 0.02 | Calibrated [1] |
| $Z$ | 0.32 | - | [58, 79, 114] |

* HIV stages are defined as acute HIV ($s$ = 1), ART-naïve with CD4 ≥ 350 ($s$ = 2), ART-naïve with CD4 200-349 ($s$ = 3), ART-naïve with CD4 < 200 ($s$ = 4), on ART ($s$ = 5) or interrupting ART ($s$ = 6).

Studies conducted in sub-Saharan Africa have found conflicting evidence regarding the relationship between male socio-economic status and reporting of multiple sexual partners. DHSs mostly find positive associations between household wealth and male reporting of multiple partners [115-117], but not in all countries [117]. DHS data also suggest a positive association between male employment and reporting of multiple partners, with an adjusted OR of 1.22 (95% CI: 1.12-1.34) across 6 countries [115]. Outside of South Africa, most studies that have assessed the effect of educational attainment on men's reporting of multiple partners have found non-significant associations [80, 115, 118], but some have suggested a positive relationship [116, 119, 120]. Mmbaga *et al* [121] found that in Tanzania, the relationship between education and male reporting of multiple/concurrent partners changed over time, from strongly positive in the early stages of the HIV epidemic, to negative later in the epidemic. Randomized trials of economic interventions have also found mixed evidence. Schaefer *et al* [75] found that in Zimbabwe the receipt of cash transfers was associated with increased reporting of multiple partners by younger men (aged <30) but not by older men. Kohler and Thornton [122] found that in Malawi, men who had recently received a cash transfer were more likely to report recent sexual activity – although it is difficult to determine whether this represents an increase in the number of partners. In summary, there is some evidence of a positive relationship between men's income/employment and their reporting of multiple partners, but evidence regarding the relationship between education and multiple partners is less clear.

South African data suggest broadly similar conclusions. Okafor *et al* [114] found that educational attainment was negatively related to male reporting of multiple partners, while there was a non-linear relationship between income and multiple partners, with reporting of multiple partners being lowest in the lowest income groups and similarly high at incomes of more than R1000 per month (OR 1.70 relative to the lowest income group). Hargreaves *et al* [123] found no significant association between male reporting of multiple partners and either household income or education. Dinkelman *et al* [58] found that in young men, higher educational attainment was strongly positively associated with reporting of multiple partners, but when controlling for education, household income was not significantly associated with multiple partners. In our analysis of the 2016 South African DHS data [79], we found that male reporting of multiple partners was significantly associated with employment and higher education (Table S23) but the effect of household wealth was not significant (results not shown). We have chosen to model only an effect of employment on male rates of partnership formation. We represent the uncertainty in the $Z$ parameter using a gamma hurdle distribution, with a weight of 0.25 assigned to a value of 0 (no effect) and a weight of 0.75 assigned to a gamma distribution with mean 0.43 (the average of the three studies that most directly measured the effect of male employment/income [79, 114, 115]) and standard deviation of 0.25.

Table S23: Predictors of reporting multiple partners in the last 12 months (aOR, 95% CI)

| | Males | Females |
|---|---|---|
| Age | 1.14 (1.08-1.20) | 1.36 (1.24-1.48) |
| Age-squared | 0.998 (0.997-0.999) | 0.995 (0.993-0.996) |
| Married | 0.59 (0.46-0.76) | 0.40 (0.30-0.53) |
| Urban | 0.74 (0.61-0.89) | - |
| Employed | 1.37 (1.11-1.70) | 1.47 (1.17-1.85) |

Source: 2016 DHS (author’s own calculations)

For women, evidence of a relationship between socio-economic status and multiple partnerships in Africa is not consistent. Although some studies find evidence of a negative association [59, 119, 124], other studies find positive associations [58, 118], and most find that the association is non-significant after controlling for other factors [80, 117, 123]. As in men, Mmbaga *et al* [121] found in Tanzania that the association between educational attainment and women’s reporting of multiple partnerships changed from being positive in 1991 to negative in 2005. Randomized trials of economic strengthening have not detected significant effects on women’s reporting of multiple partners [73, 75]. In our analysis of the 2016 South African DHS data, we found that female reporting of multiple partners was significantly associated with being employed (aOR 1.47, 95% CI: 1.17-1.85), but was not significantly associated with educational attainment or household wealth (Table S23). Given the inconsistency of the evidence, we do not model any effect of socio-economic status on women’s entry into short-term relationships.

##### 1.4.5 Condom use

We model condom use as occurring either consistently (in all sex acts) or not at all, at the partnership level, but allow for changes in condom use at specific times (e.g. when a couple starts cohabiting, or when an HIV-positive partner discloses their HIV status). The parameter $\gamma_{2,h,c,b}(x,t)$ represents the probability that condoms are used consistently, at the time of entering a new short-term relationship, for a woman aged $x$ in year $t$ (counted in years from

1985), with $h$ years of completed education and binge drinking frequency $b$ (in days per week). The model of condom use is the same as described previously [1, 30]:

$$\log\left(\frac{\gamma_{2,h,c,b}(x,t)}{1-\gamma_{2,h,c,b}(x,t)}\right) = \log\left(\frac{\gamma^*}{1-\gamma^*}\right) + \nu(x-15) + \varsigma_h(t) + \beta(h-10) + \eta b$$

where $\gamma^*$ is the 'base' probability of condom use in 1998, $\exp(\nu)$ is the factor by which the odds of condom use reduces per year of age, $\exp(\varsigma_h(t))$ is the odds of using a condom in year $t$, relative to that in 1998, $\exp(\beta)$ is the odds ratio for condom use per additional year of schooling, and $\exp(\eta)$ is the odds ratio for condom use per day of binge drinking. The time function $\varsigma_h(t)$ is further defined as

$$\varsigma_h(t) = \kappa_1 + (\kappa_2 - \kappa_1)\left(1 - 0.5^{\left(t/\left(M\alpha^{h-10}\right)\right)^Q}\right)$$

where $\exp(\kappa_1)$ is the odds ratio of condom use in 1985 relative to that in 1998, $\exp(\kappa_2 - \kappa_1)$ is the odds ratio of the 'ultimate' rate of condom use (after condom promotion and distribution programmes have been fully rolled out) relative to the initial rate in 1985, $M$ is the median time to change in condom use (in years after 1985) in people who have completed 10 years of schooling, $\alpha$ is the factor by which this median reduces per additional year of schooling, and $Q$ is the 'shape parameter' controlling how abruptly/gradually condom use changes over time. The $M$ parameter is calculated such that $\varsigma_h(13) = 0$, since the base rate of condom use ($\gamma^*$) applies in 1998 ($t$ = 13). The default values assigned to the above parameters are summarized in Table S24; a more detailed description of the sources on which they are based is provided elsewhere [1, 30].

Table S24: Female rates of condom use in short-term relationships

| Parameter | Description | Value |
|---|---|---|
| $\gamma^*$ | Baseline condom use (1998) | 0.179 |
| $\nu$ | Log OR for change in condom use per year of age | -0.025 |
| $\exp(\kappa_1)$ | OR for condom use in 1985, relative to 1998 | 0.07 |
| $\exp(\kappa_2)$ | OR for ultimate condom use, relative to 1998 | 5.56 |
| $Q$ | Shape parameter controlling speed of behaviour change | 3.05 |
| $\exp(\eta)$ | OR for condom use, per day of binge drinking (per week) | 0.93 |
| $\exp(\beta)$ | OR for condom use, per year of schooling | 1.05 |
| $\alpha$ | Factor by which median time to behaviour change reduces, per year of schooling | 0.97 |

OR = odds ratio

From the previous description it is apparent that the model allows for educational attainment to affect condom use in two different ways: the overall level of condom use (through the $\beta$ parameter) and the timing of behaviour change (through the $\alpha$ parameter). This means that the effect of education on condom use can change over time. This is in line with 'Diffusion of innovations' theory, which posits that a subset of the population are early adopters of a new health intervention, and the rest of the population eventually follow their example [125]. To the extent that the early adopters are more likely to be well-educated individuals of higher socio-economic status [125], one might expect to see a larger socio-economic difference in the adoption of the new intervention early on than later, once the intervention has been widely adopted. Indeed, evidence of such attenuations in the socio-economic gradient of condom use have been found in countries such as Burkina Faso [126] and Brazil [127]. We can roughly estimate the $\alpha$ parameter from the study of Adair [126], who analysed DHS data on men's condom use in non-marital relationships in five different African countries, in each case comparing the effect of educational attainment on condom use in DHSs conducted approximately five years apart. In countries such as Cameroon, in which there was a low initial rate of condom use, the socioeconomic gradient increased over the 5-year period, while in countries such as Burkina Faso, with a high initial condom use, the socioeconomic gradient declined. Figure S9 shows the results obtained from fitting the above model to the data from the Adair study: approximate consistency is achieved when we set $\alpha = 0.96$. When fitting our model to South African condom use data, we find the simulated relationship between condom use and education (over time) is most consistent with the observed relationship when values of $\alpha$ are between 0.96 and 1.

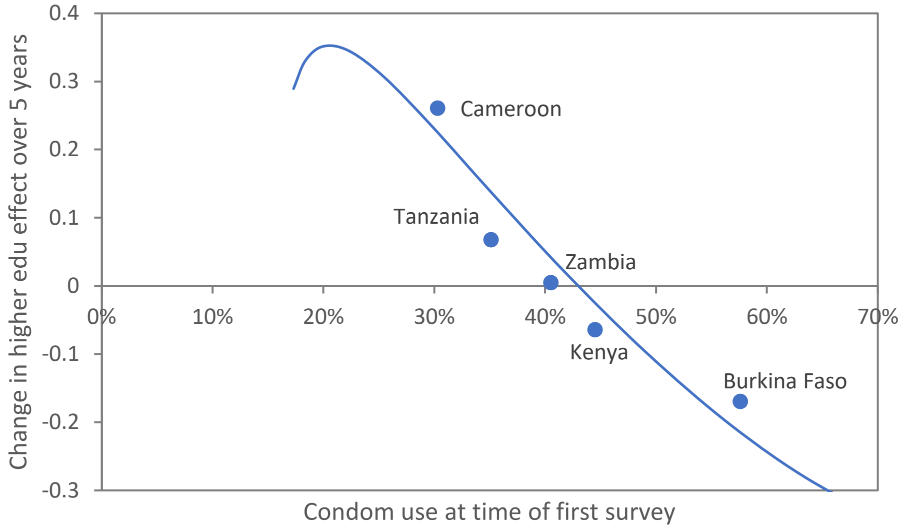

Figure S9: Changes in the relationship between educational attainment and condom use over time

Dots represent estimates based on data presented by Adair [126]. The change in higher education effect (y axis) is the change in the coefficient from an ordinal logistic regression model, on a logit scale, that relates condom use at last sex to an educational attainment categorical variable (with 4 levels), the change being assessed between successive DHSs, five years apart. For the purpose of fitting a model (solid line), we set $\kappa_1$ and $\kappa_2$ to -1.66 and 2.20 respectively (based on the data presented by Adair [126]), and $Q = 3.25$ and $M = 14$ (based on values previously estimated in fitting the MicroCOSM model [1]). For the purpose of estimating $\alpha$, we assume each change in education category (from the ordinal logistic regression model) is equivalent to a 3-year change in completed years of schooling.

Studies conducted in sub-Saharan Africa have generally found a positive association between educational attainment and condom use in non-marital relationships, both in men [80, 118, 120, 126] and women [80, 118-120, 128]. South African studies are generally consistent with this [129-132], and the results from the 1998 and 2003 South African DHSs suggest values of $\beta$ around 0.15. However, some of this 'effect' may be due to the diffusion of innovations dynamic described previously (i.e. more educated people may be quicker to adopt condoms in the early stages of the South African condom promotion campaigns, but aren't necessarily more likely to use condoms in the longer term). In addition, RCT evidence of an effect of education on condom use is generally lacking. The only RCT of an education support intervention that was found to increase condom use or reduce unprotected sex was a trial that involved a substantial cash transfer, and this trial had no significant impact on levels of school dropout (suggesting that the impact on unprotected sex was not mediated by any increase in education) [73].

Because the $\alpha$ and $\beta$ parameters are difficult to set independently of one another, our approach is to assign a prior distribution to represent the uncertainty in $\beta$ and then determine the $\alpha$ value that we consider most plausible conditional upon $\beta$. To do this we run the MicroCOSM model 50 times using different randomly sampled values of $\alpha$ (between 0.9 and 1) and $\exp(\beta)$ (between 1 and 1.25), and for each simulation we calculate the ratio of condom use in non-marital relationships in the highest education category (tertiary education) to that in the lowest category (no education), for each of three years (1998, 2003 and 2016). These simulated ratios are compared to the actual ratios, as reported in the 1998, 2003 and 2016 DHSs, and a sum of squared differences statistic is calculated, for each of the 50 parameter combinations. We fit regression models to these results to smooth out the stochastic variation and identify the combinations of $\alpha$ and $\exp(\beta)$ parameters that give the lowest sum of squared differences statistics. The plausible range of $\alpha$ values is 0.96-1 (as noted previously), and the plausible range of $\exp(\beta)$ values is 1-1.22. For a given value of $\exp(\beta)$, the value of $\alpha$ that appears to minimize the sum of squares statistic is approximately

$$\alpha \approx \left(\frac{\exp(\beta)}{1.165}\right)^{1/3.5}$$

We represent the uncertainty in the $\beta$ parameter using a gamma hurdle distribution with a probability of 0.5 assigned to a value of zero (no long-term effect of education on condom use) and the remaining probability assigned to a gamma distribution with mean 0.10 and standard deviation 0.05. The probability mass at 0 corresponds to the lack of high-quality RCT evidence of a true effect of education on condom use, while the gamma mean and standard deviation were chosen to yield a range of $\exp(\beta)$ values within the plausible range of 1-1.22 noted previously, based on model fits to DHS data. For any $\beta$ value sampled from this prior distribution, we determine the $\alpha$ value using the above equation.

Men's average age-specific rates of condom use in short-term relationships are calculated as weighted averages of the age-specific rates in females (weighting the female rates by the proportions of partners in each age group) [1]. Rates of condom use are further adjusted to take into account inter-individual variation in condom preference, racial differences in condom use, the effect of disclosure of HIV status on condom use, and changes in condom use when short-term partnerships transition to long-term, as described previously [1]. In men, rates of condom use are also adjusted to take account of the man's Gender Equitable Men's score [30]. The odds of condom use in casual sex is assumed to be 2.2 times that in short-term partnerships [129, 133]. A similar approach is also followed in specifying the probability of condom use in sex worker-client interactions, but with $\gamma^*$ adjusted by an odds ratio of 13.17 (to reflect the higher rate of condom use in sex worker-client relationships), $\exp(\kappa_1)$ set to 0.17, $\exp(\kappa_2)$ set to 6.29 and $Q$ set to 4.00.

##### 1.4.6 Marriage/cohabitation

In our model, the term 'marital relationship' includes relationships that are cohabiting. Our model of rates of marriage and divorce is similar to that in the Thembisa model [43]. In the Thembisa model, age-specific rates of marriage, in men and women, are calculated on the assumption that the time to first marriage (after age 16) follows a log-logistic distribution. For an individual of sex $g$ who was born in year $t$ (measured in years after 1985), the probability that they have never married by age $x$ is

$$S_g(x,t) = \frac{1}{1 + \left((x-16)\exp(-C_g - B_g t)\right)^{1/\gamma_g}}$$

The scale parameter of the log-logistic distribution ($C_g + B_g t$) changes for successive birth cohorts, so that the age at first marriage increases over time. Age-specific rates of divorce, in contrast, decrease slightly over time. The model assumes that in the first 12 months after divorce or widowhood, men have a significantly higher rate of remarriage, but thereafter the annual rate of marriage is the same as for other men of the same age who have never previously been married. The Thembisa model has been fitted to age- and sex-specific data on the prevalence of marriage (or cohabitation) from the 1996 and 2001 census and the 2006 and 2016 community surveys.

Although the model of marriage in MicroCOSM is structurally similar to that in Thembisa, being based on log-logistic distributions, there are a number of modifications. Firstly, the constant scale parameters of the log-logistic distribution are specified separately for each race group $r$ ($C_{g,r}$), to account for racial differences in the prevalence of marriage. Secondly, we include adaptations to the marriage rates to account for differences in marriage rates depending on educational attainment and current schooling. This means that if the probability of marriage at age $x$ in the 'baseline' category (people out of school who have not completed high school) is

$$p_{g,r}(x,t) = 1 - \frac{S_{g,r}(x+1,t)}{S_{g,r}(x,t)}$$

then the adjusted odds of marriage in people with educational attainment $h$ and current schooling status $j$ is

$$\log\left(\frac{p_{g,r,h,j}(x,t)}{1 - p_{g,r,h,j}(x,t)}\right) = \log\left(\frac{p_{g,r}(x,t)}{1 - p_{g,r}(x,t)}\right) + \beta_{g,h} + \delta_j + \alpha_{g,r}(x)$$

where $\beta_{g,h}$ and $\delta_j$ represent the effects of education and current schooling respectively, and $\alpha_{g,r}(x)$ represents an age-race interaction (described below). Lastly, the age-specific marriage rates are modified so that they apply only to unmarried people who are currently in short-term relationships (whereas the Thembisa rates apply to all unmarried people who are sexually experienced).

The MicroCOSM model also differs from the Thembisa model in that we use a logistic regression model, fitted to data from the 1993 October Household Survey, to determine the probabilities of being married at the start of the simulation (in 1985), and use these probabilities to randomly assign a marital status to everyone in the simulated population at the start of the simulation. Table S25 shows the results from the multivariable logistic regression model, fitted to the 1993 October Household Survey. A cubic function has been used to represent the relationship between age and marital status. In addition, we allow for an interaction between age and employment status, with the log odds of marriage in employed individuals increasing by $\beta$ for each year of age below age 65 (on the assumption that employment status has more effect on the odds of marriage at young ages than at older ages, and that employment status is unlikely to influence marriage after retirement age). Results suggest a strong positive association between employment and marriage in men, but a negative relationship between employment and marriage in women. The results also suggest that the prevalence of marriage is highest in individuals with education to grade 2 or less.

Table S25: Multivariable analysis of factors associated with marriage in 1993

| | Males | Females |
|---|---|---|
| Per unit increase in (age – 15) | 1.56 (1.54-1.59) | 1.52 (1.51-1.54) |
| Per unit increase in $(age – 15)^2$ | 0.9915 (0.9909-0.9921) | 0.9894 (0.9890-0.9898) |
| Per unit increase in $(age – 15)^3$ | 1.000050 (1.000045-1.000056) | 1.000073 (1.000069-1.000076) |
| Race | | |
| Black African | 1 | 1 |
| ‘Coloured’ | 1.50 (1.39-1.63) | 1.38 (1.30-1.47) |
| White | 2.56 (2.37-2.77) | 2.94 (2.77-3.12) |
| Education | | |
| None/grades 1-2 | 1 | 1 |
| Grades 3-7 | 0.94 (0.86-1.04) | 0.79 (0.74-0.85) |
| Grades 8-11 | 0.89 (0.81-0.98) | 0.64 (0.60-0.69) |
| Grade 12 | 0.84 (0.75-0.93) | 0.75 (0.69-0.81) |
| Tertiary | 0.83 (0.70-0.98) | 0.64 (0.55-0.74) |
| Employed | 1.64 (1.42-1.89) | 0.43 (0.38-0.48) |
| Per unit decrease in age below 65, if employed | 1.031 (1.026-1.036) | 1.018 (1.014-1.022) |
| Constant | 0.0020 (0.0017-0.0024) | 0.0260 (0.0234-0.0290) |

Source: 1993 October Household Survey (author’s own calculations)

With the exception of the constant term, all the coefficients in Table S25 are used in MicroCOSM to assign initial marital status. The constant terms are increased (to 0.0036 in men and 0.0450 in women) because the prevalence of marriage has been steadily decreasing over time, and the model assumptions apply in 1985, whereas the regression model is applied to 1993 data.

Table S26 summarizes the changes to the default parameters in the Thembisa model. The log-logistic scale parameters have been adjusted to match the age-specific prevalence of marriage in the 2001 census (it is worth noting that a higher scale parameter implies a *lower* rate of marriage). Although the standard log-logistic shape parameters in Thembisa produce a consistent age pattern when compared with that in Africans, the age pattern is not consistent with that observed in ‘coloured’ and white South Africans, and we therefore apply a few age-specific adjustments ($\alpha_{g,r}(x)$) in these two race groups in order to bring the modelled prevalence of marriage in these two race groups more in line with the age-specific prevalence levels in the 2001 census (Figure S10).

Table S26: Effects of race, educational attainment and same-sex relationship on marriage rates

| Parameter | Males | Females |
|---|---|---|
| Log-logistic constant scale parameter ($C_g$) | | |
| Black African | 3.30 | 2.90 |
| 'Coloured' | 2.80 | 2.40 |
| White | 1.30 | 0.90 |
| Change to scale parameter per year increase in age at birth ($B_g$) | 0.040 | 0.051 |
| Effect of educational attainment (OR, $\exp(\beta_{g,h})$) | | |
| Completed grade 12 | 1.50 | 0.70 |
| Tertiary education | 3.50 | 0.70 |
| Effect of currently being in school (OR, $\exp(\delta_j)$) | 0.30 | 0.30 |
| Whites: aged <25 (OR, $\exp(\alpha_{g,r}(x))$) | 0.50 | 0.50 |
| Whites: aged 25-34 (OR, $\exp(\alpha_{g,r}(x))$) | 2.00 | 2.00 |
| 'Coloureds': aged 45+ (OR, $\exp(\alpha_{g,r}(x))$) | 0.50 | 1.00 |
| Effect of being in a same-sex relationship (RR) | 0.48 | - |

Adjustment to annual probabilities of marriage are specified either as odds ratio (OR) adjustments or as relative risk (RR) adjustments.

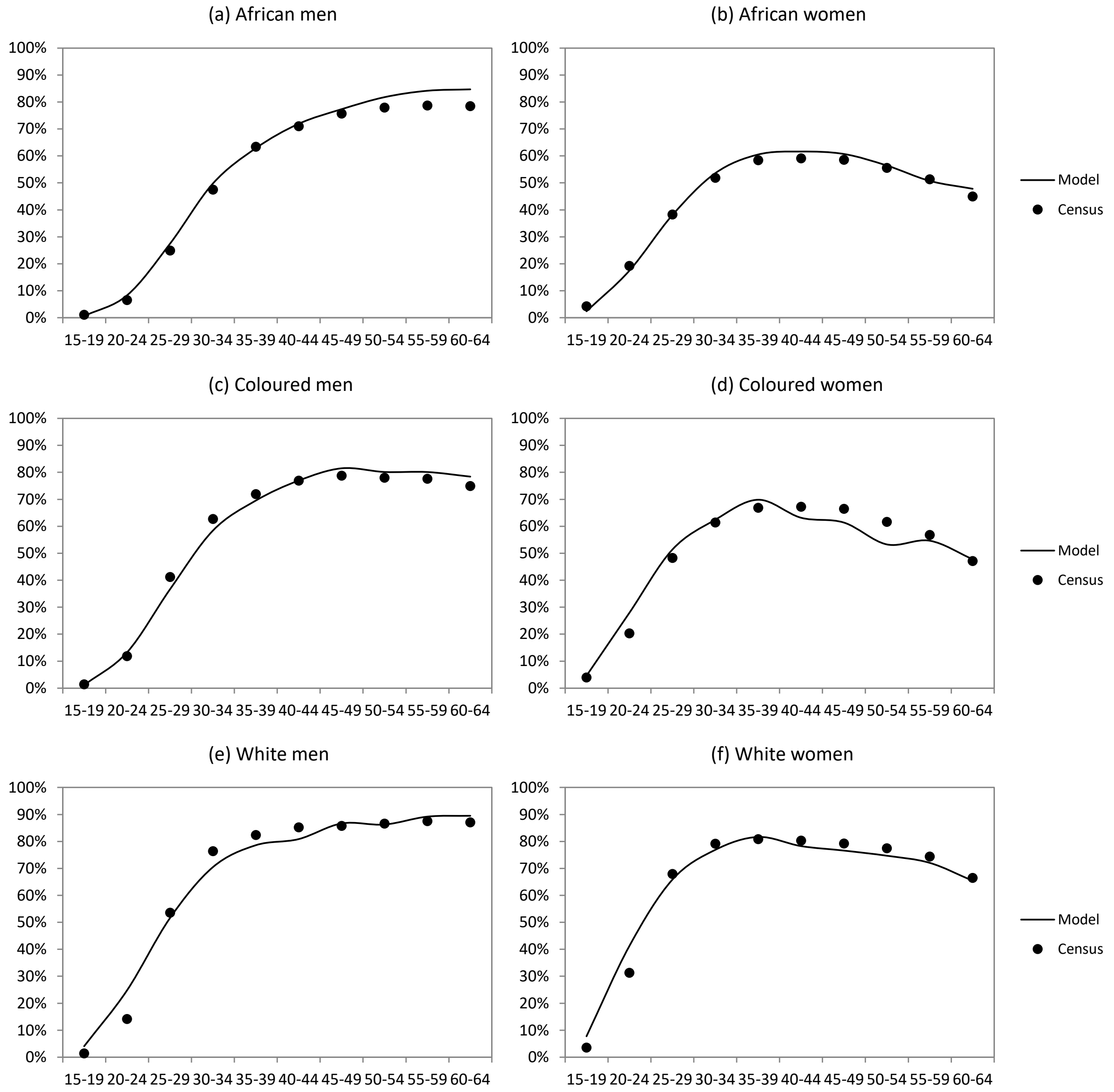

Figure S10: Fraction of individuals who are in marital/cohabiting relationships in 2001

The change in scale parameter per year of increase in age at birth ($B_g$) is set in such a way that the modelled trend in the fraction of the population that is married is consistent with that observed in the censuses and national surveys (Figure S11). The assumed annual changes in scale parameter are more substantial than assumed in Thembisa (0.040 in men and 0.051 in women, compared to 0.017 and 0.020 respectively in Thembisa). The modelled difference in the prevalence of marriage, between men and women, is slightly greater than that observed in the data. This is partly because our model does not allow for polygamy, which – although not common in South Africa – could account for the modelled male prevalence of marriage being slightly exaggerated. Alternatively, there may be reporting biases affecting the census and survey data (for example, the male partner in a relationship might be less likely to describe the relationship as "living together" than the female partner).

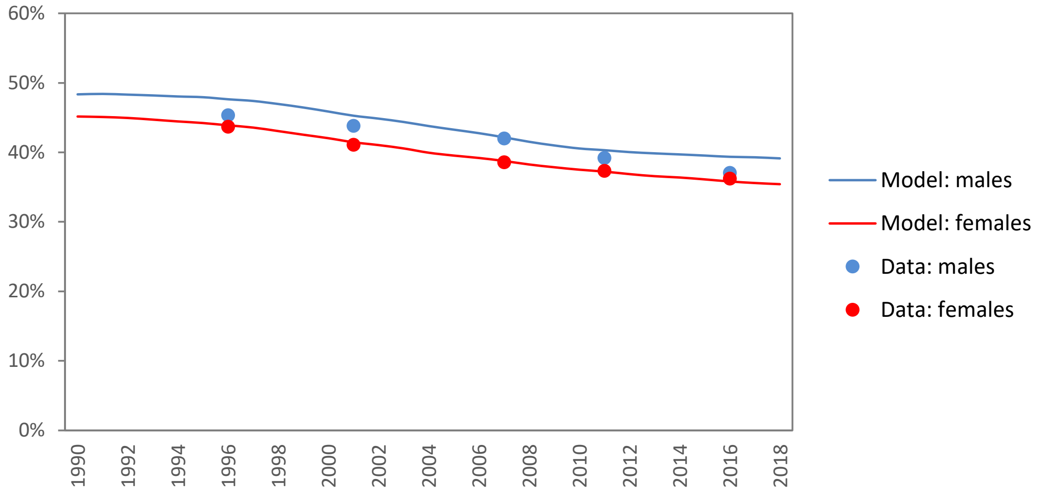

Figure S11: Proportion of the population aged 15 and older that is married/cohabiting
Data are from the 1996 and 2001 census, the 2007 and 2016 community surveys and the 2011 General Household Survey (GHS) [134]. Data from the 2011 GHS have been used in place of the 2011 census data due to concerns regarding the reliability of the 2011 census data on marital status.

The assumptions about the effect of socio-economic status on marriage are set so that the model matches (approximately) the relationship between educational attainment and marital status in the 2011 General Household Survey [134] (Figure S12). Both in men and in women there is a high proportion married among those with tertiary education, a pattern that is consistent with the 2001 census (results not shown). Marriage rates are assumed to be influenced principally by educational attainment, and it is also assumed that individuals who are currently at school/university have a lower rate of marriage (independent of their educational attainment). It may seem more natural to assume that rates of marriage depend on employment status. In men, for example, Table S25 shows that employment is more strongly associated with marriage than educational attainment. However, association does not imply causation, and it is the causal relationship in which we are principally interested. In women, the negative association between marriage and employment probably reflects an effect of marriage on employment (married women may have less need to work if their husband is employed) rather than an effect of employment on marriage. Another reason for preferring to model an education effect over an employment effect is that it is difficult to match the pattern observed in Figure S10 if the socioeconomic effect is assumed to operate only through employment status. It is worth noting that in women the completion of secondary education is assumed to be associated with a slightly reduced *incidence* of marriage, consistent with data from other African studies [80, 135, 136]. However, the more educated women nevertheless end up having a higher *prevalence* of marriage in Figure S10 because higher educational attainment is associated with many of the factors that are positively associated with marriage (e.g. higher partner educational attainment, white race).

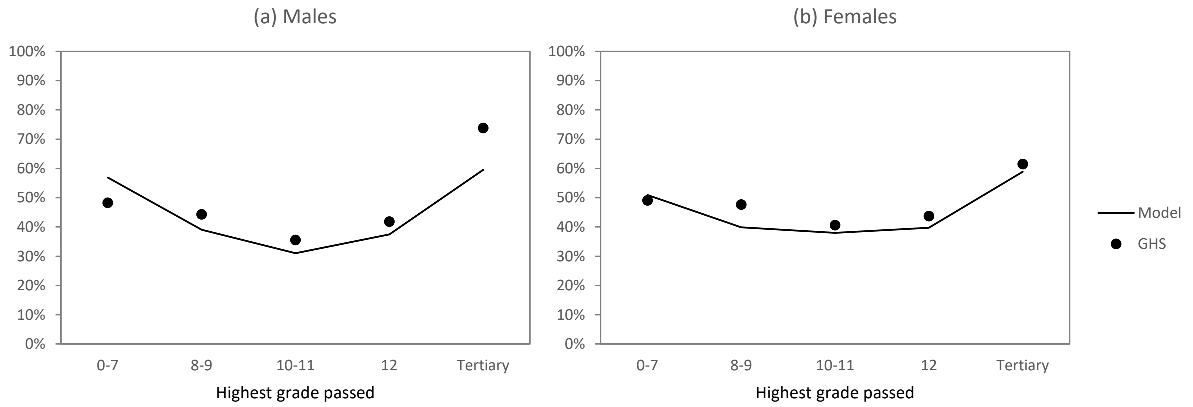

Figure S12: Fraction of 20-59 year olds who are married/cohabiting in 2011

Data from RCTs of school support interventions show that these interventions tend to reduce rates of entry into marriage in both girls and boys, with reductions in marriage often being similar to reductions in school dropout [67, 70, 71]. This strongly suggests that being in school is associated with a reduced rate of entry into marriage. When re-analysing the 1993 October Household Survey (Table S25) and including a term to represent the effect of currently being in school/university, we found that being in school/university reduced the odds of marriage substantially (adjusted OR of 0.44 [95% CI: 0.36-0.54] in males and 0.17 [95% CI: 0.15-0.19] in females), with modest attenuation of the previously estimated effects of educational attainment. Based on these survey data and the RCT estimates, we represent the uncertainty in the relative odds of entry into marriage if currently in school/tertiary education using a beta hurdle distribution, with a probability of 0.1 assigned to an OR of 1 (no effect) and the remaining 0.9 probability being assigned to a beta distribution with mean of 0.3 and standard deviation 0.2.

We cannot rule out the possibility that employment status does directly affect the rate of marriage, and it may seem plausible that it does, given that unemployed men are less likely to have the financial means to pay *lobola*. However, employment status is less stable over time than educational attainment, and this makes it difficult to infer causality when observing an association between employment and marriage. Further work is required to better understand the specific socio-economic factors that determine entry into marriage. It is also possible that socio-economic factors may influence rates of divorce [137], but such an effect is not currently allowed for in our model.

Finally, men in same-sex relationships are assumed to have lower rates of marriage than men of the same age in heterosexual relationships. The assumed relative rate of marriage in same-sex relationships is based on a previous analysis in which MicroCOSM was fitted to data on sexual behaviour in South African men who have sex with men [138].

#### 1.5 Other socio-economic effects

##### 1.5.1 Effects on hormonal contraceptive use

The model assumptions about contraceptive uptake and discontinuation have been described in detail previously [1]. Briefly, the model considers four forms of contraception: condoms (already described in section 1.4.5), injectable contraceptives, oral contraceptives and female sterilization. Female sterilization is assumed to be irreversible; rates at which women become sterilized differ by age and race, but not by educational attainment or other measures of socio-economic status.

'Hormonal contraception' here refers to injectable and oral contraception. The initiation of hormonal contraception is modelled as occurring at a number of possible time points: at the start of a new relationship, on discontinuation of condom use within an existing relationship, on entry into commercial sex, after birth, or at other times:

- In the case of hormonal contraceptive initiation at the start of a new relationship, the probability depends on age, race, educational attainment, previous pregnancy history, fecundability and condom use. For each additional grade passed, the odds of initiating hormonal contraception is assumed to increase by a factor of 1.15; this multiple was chosen such that the model matched the reported patterns of contraceptive use by educational attainment in the 1998 DHS [139].
- The probability of initiating hormonal contraception at the time of discontinuing condom use is calculated as a function of the previous probability, adjusted to take into account the relative rates of hormonal contraceptive use in condom users relative to non-users.
- The probability of initiating hormonal contraception at the time of becoming a sex worker is the same as that calculated at the time of starting a new relationship, except that no adjustment for condom use is made (on the assumption that sex workers would use condoms primarily for HIV/STI prevention rather than pregnancy prevention).
- The probability of initiating hormonal contraceptive use after birth (or weaning if the mother breastfeeds) depends on age, educational attainment, fecundability and current relationship status. The education adjustments are the same as for the probability of initiating hormonal contraception at the time of starting a new relationship, based on the findings of a KwaZulu-Natal study that assessed uptake of dual protection (condoms and non-barrier methods) by 14 weeks postpartum [140].
- The annual probability of initiating hormonal contraception for other reasons is very low, and depends only on age.

Among women who initiate hormonal contraception, the probability of choosing injectable over oral contraception is assumed to depend on previous contraceptive method choice (if the women used hormonal contraception previously), previous birth, educational attainment and race. The odds of using injectable contraception (over oral) are multiplied by a factor of 0.95 for each additional grade completed, consistent with the range of ORs (0.90-0.98) estimated by Burgard *et al* [141].

Contraception is assumed to reduce the risk of falling pregnant by 78% in the case of condoms and hormonal contraceptive use, by 90% in the case of injectable contraceptives and by 99.5% in the case of female sterilization.

Women are assumed to discontinue hormonal contraception if they fall pregnant (i.e. when experiencing contraceptive failure), after ceasing sexual activity (i.e. if they are no longer in a relationship), after sterilization, or for other reasons (these other reasons are not modelled, but might include factors such as desiring children or health concerns). Rates of discontinuing hormonal contraception are assumed not to depend on socio-economic status.

##### 1.5.2 Effect of socio-economic status on male circumcision

Several African studies suggest that male circumcision is positively associated with socio-economic status. Some studies suggest an effect of socio-economic status on rates of traditional male circumcision, the predominant form of circumcision in African countries prior to the promotion of voluntary medical male circumcision (VMMC) as an HIV prevention strategy. For example, in a review of African DHSs conducted prior to the promotion of VMMC, Lau *et al* [142] found male circumcision was positively associated with wealth and there was a 'U-shaped' relationship between educational attainment and circumcision. In another review of the same surveys, Tram *et al* [143] found the relationship between socio-economic status and male circumcision was inconsistent across countries – for example, the relationship with educational attainment was strongly negative in Lesotho but strongly positive in Mozambique. The high cost of traditional male circumcision is sometimes cited as a barrier [144], and could be an explanation for the positive association between socio-economic status and circumcision in some settings.

There is more uniform evidence of a positive association between socio-economic status and VMMC. In a more recent review of African surveys conducted between 2010 and 2018 (i.e. after the start of VMMC promotion programmes), Hamidouche *et al* [145] found that men in the highest wealth quintile were on average twice as likely to be medically circumcised as men in the lowest wealth quintile. The 2020 Lesotho PHIA survey found a more dramatic 3.5-fold difference in proportions circumcised when comparing the richest to the poorest wealth quintiles [146]. In a South African community surveyed after the rollout of VMMC, male circumcision levels were significantly higher in males who had completed their secondary education [147]. A RCT of a school support intervention in Kenya found that boys who received material support (payment of school fees and uniforms) were significantly more likely to get circumcised [69], an outcome that the authors attributed to the promotion of VMMC in school settings. Better knowledge of the health benefits of VMMC among men of higher socio-economic status could be an explanation for the greater uptake of VMMC.

Given that the evidence of an association between traditional male circumcision and socio-economic status is unclear, we model only an effect of socio-economic status on VMMC uptake. In South Africa, most VMMC occurs in adolescent males who have not yet completed their education [148], and it is therefore most practical to model an effect of per-capita household income on uptake of VMMC (rather than an effect of educational attainment). We define the parameter $C$ to be the log of the relative rate of medical male circumcision per unit increase in the log of the per capita household income, i.e. a value of 0 corresponds to no effect of income on the rate of medical male circumcision. We define $H$ as the relative rate of per capita household income per income decile, and $m$ as the relative rate of medical male circumcision per income decile. This means that if $B$ is the average income in the poorest income decile, the average income in decile $n$ is $BH^{n-1}$, or equivalently, the log of the per capita income in decile $n$ is $\log(B) + (n-1)\log(H)$. This in turn means that

$$C = \frac{\log(m)}{\log(H)}.$$

We can estimate the $m$ and $B$ parameters from Lesotho data. If $F$ is the probability of not being medically circumcised in the poorest income decile, then the probability of not being medically circumcised in decile $n$ is $Fm^{1-n}$, so that the relative probability of being uncircumcised per quintile is $m^{-2}$. Based on the data in the 2020 Lesotho PHIA [146], we estimate that $m$ is approximately 1.14. It can further be shown[2] that if the Gini coefficient for a country is $G$, then

$$G = 100\left(1 - 2\left(\frac{1}{log(H^{10})} - \frac{1}{H^{10} - 1}\right)\right)$$

Lesotho had a Gini coefficient of 44.6 in 2017 [149], from which it follows that $H$ = 1.36. Substituting these values of $m$ and $H$ into the equation for $C$ gives a $C$ estimate of 0.42. This is likely to be an upper bound on the parameter we might expect in South Africa, for two reasons. Firstly, as noted previously, the income gradient in medical male circumcision in Lesotho is substantially greater than the average estimated for the southern and eastern African region [145]. Secondly, the calculation does not take into account the competing risk of traditional circumcision, which is greater in lower wealth quintiles in Lesotho [146]. To represent the uncertainty in the $C$ parameter, we therefore assign a gamma prior distribution with a mean of 0.2 and a standard deviation of 0.1 (which has a 97.5 percentile of 0.44, close to the upper bound estimated from Lesotho data).

##### 1.5.3 Effects on HIV testing

The model assumptions about HIV testing have been described in detail previously [2]. Briefly, we model eight *current* HIV testing modalities: testing in antenatal clinics, testing in patients with HIV-related opportunistic infections, testing in STI patients, testing in PrEP users, testing in males who seek medical circumcision, testing in prisons, testing in partners of newly diagnosed individuals (‘passive referral’) and testing for other reasons (mostly people who independently seek HIV testing). Of these testing modalities, the last is the most significant (accounting for roughly 63% of adult HIV tests over the 2010-2015 period [2]), followed by the STI, OI and ANC modalities (11%, 10% and 7% respectively). We focus here on the testing for other reasons (which we refer to here as ‘general testing’), as the rates of testing for the first seven modalities are not assumed to depend on socio-economic status (although socio-economic status does affect factors such as incarceration and fertility).

Rates of general testing are assumed to change over time, with the rates being set such that the model matches total numbers of HIV tests performed in South Africa. These general testing rates further depend on age, sex, educational attainment, prior HIV testing history (and knowledge of HIV status/receipt of ART) and sexual experience. For each additional grade completed, the rate at which individuals seek general HIV testing is assumed to increase by a factor of 1.12. (For example, the rate of HIV testing in someone who has completed grade 8 would be 0.71 times ($1.12^{-3}$) the base rate that applies in an individual who has completed grade 11.) The factor of 1.12 is the estimated effect of educational attainment on the uptake of HIV testing by black South Africans between 2010 and 2012, after controlling for age, sex and other factors, which was estimated using data from nationally-representative surveys [150]. This is

[2] Suppose we want to estimate the total income earned by the poorest proportion $x$ of the population. This is $\int_0^x BH^{10x}dt = \frac{B}{\log(H^{10})}(H^{10x} - 1)$, where $B$ is the lowest income. Expressed as a fraction of all income earned, this is $\frac{H^{10x}-1}{H^{10}-1}$. This means that the integral of the area below the Lorenz curve is $L = \int_0^1 \frac{H^{10x}-1}{H^{10}-1}dx = \frac{1}{log(H^{10})} - \frac{1}{H^{10}-1}$. The Gini coefficient is the proportion of the area below the line of equality that lies above the Lorenz curve, i.e. $\frac{0.5-L}{0.5} = 1 - 2L$, expressed as a percentage (0-100).

consistent with data from other African countries, which also show substantially higher rates of HIV testing in more educated individuals [151].

#### 1.6 Modelling socio-economic interventions

We consider three intervention types. The first is vocational training interventions, which may include microfinance interventions (which are typically intended to assist people in establishing their own businesses). In a systematic review and meta-analysis of vocational training programmes in low- and middle-income countries, Tripney *et al* [152] estimated that these programmes increased the employment probability by an average of 0.134 standard deviations (95% CI: 0.024-0.243). In our model, we wish to express the intervention effect as an odds ratio, comparing the odds of unemployment in those participating in the intervention to the odds in people who did not participate in the intervention. If $p$ is the probability of unemployment in the control group, the standard deviation is $\sqrt{p(1-p)}$, and so if the intervention increases the employment probability by 0.134 standard deviations, the relative risk of unemployment in the intervention group is

$$\frac{p-0.134\sqrt{p(1-p)}}{p} = 1 - 0.134\sqrt{\frac{1-p}{p}}$$

As a first approximation to the unemployment odds ratio, we might consider a value of 0.866 (i.e., 1 – 0.134). This could be an underestimate because the $\sqrt{(1-p)/p}$, term is generally < 1 when there is a high baseline probability of unemployment in the trial participants. On the other hand, it could be an over-estimate because 0.866 is calculated as a relative risk, and we would expect the corresponding odds ratio to be smaller (assuming the odds ratio is <1). To represent the uncertainty in the odds ratio, we assign a beta prior distribution with a mean of 0.87 and a standard deviation of 0.10. This prior distribution has 2.5 and 97.5 percentiles of 0.62 and 0.99 respectively. The lower limit is roughly consistent with the Tripney *et al* meta-analysis [152]: the greatest effect measured by any intervention was a 0.37 standard deviation difference in employment, which corresponds to an unemployment odds ratio of 0.63 when using our crude approximation.

RCTs of vocational training programmes have mostly assessed employment outcomes over the short term, and it is not clear if the effects of vocational training programmes are sustained over the longer term. For example, if an individual has not found work within 12 months after completing a vocational training programme, should we assume that they are still more likely to find employment in the next 12 months than unemployed individuals who never received vocational training? For the purpose of model calibration, we assume that this is the case.

The second type of intervention that we consider is cash transfer interventions. For the purpose of this analysis, we do not distinguish between conditional and unconditional transfers, as there is little strong evidence to suggest that conditionality affects behavioural outcomes [71]. The value of the cash transfer is assumed to be added to the total household income and divided by the number of individuals in the household (i.e. we assume the cash transfer affects the income of the household primarily). For the sake of model calibration, we assume that households are only eligible to receive cash transfers if the per capita household income (before the transfer) is below the national average. We assume that the effects of cash transfers on sexual risk behaviours are not sustained after these interventions end, in line with medium-term follow-up data after the completion of a major cash transfer intervention [153].

The third type of intervention that we consider is school support interventions, which are intended to improve school attendance and completion, and prevent school dropout. These interventions typically include both a material support component (e.g. providing school uniforms or covering the costs of school fees) and non-material support (e.g. attendance monitoring and counselling). We model these two components separately. We define *S* as the relative rate of school dropout in learners who receive non-material support (compared to learners who receive no support). In a systematic review and meta-analysis of interventions to reduce school dropout , Wilson *et al* [154] estimated that attendance monitoring interventions significantly reduced the odds of school dropout (aOR 0.68, 95% CI: 0.61-0.76); more significant effects were estimated for other types of dropout intervention, but these interventions were generally different from the school support interventions evaluated in African settings. A limitation of this review is that it reported almost exclusively on US data; in addition, no estimate of the heterogeneity in attendance monitoring effects was provided (though overall heterogeneity across all interventions was substantial). Given the relatively weak data to inform the assumed value of *S*, we assign a beta prior with a mean of 0.68 and a standard deviation of 0.18. (This distribution has 2.5 and 97.5 percentiles at 0.29 and 0.96 respectively, i.e. reflecting the large uncertainty in the *S* parameter.)

For the material support component of the school support intervention, we assume that the magnitude of the benefit is proportional to the cash equivalent value of the material support. We define *C* as the multiple by which the probability of school dropout is reduced for a R800 increase in the annual cash equivalent value of the material support (in 2005 rand terms). This means that for a school support intervention that includes both attendance monitoring/counselling and material support values at R*x* per annum, the relative rate of school dropout is

$$SC^{\sqrt{x/800}},$$

compared to that in learners who receive no intervention. In a systematic review and meta-analysis of cash transfer programmes in low- and middle-income countries, García and Saavedra [155] estimated that these interventions reduced the annual probability of secondary school dropout by an average of 2.9% (95% CI: 1.9-3.8%). If the average annual probability of high school dropout is around 20% (as we have previously estimated in South Africa [1], and in line with settings in which high school dropout interventions are typically tested [154]), this 2.9% reduction is equivalent to a relative rate of dropout of 0.86. Although García and Saavedra did not find the impact of the intervention to be proportional to the dollar value of the cash transfer, this may be because they did not make purchasing price parity (PPP) adjustments across countries. Without knowing the PPP and market exchange rates in the countries in which each trial was conducted, it is difficult to define an appropriate comparator for the African setting, but we use R800 as it is close to the median value of South African rand-equivalent values of cash transfers in sub-Saharan African RCTs (see section 2.2). We therefore set the prior distribution for the *C* parameter to have a mean of 0.86 and a standard deviation of 0.10.

In addition, school support interventions are assumed to increase the probability of re-enrolment in school, for those who have dropped out of school. In a trial of cash transfers in Malawi, girls aged 13-22 who were out of school at baseline had a 57% probability of re-enrolling in school after two years [71], equivalent to a 34% probability of re-enrolment per year ($1 – (1 – 0.57)^{0.5}$). Based on this, we assume that the non-material component of the school support is associated with a 0.34 annual probability of return to school, in those who have dropped out.

#### 2. Calibration to randomized controlled trial data

We adopt a Bayesian approach to estimating the parameters in our model and quantifying the uncertainty around the model outputs. Prior distributions are specified to represent the uncertainty around (a) the effects of structural factors on sexual risk behaviour and (b) the effects of structural interventions. Likelihood distributions are calculated to represent the extent of the agreement between the model estimates and estimates from randomized controlled trials (RCTs) of the effectiveness of different structural interventions. The posterior distribution, which represents the set of estimates most consistent with both the prior distributions and the data that define the likelihood calculations, is then approximated using numeric methods. Each of these steps is described in more detail in the sections that follow.

##### 2.1 Prior distributions

We distinguish here between the effects of structural factors on sexual risk behaviours and the effectiveness of structural interventions in changing structural factors. Examples of the former parameters include the effect of male employment on engaging in casual sex, and the effect of education on condom use. As noted in the literature review, it is in many cases difficult to determine whether the observed associations between structural factors and HIV risk behaviours represent a 'true effect' or whether the observed associations are merely due to confounding. The prior distributions therefore need to reflect both (a) the uncertainty about whether there is a true effect, and (b) the uncertainty regarding the size of that effect, assuming it exists. To represent this uncertainty we use hurdle distributions. For example, a gamma hurdle distribution is used to represent the uncertainty around parameter $A_1$, the proportionate increase in men's rate of engaging in casual sex due to employment. This means that

$$\Pr[A_1 = x] = \begin{cases} \theta & \text{if } x = 0 \\ \frac{(1-\theta)\lambda^{\alpha}x^{\alpha-1}}{\exp(\lambda x)\Gamma(\alpha)} & \text{if } x > 0 \end{cases}$$

where $\theta$ is the hurdle parameter, and $\alpha$ and $\lambda$ are the shape and scale parameters of the gamma distribution respectively. The hurdle parameter $\theta$ therefore represents the prior belief in the probability of a null association. For the sake of setting assumed values of $\theta$, we grade the strength of evidence for each of the hypothesized effect parameters into one of three categories:

I. Evidence from observational studies suggests a possible effect, but there are no RCTs confirming/suggesting that an effect exists.
II. There is some evidence from RCTs to suggest that an effect exists, but RCTs are not consistent or the evidence is considered weak (e.g. only one trial, or the trial outcome doesn't correspond exactly to the outcome in our model).
III. There is evidence from at least two RCTs suggesting a significant effect, and the results from different trials are consistent.

We consider only RCTs conducted in sub-Saharan Africa in grading the evidence, as the social determinants of sexual risk behaviour are likely to be different outside of Africa, and we would therefore not expect structural interventions to have the same effects. The hurdle parameter $\theta$

is set to 0.50 for category I effects, 0.25 for category II effects and 0.10 for category III effects. For example, in the case of the effect of income on medical male circumcision, there is observational evidence suggesting an association, but only one randomized trial found significant increases in medical male circumcision following school support with financial assistance [69]. We therefore grade the evidence as 'category II' and assign a 0.25 weight to the probability of no true effect.

We generally use gamma hurdle models for parameters that are defined on the range [0, ∞). However, many of the effect parameters are expressed as relative risks that are bounded on the range [0, 1]. In these cases we use a beta hurdle model, with the hurdle at 1 rather than at 0, since a relative risk of 1 implies no effect. Mathematically, if $\beta$ is the parameter for which a beta hurdle prior is specified,

$$\Pr[\beta = x] = \begin{cases} \theta & \text{if } x = 1 \\ \frac{(1-\theta)x^{m-1}(1-x)^{n-1}}{\mathrm{B}(m,n)} & \text{if } x < 1 \end{cases}$$

where $\theta$ is the hurdle parameter, and *m* and *n* are the parameters of the beta distribution.

Table S27 summarizes the parameters for which we have specified prior distributions in this analysis. These parameters have all been explained in previous sections (referenced in the final column of the table).

Table S27: Prior distributions for socioeconomic effect parameters

| Parameter | Prior type | Evidence grading | Mean* | Standard deviation* | Section |
|---|---|---|---|---|---|
| Increase in rate of debut in females per log reduction in APCHI† (RR – 1) | Gamma hurdle | III | 0.25 | 0.15 | 1.4.1 |
| RR of debut in females if currently in school | Beta hurdle | III | 0.40 | 0.23 | 1.4.1 |
| RR of debut in males if currently in school | Beta hurdle | I | 0.60 | 0.23 | 1.4.1 |
| Increase in casual sex in females, per log reduction in APCHI† (RR – 1) | Gamma hurdle | I | 0.85 | 0.40 | 1.4.2 |
| Increase in casual sex in men who are employed (RR – 1) | Gamma hurdle | I | 0.50 | 0.25 | 1.4.2 |
| Increase in commercial sex in men who are employed (RR – 1) | Gamma hurdle | I | 0.80 | 0.40 | 1.4.3 |
| RR of partner acquisition in men if employed (RR – 1) | Gamma hurdle | II | 0.43 | 0.25 | 1.4.4 |
| Increase in consistent condom use per year of schooling (OR – 1) | Gamma hurdle | I | 0.16 | 0.08 | 1.4.5 |
| RR of marriage if currently in school | Beta hurdle | III | 0.30 | 0.20 | 1.4.6 |
| Increase in medical male circumcision per log increase in APCHI (RR – 1) | Gamma hurdle | II | 0.20 | 0.10 | 1.5.2 |
| RR of school dropout if receiving school support (non-material) | Beta | - | 0.68 | 0.18 | 1.6 |
| RR of dropout per R800 of school support | Beta | - | 0.86 | 0.10 | 1.6 |
| OR of unemployment if receiving vocational training/microfinance | Beta | - | 0.87 | 0.10 | 1.6 |

APCHI = adjusted per capita household income. OR = odds ratio. RR = relative risk.
* In the case of gamma hurdle and beta hurdle distributions, the mean and standard deviation are for the gamma and beta components of the distribution respectively (i.e. ignoring the probability of a null association). † Per unit difference between the natural log of the APCHI and the log of the national average APCHI, for households that have an APCHI below the national average (for those above the average, no income effect is modelled).

#### 2.2 Likelihood function

##### 2.2.1 Study selection

We calibrated the model to data from randomized controlled trials of economic interventions in sub-Saharan Africa. Relevant trials were identified from systematic reviews of cash transfer interventions and interventions to reduce school costs [156, 157], household economic strengthening interventions [158] and economic empowerment interventions in sub-Saharan Africa [159]. Trials were classified as being pure 'cash transfer' interventions (including both conditional and unconditional cash transfers, but without any strong conditioning on school attendance), school support interventions (which typically aimed to promote school retention, often through the provision of financial support, or through cash transfers conditional on school attendance) and vocational training programmes (directed to individuals who were out of school, providing training to improve their employment prospects and/or credit to enable them to establish their own business).

We excluded studies that were not randomized controlled trials. We also excluded studies of savings programmes, if these did not include elements of vocational training [160], and lottery-based interventions [161], in both cases because the hypothesized effect was on future wealth rather than short-term socio-economic improvement. Cash transfer and school support interventions were excluded if it was not possible to establish the value of the cash transfer (or cash equivalent in the case of school support). Interventions were also excluded if there was no behavioural or HIV/STI/pregnancy outcome evaluated [160]. One Ugandan trial was excluded because it combined vocational training with sexual and reproductive health information sessions, and it was not possible to separate the effects of these two interventions [162].

In some cases it was challenging to make a clear distinction between cash transfers and school support interventions. The RCTs of Pettifor *et al* [73] and Abdool Karim *et al* [163] were classified as cash transfer interventions because although both trials evaluated cash transfers conditional on school attendance, the conditioning was relatively weak (in the former, school attendance was unexpectedly high in both trial arms, and in the latter school attendance was only one of a number of criteria that could qualify young girls for the cash transfer). The RCT of Baird *et al* [71] was treated as two different interventions: an unconditional cash transfer intervention and a school support intervention (the conditional cash transfer arm). The RCT of Schaefer *et al* [75] evaluated both conditional and unconditional cash transfers, but because the outcomes were not reported separately for the two groups, and because the conditions in the CCT arm were not strictly enforced, we treated this as a 'pure' cash transfer intervention.

##### 2.2.2 Extraction of data from selected studies

For each of the included trials, we recorded the type of intervention, and for each outcome that was reported in the trial we recorded

- The type of outcome (e.g. school dropout, condom use at last sex, HIV incidence)
- The demographic characteristics (sex and age range, and whether participants were in school or not)
- The type of statistical measure that was used to compare the intervention and control arms (log odds ratio, log relative risk, or absolute change on a log scale)
- The difference in outcome between intervention and control arms, and the standard error associated with the difference

- The follow-up duration to which the outcome measure related (in months)

In the case of cash transfer and school support interventions, we also recorded the annual cash value of the intervention, expressed in 2005 South African rand terms (further details of the currency conversion are discussed below).

We limited our analysis to outcomes that could reasonably be matched to outcomes simulated in our model. These outcomes included:

- Employment
- Monthly earnings (on a log($x$ + 1) scale)
- School dropout
- Male circumcision
- Recent unprotected sex
- Condom use at last sex
- Multiple partners in the last year
- Casual or transactional sex in the last year
- Currently engaging in casual or transactional sex
- Sexual debut
- Any recent sexual activity
- Partner age difference of 5 or more years
- Cumulative incidence of marriage
- Cumulative HIV incidence
- Cumulative HSV-2 incidence
- Cumulative incidence of curable STIs (gonorrhoea, chlamydia or trichomoniasis)
- Cumulative pregnancy incidence (teenagers only)
- HIV prevalence
- HSV-2 prevalence

In some cases we included estimates of intervention effects even if the recorded outcome was not exactly consistent with the model definition, but the definitions were deemed close enough. For example, in the study of Dunbar *et al* [86], the model outcome is any teenage pregnancy, whereas the RCT outcome is any unintended pregnancy. Because MicroCOSM does not determine whether pregnancies are intended or not, we cannot use the same outcome definition, but since most teenage pregnancies are unintended [164], we can assume that there is approximate equivalence. As another example, our model does not include 'transactional sex' per se, but we expect that casual sex relationships would tend to be more transactional in nature [76, 77], and we therefore compare our modelled casual sex outcomes against transactional sex outcomes in trials.

It is also worth noting that in a few cases (specifically incidence of HIV, HSV-2 and teenage pregnancy) we replaced the actual trial duration with a longer trial duration when simulating the modelled trial outcome, because these events are rare outcomes, and using a longer simulated trial duration therefore helps to reduce the stochastic variation in the model outputs.

**2.2.3 Summary of included studies**

Table S28 summarizes the data recorded for each of the included trials. In total 18 RCTs were identified: four of vocational training or microfinance interventions, nine of cash transfer interventions and six of school support interventions (one trial was classified as evaluating both a cash transfer intervention and a school support intervention [71]). A total of 98 outcomes were recorded in these 18 trials.

Table S28: Randomized controlled trials included in model calibration

| Intervention | Study | Outcome | Sex, age | In school? | Cash value (2005 ZAR) | Measure | Timing (months) | Effect | SE |
|---|---|---|---|---|---|---|---|---|---|
| Vocational training and microfinance | Dunbar et al [86] | Employed | F, 16-19 | No | - | log(OR) | 12 | 0.205 | 0.091 |
| | | Any recent sex | F, 16-19 | No | - | log(OR) | 12 | -0.007 | 0.082 |
| | | Current casual sex | F, 16-19 | No | - | log(OR) | 12 | -0.211 | 0.181 |
| | | Unprotected sex with recent partners | F, 16-19 | No | - | log(OR) | 12 | -0.328 | 0.284 |
| | | Cumulative HIV incidence | F, 16-19 | No | - | log(RR) | 24 | -0.062 | 0.535 |
| | | Cumulative HSV-2 incidence | F, 16-19 | No | - | log(RR) | 24 | 0.405 | 0.387 |
| | | Cumulative pregnancy incidence | F, 16-19 | No | - | log(RR) | 24 | -0.478 | 0.252 |
| | Kim et al [165] | Condom use at last sex | F, 18+ | NS | - | log(RR) | 24 | 0.157 | 0.662 |
| | | Employed | F, 18+ | NS | - | log(RR) | 24 | 0.322 | 0.149 |
| | Gibbs et al [166] | Monthly earnings (on log+1 scale) | M, 18-30 | No | - | log change | 12 | 0.480 | 0.436 |
| | | Monthly earnings (on log+1 scale) | M, 18-30 | No | - | log change | 24 | 0.240 | 0.339 |
| | | Monthly earnings (on log+1 scale) | F, 18-30 | No | - | log change | 12 | 0.710 | 0.362 |
| | | Monthly earnings (on log+1 scale) | F, 18-30 | No | - | log change | 24 | 0.980 | 0.301 |
| | | Cumulative casual sex | F, 18-30 | No | - | log(OR) | 12 | 0.315 | 0.215 |
| | | Cumulative casual sex | F, 18-30 | No | - | log(OR) | 24 | -0.211 | 0.215 |
| | Adoho et al [167] | Employed | F, 16-27 | No | - | log(RR) | 13 | 0.389 | 0.046 |
| | | Condom use at last sex | F, 16-27 | No | - | log(RR) | 13 | 0.044 | 0.045 |
| Cash transfers (excluding school support) | Pettifor et al [73] | Cumulative HIV incidence | F, 13-20 | Yes | 2060.6 | log(RR) | 36 | 0.157 | 0.195 |
| | | Cumulative HSV-2 incidence | F, 13-20 | Yes | 2060.6 | log(RR) | 36 | -0.105 | 0.139 |
| | | School dropout | F, 13-20 | Yes | 2060.6 | log(RR) | 36 | -0.105 | 0.157 |
| | | Cumulative pregnancy incidence | F, 13-20 | Yes | 2060.6 | log(RR) | 36 | -0.062 | 0.110 |
| | | Sexual debut | F, 13-20 | Yes | 2060.6 | log(RR) | 36 | -0.083 | 0.083 |
| | | Unprotected sex with recent partners | F, 13-20 | Yes | 2060.6 | log(RR) | 18 | -0.211 | 0.102 |
| | | Cumulative partners (last year) | F, 13-20 | Yes | 2060.6 | log(RR) | 18 | -0.151 | 0.126 |
| | | Partner age difference 5+ years | F, 13-20 | Yes | 2060.6 | log(RR) | 18 | -0.105 | 0.113 |
| | | Cumulative casual sex | F, 13-20 | Yes | 2060.6 | log(RR) | 18 | -0.051 | 0.099 |
| | Abdool Karim et al [163] | Cumulative HIV incidence | M+F, | Yes | 629.6 | log(RR) | 24 | 0.182 | 0.247 |
| | | Cumulative HSV-2 incidence | 15-19 | Yes | 629.6 | log(RR) | 24 | -0.357 | 0.105 |
| | Handa et al [72, 168] | Condom use at last sex | M+F, | NS | 2160.6 | log(RR) | 48 | 0.084 | 0.108 |
| | | Cumulative partners (last year) | 11-21 | NS | 2160.6 | log(RR) | 48 | -0.458 | 0.338 |
| | | Sexual debut | F, 11-21 | NS | 2160.6 | log(OR) | 48 | -0.552 | 0.209 |
| | | Sexual debut | M, 11-21 | NS | 2160.6 | log(OR) | 48 | -0.298 | 0.171 |
| | | Cumulative pregnancy incidence | F, 8-20 | NS | 2160.6 | log(RR) | 48 | -0.298 | 0.144 |
| | | Cumulative incidence of marriage | F, 8-20 | NS | 2160.6 | log(RR) | 48 | -0.038 | 0.087 |
| | | Cumulative incidence of marriage | M, 14-26 | NS | 2160.6 | log(RR) | 48 | -0.044 | 0.140 |

| | | | | | | | | | |
|---|---|---|---|---|---|---|---|---|---|
| | Baird et al [71]* | School dropout | F, 13-22 | Yes | 1081.8 | log(OR) | 12 | -0.199 | 0.238 |
| | | Cumulative pregnancy incidence | F, 13-22 | Yes | 1081.8 | log(OR) | 12 | -1.833 | -0.723 |
| | | Sexual debut | F, 13-22 | Yes | 1081.8 | log(OR) | 12 | -0.329 | 0.339 |
| | | HIV prevalence | F, 13-22 | Yes | 1081.8 | log(OR) | 18 | -0.755 | 0.620 |
| | | HSV-2 prevalence | F, 13-22 | Yes | 1081.8 | log(OR) | 18 | -2.526 | 1.036 |
| | De Walque et al [169] | Cumulative NG/CT/TV incidence | M, 18-30 | NS | 299.5 | log(RR) | 12 | 0.270 | 0.243 |
| | | Cumulative NG/CT/TV incidence | F, 18-30 | NS | 299.5 | log(RR) | 12 | -0.020 | 0.191 |
| | | Cumulative NG/CT/TV incidence | M, 18-30 | NS | 599.0 | log(RR) | 12 | -0.386 | 0.378 |
| | | Cumulative NG/CT/TV incidence | F, 18-30 | NS | 599.0 | log(RR) | 12 | -0.274 | 0.215 |
| | Schaefer et al [75] | Sexual debut | M, 15-20 | NS | 1025.3 | log(RR) | 12 | 0.276 | 0.398 |
| | | Sexual debut | F, 15-20 | NS | 1025.3 | log(RR) | 12 | 0.153 | 0.186 |
| | | Condom use at last sex | M, 15-29 | NS | 1025.3 | log(RR) | 12 | 0.070 | 0.185 |
| | | Condom use at last sex | F, 15-29 | NS | 1025.3 | log(RR) | 12 | -0.011 | 0.240 |
| | | Condom use at last sex | M, 30-54 | NS | 1025.3 | log(RR) | 12 | -0.101 | 0.245 |
| | | Condom use at last sex | F, 30-54 | NS | 1025.3 | log(RR) | 12 | 0.051 | 0.136 |
| | | Cumulative partners (last year) | M, 15-29 | NS | 1025.3 | log(RR) | 12 | 0.396 | 0.273 |
| | | Cumulative partners (last year) | M, 30-54 | NS | 1025.3 | log(RR) | 12 | -0.225 | 0.366 |
| | | Cumulative partners (last year) | F, 30-54 | NS | 1025.3 | log(RR) | 12 | -0.274 | 0.508 |
| | | School dropout | M, 15-20 | NS | 1025.3 | log(RR) | 12 | -0.394 | 0.181 |
| | | School dropout | F, 15-20 | NS | 1025.3 | log(RR) | 12 | 0.034 | 0.131 |
| | Kohler & Thornton [122] | Unprotected sex with recent partners | M, 18+ | NS | 36.1 | log(RR) | 1 | 0.161 | 0.078 |
| | | Unprotected sex with recent partners | F, 18+ | NS | 36.1 | log(RR) | 1 | -0.154 | 0.090 |
| | | Unprotected sex with recent partners | M, 18+ | NS | 144.3 | log(RR) | 1 | 0.168 | 0.092 |
| | | Unprotected sex with recent partners | F, 18+ | NS | 144.3 | log(RR) | 1 | -0.085 | 0.081 |
| | American Institutes for | Sexual debut | M+F | NS | 928.1 | log(RR) | 36 | 0.018 | 0.069 |
| | Research [170] | First pregnancy | F, 13-20 | NS | 928.1 | log(RR) | 36 | 0.095 | 0.098 |
| | | Condom use at first sex | M+F, | NS | 928.1 | log(RR) | 36 | 0.033 | 0.132 |
| | | First partner age difference >10 years | 13-20 | NS | 928.1 | log(RR) | 36 | 0.438 | 0.168 |
| | Abdoulayi et al [74] | Sexual debut | M, 13-19 | NS | 797.9 | log(RR) | 27 | 0.037 | 0.095 |
| | | Sexual debut | F, 13-19 | NS | 797.9 | log(RR) | 27 | -0.146 | 0.122 |
| | | First pregnancy | F, 13-19 | NS | 797.9 | log(RR) | 27 | 0.076 | 0.112 |
| | | Condom use at first sex | M+F, | NS | 797.9 | log(RR) | 27 | -0.123 | 0.109 |
| | | First partner age difference >5 years | 13-19 | NS | 797.9 | log(RR) | 27 | -1.211 | 0.947 |
| School | Cho et al [68] | School dropout | M+F, | Yes | 900.2¶ | log(OR) | 12 | -1.297 | 0.854 |
| support | | Sexual debut | 12-14 | Yes | 900.2 | log(RR) | 12 | -2.103 | 1.038 |
| | Cho et al [69] | School dropout | M+F, | Yes | 900.2¶ | log(OR) | 36 | -1.204 | 0.271 |
| | | Cumulative HIV incidence | 11-20 | Yes | 900.2 | log(OR) | 36 | -0.329 | 0.798 |
| | | Cumulative HSV-2 incidence | M+F, | Yes | 900.2 | log(OR) | 36 | -0.020 | 0.304 |
| | | Cumulative incidence of marriage | 11-20 | Yes | 900.2 | log(OR) | 36 | -0.386 | 0.537 |

| | | | | | | | | |
|---|---|---|---|---|---|---|---|---|
| | Cumulative pregnancy incidence | F, 11-20 | Yes | 900.2 | log(OR) | 36 | -0.431 | 0.312 |
| | Sexual debut | M+F | Yes | 900.2 | log(OR) | 36 | -0.186 | 0.213 |
| | Circumcision | M, 11-20 | Yes | 900.2 | log(OR) | 36 | 0.507 | 0.248 |
| Duflo et al [67, 171] | Sexual debut | M, 11-16 | Yes | 71.8¶ | log(RR) | 24 | 0.019 | 0.035 |
| | Sexual debut | F, 11-16 | Yes | 71.8 | log(RR) | 24 | -0.137 | 0.064 |
| | Condom use at last sex | M, 11-16 | Yes | 71.8 | log(RR) | 24 | 0.108 | 0.081 |
| | Condom use at last sex | F, 11-16 | Yes | 71.8 | log(RR) | 24 | 0.039 | 0.048 |
| | School dropout | M, 11-16 | Yes | 71.8 | log(RR) | 36 | -0.145 | 0.063 |
| | School dropout | F, 11-16 | Yes | 71.8 | log(RR) | 36 | -0.182 | 0.090 |
| | Cumulative pregnancy incidence | F, 11-16 | Yes | 71.8 | log(RR) | 36 | -0.098 | 0.062 |
| | HSV-2 prevalence | M, 11-16 | Yes | 71.8 | log(RR) | 84 | -0.027 | 0.093 |
| | HSV-2 prevalence | F, 11-16 | Yes | 71.8 | log(OR) | 84 | -0.061 | 0.086 |
| Hallfors et al [172] | School dropout | F, 10-16 | Yes | 804.6¶ | log(OR) | 24 | -2.138 | 0.435 |
| | Cumulative incidence of marriage | F, 10-16 | Yes | 804.6 | log(OR) | 24 | -1.072 | 0.540 |
| Gorgens et al [173, 174] | Cumulative HIV incidence | F, 15-22 | NS | 1075.4 | log(OR) | 36 | -0.261 | 0.128 |
| Baird et al [71]† | School dropout | F, 13-22 | Yes | 1081.8 | log(OR) | 12 | -0.732 | 0.308 |
| | Cumulative pregnancy incidence | F, 13-22 | Yes | 1081.8 | log(OR) | 12 | 0.157 | 0.374 |
| | Sexual debut | F, 13-22 | Yes | 1081.8 | log(OR) | 12 | -0.545 | 0.351 |
| | HIV prevalence | F, 13-22 | Yes | 1081.8 | log(OR) | 18 | -1.238 | 0.609 |
| | HSV-2 prevalence | F, 13-22 | Yes | 1081.8 | log(OR) | 18 | -0.994 | 0.528 |
| | Cumulative pregnancy incidence | F, 13-22 | No | 1081.8 | log(OR) | 12 | -0.598 | 0.365 |
| | Sexual debut | F, 13-22 | No | 1081.8 | log(OR) | 12 | -0.357 | 0.370 |
| | HIV prevalence | F, 13-22 | No | 1081.8 | log(OR) | 18 | 0.315 | 0.329 |
| | HSV-2 prevalence | F, 13-22 | No | 1081.8 | log(OR) | 18 | 0.030 | 0.397 |

* Unconditional cash transfer trial arm. † Conditional cash transfer trial arm. ¶ The cash-equivalent value is halved to represent the lower value attached to non-cash material support. CT = *Chlamydia trachomatis* (chlamydia). F = female. M = male. NG = *Neisseria gonorrhoeae* (gonorrhoea). NS = not specified. OR = odds ratio. RR = risk ratio. SE = standard error of intervention effect. TV = *Trichomonas vaginalis* (trichomoniasis). ZAR = South African rands.

##### 2.2.4 Likelihood definition

For each trial, and for each outcome measured in that trial, we calculate a likelihood that measures the extent of consistency between the outcome measured in the trial and the outcome predicted by the model. The outcome predicted by the model is calculated by simulating the outcome in the intervention arm (starting in 2005, we assign individuals to receive the intervention if they are eligible, and track their outcomes over the term of the trial) and comparing this to the simulated outcome in the control arm (restarting the model from 2005, in the same simulated individuals who would have qualified for the intervention, but now assuming that they do not receive the intervention). The time at which the intervention starts is arbitrary; most of the RCTs of structural interventions in Africa have been conducted at a time of declining HIV incidence, after the start of ART rollout, and we chose 2005 to approximately match this. For the sake of defining eligibility in our model, we consider only people whose log per capita household income is below the national average, as economic interventions are almost always targeted toward people of lower socio-economic status. The modelled eligible population is further restricted to match the age, sex and schooling profile of the population recruited into the corresponding RCT.

The likelihood is calculated on the assumption that the observed trial outcome is normally distributed, with mean equal to the model prediction and variance calculated from the standard error reported for the trial outcome. For example, consider the vocational training and microfinance intervention for adolescent girls evaluated by Dunbar *et al* [86] in Zimbabwe. In this RCT, the risk of unintended pregnancy in the intervention arm was 0.62 (95% CI: 0.38-1.02) times that in the control arm. In order to normalize the variance of the hazard ratio, we convert it to the log scale, so that the effect size is -0.48 (log(0.62)) and the standard deviation is 0.25 (which we approximate from the 95% confidence interval: (log(1.02) – log(0.38))/(2 × 1.96)). Suppose that for a given sampled parameter combination, the model estimates that the relative rate of pregnancy in intervention recipients is 0.8. If we ignore random effects, the likelihood for that parameter combination, in respect of the pregnancy outcome in the Dunbar trial, is calculated as the density of the $N(0, 0.25^2)$ distribution at the value -0.25 (-0.48 – log(0.8)), which is 0.97.

*Accounting for random effects*

In the above example, we were calculating the likelihood on the assumption that when matching the age and sex of the intervention participants, our South African model can reliably approximate the intervention effect we would expect to observe in Zimbabwe. In reality there are differences across settings in the salience of different structural drivers, and RCTs are likely to differ in their eligibility criteria and design, even when they test broadly similar interventions. In the meta-analyses that were used to set the prior distributions for the economic intervention impacts (section 1.6), estimates of the between-study variance (tau-squared statistics) were included: 0.02 in the case of the Tripney *et al* meta-analysis of vocational training intervention effects on paid employment [152], 0.15 in the case of the Garcia and Saavedra meta-analysis of the effects of cash transfers on school dropout [155], and 0.19 in the case of the Wilson *et al* meta-analysis of the effect of school support programmes on school dropout [154] (in all cases the effects were expressed on a log odds ratio or log risk ratio scale). The corresponding estimates of the standard deviation of between study differences (tau) are 0.14, 0.38 and 0.44 respectively. We take the average of these standard deviations (0.32) when determining the likelihood for outcomes defined on a log odds ratio or log risk ratio scale. This means increasing the standard deviation around the RCT estimate to take account of the ‘random error’ specific to the RCT. In the Dunbar example above, this means replacing the standard deviation of 0.25 with a standard deviation of 0.41 ($\sqrt{0.25^2 + 0.32^2}$).

*Accounting for stochastic variation*

A limitation of using a stochastic model such as MicroCOSM is that the model outputs, for a given input parameter combination, are never precise – they vary depending on the random numbers selected. In an attempt to quantify this stochastic variation, we run the model twice in each scenario (with different random numbers). Suppose that $Y_{ij}$ represents the observed value of the $i^{th}$ outcome in study $j$, after appropriate transformation (in the previous example from the Dunbar RCT, this would be -0.48). Further suppose that $X_{ijk}(1)$ and $X_{ijk}(2)$ represent the corresponding model estimates of the RCT impacts when using the first and second sets of random numbers respectively, and the $k^{th}$ set of input parameters. If we were to run the model with 5000 different input parameter combinations ($k$ = 1, 2, …, 5000), we would approximate the stochastic variance corresponding to the $i^{th}$ outcome in study $j$ as

$$\sigma_S^2(i,j) = \frac{0.5}{5000}\sum_{k=1}^{5000}\left(X_{ijk}(1) - X_{ijk}(2)\right)^2.$$

This is used to inflate the total variance around the difference between the average model prediction ($0.5 \times \left(X_{ijk}(1) + X_{ijk}(2)\right)$) and the observation ($Y_{ij}$). Thus the difference between the average model prediction and the observation is assumed to be normally distributed with mean zero and variance

$$\sigma_S^2(i,j) + \sigma_O^2(i,j) + \sigma_R^2,$$

where $\sigma_O^2(i,j)$ is the variance reported for the corresponding trial outcome ($0.25^2$ in the Dunbar example outlined previously), and $\sigma_R^2$ is the random effect variance ($0.32^2$, from the previous section). Suppose that in the Dunbar example, we estimated $\sigma_S^2(i,j)$ = 0.027; then the total variance would be 0.192 ($0.027 + 0.25^2 + 0.32^2$).

*Further refinements: cash transfers*

In the case of the cash transfer interventions and school support interventions that involve some form of financial assistance, we also scale the modelled impact of the intervention to the magnitude of the cash transfer (or the cash value of the school support if it is in the form of uniforms, school fees, etc.). For scaling purposes, the magnitude of the cash transfer is expressed in 2005 South African rand terms, converting from the US dollar equivalent value using the 2005 US dollar-rand exchange rate. In addition, because of purchasing power differences between countries we multiply by the ratio of the purchasing power of a dollar in the country in which the trial was conducted to the purchasing power of a dollar in South Africa in 2005. The ‘purchasing power of a dollar’ is in turn calculated as the ratio of the market exchange rate to the purchasing power parity (PPP) exchange rate (as reported by the World Bank [175]). In South Africa in 2005, for example, the market exchange rate was R6.80 to the dollar, while the PPP exchange rate was R3.56 to the dollar, so the purchasing power of a dollar is calculated as 1.91 (6.80/3.56). (This implies a dollar in 2005 would have bought 1.91 times as many goods and services in South Africa as it would have in the US.)

To give an example of how the adjustment is applied, the value of the cash transfer in the Kenya Cash Transfer for Orphans and Vulnerable Children (CT-OVC), started in 2007, was $240 per annum [72]. This would have been equivalent to R1632 (240 × 6.8) in 2005, ignoring dollar inflation. The purchasing power of a dollar in Kenya in 2007 was 2.53, so we inflate the cash transfer by a factor of 1.32 (2.53/1.91) to get a South African equivalent of R2161 in 2005. If the cash transfer that we simulate is R800 per annum in 2005 and the model simulates that this R800 cash transfer leads to a reduction of 0.15 in the log odds of sexual debut, then it is assumed that the corresponding reduction for a cash transfer that is R2161 is 0.25 ($0.15 \times (2161/800)^{0.5}$). This is compared against the actual reduction in the log odds of sexual debut in the Kenya CT-OVC trial, 0.55, for the purpose of calculating the likelihood. Note that we use a square root scaling rather than a linear scaling, to be consistent with our model assumption that income effects on sexual risk behaviour are generally non-linear. The choice of R800 per annum as the assumed value of the cash transfer in the base simulation is based on the median value of the cash transfers in the included studies (Table S28). The amount is assumed to increase in line with the South African consumer price index for each year after 2005.

*Further refinements: school support*

Similar scaling adjustments are made in the case of school support interventions, which typically include either a conditional cash transfer (conditional upon school attendance) or some other form of material support (e.g. providing free school uniforms or covering school fees). In the latter case, we consider the cash equivalent value of the support. The cash value of the school support is then added to the household income, in the same way as for 'pure' cash transfers. In addition to the effect of household income, we model an effect of school support on the probability of school dropout, as described in section 1.6. For example, in the RCT of Baird *et al* [71] in Malawi, the odds of retention in school, among girls who were in school at baseline, was significantly greater in the arm that was randomized to conditional cash transfers than in the control arm (OR 2.08, 95% CI: 1.14-3.82). This is equivalent to a 0.73 reduction in the log odds of dropout (-log(2.08)), with a standard deviation of 0.31. The intervention started in 2007 and the annual value of the conditional cash transfer was $120. Using the same approach as before, this would have been equivalent to R816 (120 × 6.8) in 2005, ignoring dollar inflation. The purchasing power of a dollar in Malawi in 2007 was 2.53 times that in the US (coincidentally the same as in Kenya), so we inflate the cash transfer by the same factor of 1.32 (2.53/1.91) to get a South African equivalent of R1082 in 2005. If the cash transfer that we simulate is R800 per annum in 2005 and the model simulates that this R800 cash transfer leads to a reduction of 0.15 in the log odds of school dropout, then it is assumed that the corresponding reduction for a cash transfer that is R1082 is 0.17 ($0.15 \times (1082/800)^{0.5}$). (For the purpose of this illustration, we are considering only the effect of the cash support, and ignoring the non-material support, as represented by the *S* factor in section 1.6.)

In the previous example, the Malawian school support programme involved a cash transfer. However, in most of the other school support RCTs, the material support was in the form of school uniforms, textbooks, payment of school fees, etc. One could argue that households might not value these to the same extent as the cash equivalent value of the support. For example, rather than buying new uniforms and textbooks, the household might prefer to use the money to buy food and send their child to school in an old uniform, with instructions to borrow textbooks from friends. One can only add the full cash equivalent value of the material support to the household income if the household would have purchased new uniforms and textbooks in the absence of the intervention. Because we do not know to what extent this is true, we add only half of the cash equivalent value of the material support to the household income (and similarly consider only half the value of the material support when adjusting the rate of school dropout). Although this decision to use a multiplier of 0.5 is arbitrary, we show in section 3.3 that the modelled estimate of the intervention impact on school dropout is in fact relatively insensitive to the choice of multiplier.

*Limitations*

A limitation of the model of random effects is that we do not consider the possibility that the variation in random effects might be greater for more 'down-stream' outcomes (e.g. HIV incidence) than for 'up-stream outcomes' (e.g. education), or that the extent of the random effects may be different across intervention types. Another limitation is that we have somewhat arbitrarily set the intervention start year in the model to 2005, and have not tried to match this to the actual start years in the trial. However, we do not anticipate that there would be major changes in intervention effectiveness over time, and using the same start year for all model simulations has the advantage of reducing the number of model runs that are required.

#### 2.3 Posterior sampling

We generate our posterior sample using Sampling Importance Resampling [176]. This means drawing a resample of 50 parameter combinations from the initial set of 5000 parameter combinations, using the likelihood values as weights. More detailed results are generated using these 50 parameter combinations, and the posterior distribution is approximated by these 50 simulations.

We consider four sets of posterior samples. Firstly, we consider the posterior distribution obtained when all of the data in Table S28 are used to define the likelihood (unless stated otherwise, these are the results that define the posterior distributions presented). Then we consider the posterior estimates obtained when each of the three intervention types is considered separately; these results are used only when presenting the projected impact of each intervention in isolation of the other socio-economic interventions.

#### 2.4 Posterior estimates of model parameters

Table S29 compares the prior and posterior distributions for the parameters that were varied in the model calibration. Most of the posterior distributions are not significantly different from the priors. However, when calibrating to the vocational training RCT data, we estimate a less dramatic impact of the intervention on unemployment than the prior distribution suggests. When calibrating to cash transfer RCT data, we find the posterior estimate of the effect of household income on female entry into casual sex is greater than estimated based on previously published literature. When calibrating to the school support RCT data, we estimate a greater effect of being in school on rates of female sexual debut (relative to the effects implied by the prior distributions). In addition, the school support RCT data suggest that the income component to the school support has more effect than assumed *a priori*.

Table S29: Comparison of prior and posterior distributions (means, with 95% confidence intervals in brackets)

| Parameter | Prior distribution | Posterior distribution* | | | |
|---|---|---|---|---|---|
| | | Main | Vocational training | Cash transfers | School support |
| Increase in rate of debut in females per log reduction in APCHI† (RR – 1) | 0.23 (0.00-0.61) | 0.24 (0.07-0.59) | 0.22 (0.00-0.60) | 0.24 (0.00-0.63) | 0.24 (0.00-0.70) |
| RR of debut in females if currently in school | 0.46 (0.04-1.00) | **0.25 (0.00-0.99)** | 0.41 (0.09-1.00) | 0.49 (0.06-1.00) | **0.37 (0.02-1.00)** |
| RR of debut in males if currently in school | 0.80 (0.19-1.00) | 0.71 (0.10-1.00) | 0.81 (0.31-1.00) | 0.80 (0.28-1.00) | 0.86 (0.31-1.00) |
| Increase in casual sex in females, per log reduction in APCHI† (RR – 1) | 0.43 (0.00-1.60) | 0.34 (0.00-1.46) | 0.48 (0.00-1.29) | **0.62 (0.00-1.60)** | 0.36 (0.00-1.26) |
| Increase in casual sex in men who are employed (RR – 1) | 0.25 (0.00-0.97) | 0.28 (0.00-1.50) | 0.24 (0.00-1.02) | 0.26 (0.00-0.88) | 0.29 (0.00-0.85) |
| Increase in commercial sex in men who are employed (RR – 1) | 0.40 (0.00-1.55) | 0.51 (0.00-1.38) | 0.38 (0.00-1.22) | 0.42 (0.00-1.37) | 0.36 (0.00-1.26) |
| RR of partner acquisition in men if employed (RR – 1) | 0.57 (0.00-2.34) | 0.52 (0.00-1.71) | 0.51 (0.00-1.89) | 0.55 (0.00-2.40) | 0.54 (0.00-2.12) |
| Increase in consistent condom use per year of schooling (OR – 1) | 0.05 (0.00-0.19) | 0.04 (0.00-0.16) | 0.04 (0.00-0.28) | 0.05 (0.00-0.20) | 0.05 (0.00-0.16) |
| RR of marriage if currently in school | 0.37 (0.02-1.00) | **0.46 (0.09-1.00)** | 0.32 (0.03-1.00) | 0.32 (0.05-1.00) | 0.40 (0.06-1.00) |
| Increase in medical male circumcision per log increase in APCHI (RR – 1) | 0.15 (0.00-0.42) | 0.16 (0.00-0.36) | 0.15 (0.00-0.44) | 0.19 (0.00-0.48) | 0.12 (0.00-0.37) |
| RR of school dropout if receiving school support (non-material) | 0.68 (0.29-0.96) | 0.70 (0.53-0.93) | 0.70 (0.34-0.96) | 0.70 (0.36-0.98) | 0.72 (0.49-0.92) |
| RR of dropout per R800 of school support | 0.86 (0.61-0.99) | **0.78 (0.56-0.96)** | 0.88 (0.68-0.99) | 0.86 (0.70-0.99) | **0.80 (0.51-0.98)** |
| OR of unemployment if receiving vocational training/microfinance | 0.87 (0.62-0.99) | **0.94 (0.81-0.99)** | **0.92 (0.81-1.00)** | 0.88 (0.66-1.00) | 0.87 (0.64-0.99) |

APCHI = adjusted per capita household income. OR = odds ratio. RR = relative risk
* The four posterior distributions correspond to four different ways of defining the likelihood (see section 2.3). Values in bold represent significant differences between the prior and posterior distributions.

#### 2.5 Calibration outputs

Figure S13 compares the modelled effects of the economic empowerment interventions against those observed in the RCTs, focusing only on the socio-economic outcomes. The observed effects of vocational training on employment and income are heterogeneous, and the modelled effects are generally of similar magnitude. RCT estimates of the effect of school support on school dropout are also very heterogeneous, while model estimates are more stable – greater than the modest effects observed by Duflo *et al* [67] but less than the effects measured by Hallfors *et al* [172]. Our model suggests that pure cash transfer interventions have relatively little effect on school dropout, consistent with most RCTs.

Figure S14 compares the modelled and observed effects of interventions on behavioural outcomes. The model estimates that these interventions generally have little impact on sexual behaviour outcomes. Although this is consistent with most RCT data, modelled estimates of the effects of school support interventions on sexual debut and marriage are generally smaller than those measured in RCTs.

Figure S15 compares the modelled and observed effects of economic strengthening interventions on biomedical outcomes. The model predicts modest reductions in HIV incidence and prevalence for most of the simulated interventions. However, the model predicts almost no change in the STIs and teenage pregnancy incidence. These predictions are mostly consistent with RCT data, with the notable exception of the trial of Baird *et al* [71], which found much more substantial reductions in the prevalence of HSV-2 and HIV than our model.

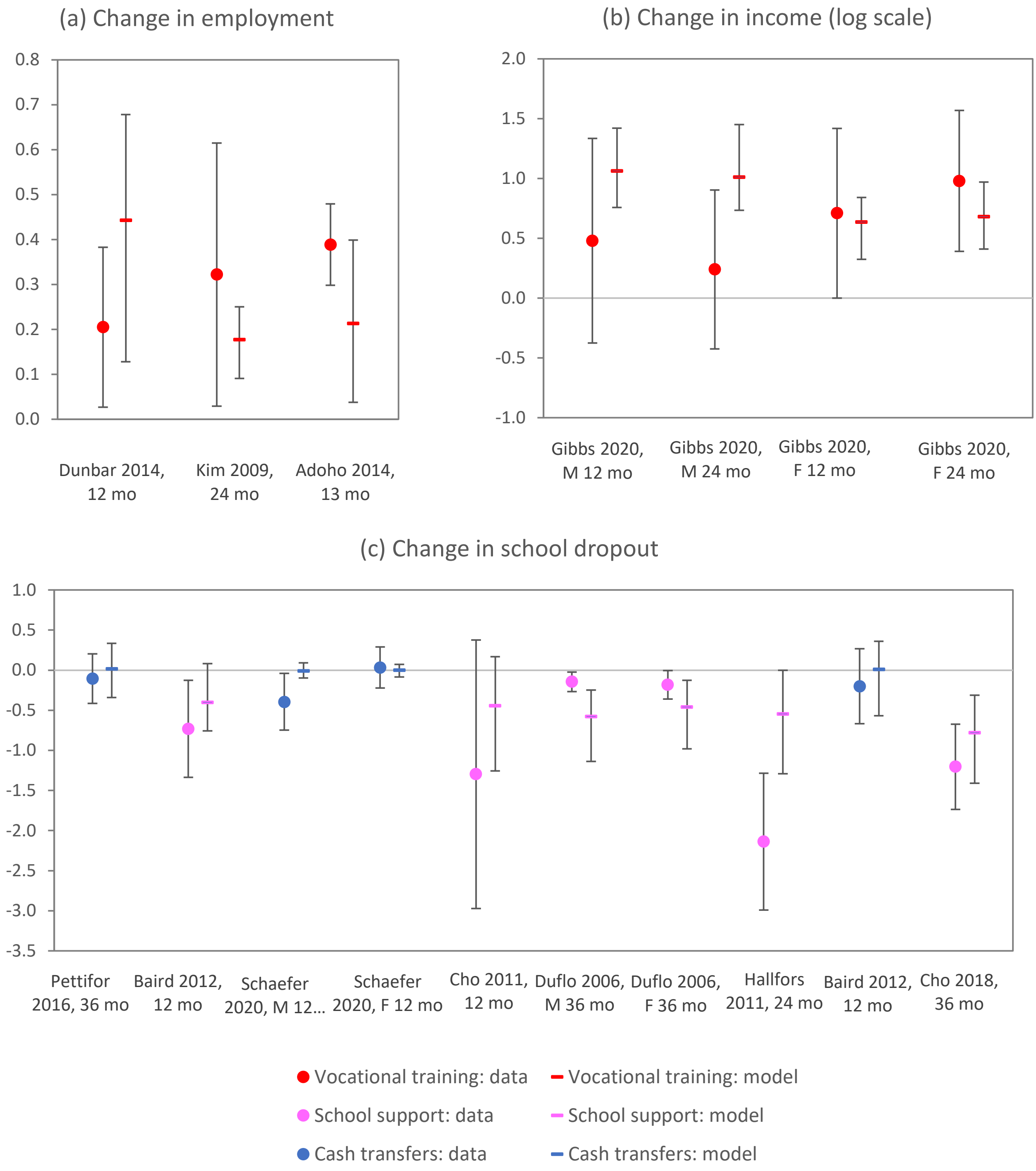

Figure S13: Calibration to socio-economic outcome data

In all panels the observed trial outcomes (dots) are compared against the model predictions of the trial impact (horizontal dashes), averaging the 50 posterior model results. Vertical lines represent 95% confidence intervals. In panels (a) and (c), the measure of intervention impact is the logarithm of the odds ratio or relative risk (comparing the intervention arm to the control arm), and in panel (b) the measure of intervention impact is the change in income on a log scale.

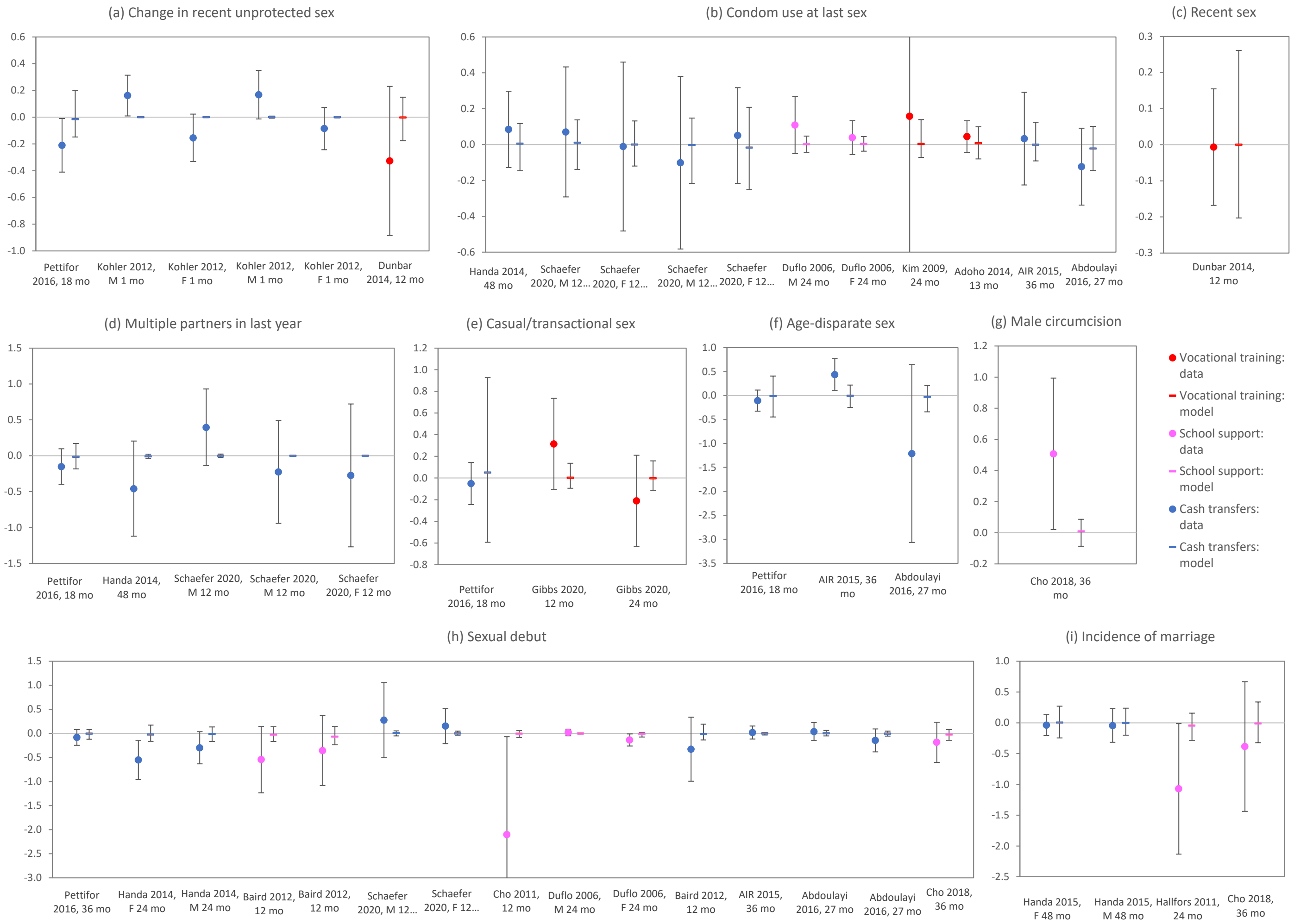

#### Figure S14: Calibration to sexual risk behaviour outcome data

In all panels the observed trial outcomes (dots) are compared against the model predictions of the trial impact (horizontal dashes), averaging the 50 posterior model results. Vertical lines represent 95% confidence intervals. In all panels, the measure of intervention impact is the logarithm of the odds ratio or relative risk when comparing the intervention arm to the control arm.

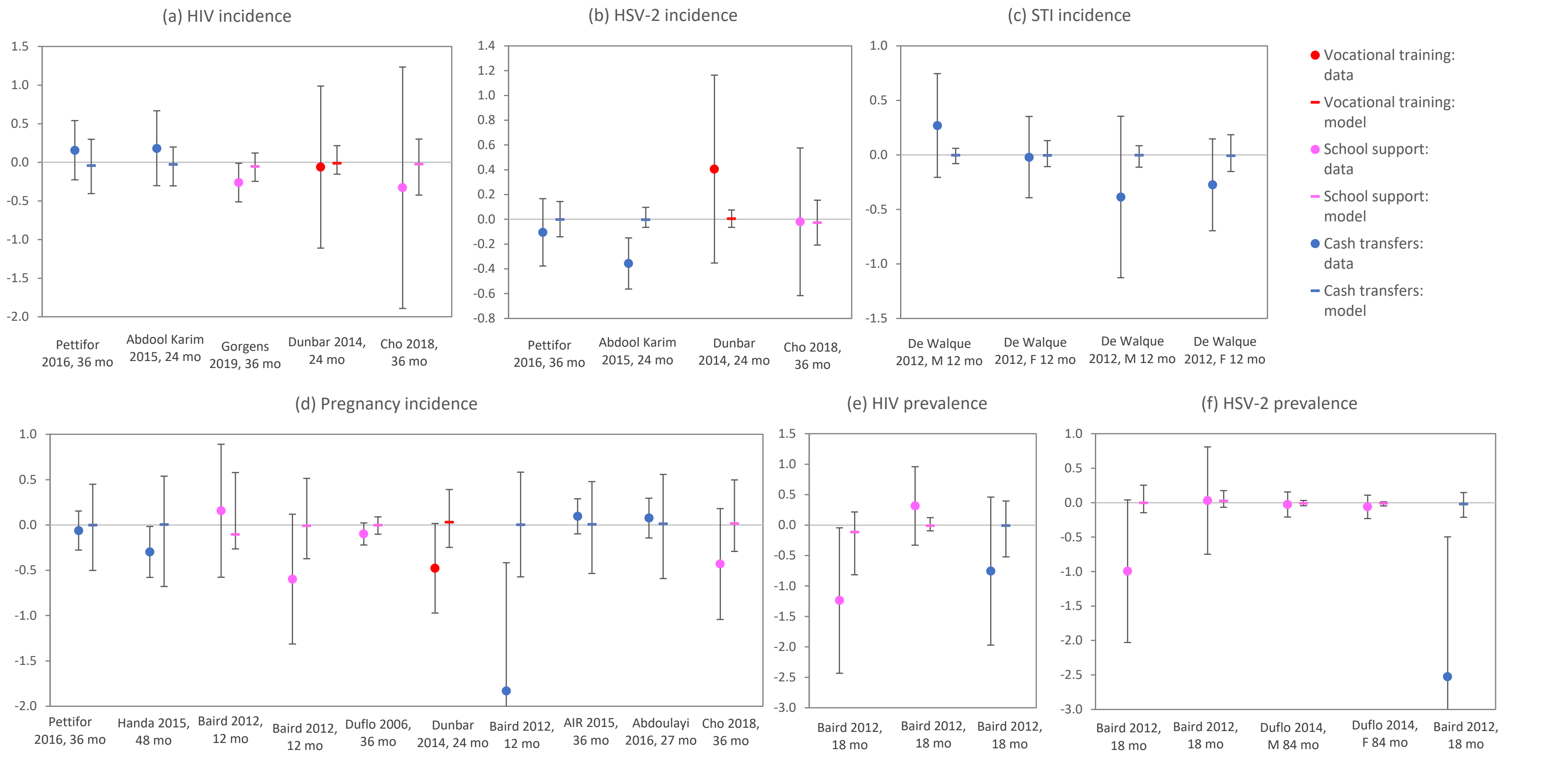

### Figure S15: Calibration to biological outcome data

In all panels the observed trial outcomes (dots) are compared against the model predictions of the trial impact (horizontal dashes), averaging the 50 best-fitting model results. Vertical lines represent 95% confidence intervals. In all panels, the measure of intervention impact is the logarithm of the odds ratio or relative risk when comparing the intervention arm to the control arm. HSV-2 = herpes simplex virus type 2 (genital herpes), STI = sexually transmitted infections (here limited to gonorrhoea, chlamydia and trichomoniasis).

### 3. Additional results

#### 3.1 Validation against household survey data

We compared the model estimates of the association between socio-economic measures, sexual risk behaviour and HIV, with the associations measured in four national household surveys. The four surveys are the Human Science Research Council (HSRC) surveys of 2008 [177], 2012 [178] and 2017 [179], and the 2016 Demographic and Health Survey [79]. (Information on HIV risk behaviours was lacking for the 2005 HSRC survey, and this survey has therefore not been included; individual-level data from the more recent 2022 survey have not yet been published.) Because socio-economic indicators vary substantially between race groups in South Africa, and we wanted to avoid potential confounding due to possibly mis-specified race effects, we have limited this comparison to black South Africans (who account for approximately 80% of the South African population). We have also limited the comparison to sexually active adults (ages 15-49). All comparisons are shown separately for men and women (although in the case of paying for sex, we only show results for men, and in the case of early marriage we only show the results for young women). The measure of association used in these comparisons is the log of the odds ratio, and for each of the surveys the log odds ratio is calculated from the individual-level survey data.

Figure S16 shows the results of the model validations. For the most part, the model results are consistent with the associations measured in the household surveys, although the confidence intervals around the model estimates are quite wide, reflecting the substantial uncertainty that remains in the relationship between socio-economic status and sexual risk behaviour, even after the model has been calibrated to the RCT data. The model does not simulate as strong an association between education and HIV in women as the survey data suggest (panel i), yet the model simulates a stronger association between education and condom use in women than is observed in the surveys (panel g) as well as a stronger reduction in multiple partnerships due to employment in women (panel c). The model also tends to suggest a positive association between employment and HIV-positive status in women (panel e), which is largely due to age confounding (i.e. unemployment is higher in young women than in older women, while older women have higher HIV prevalence). However, this is at odds with the survey data, which suggest no significant association between employment and HIV in women.

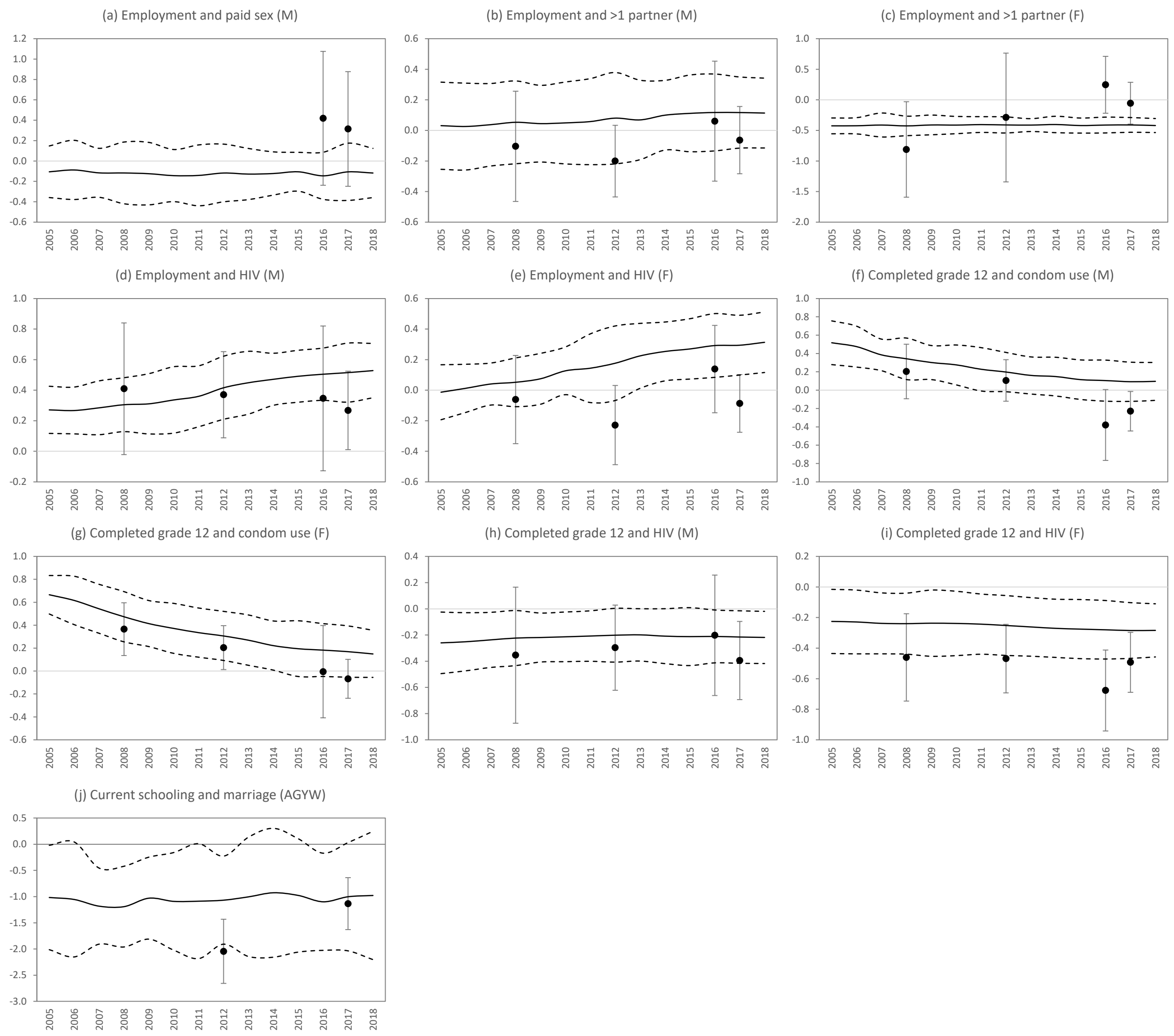

Figure S16: Modelled associations (on log(OR) scale) between socio-economic status, sexual risk behaviours and HIV, compared against associations measured in national surveys

Dots represent data for the black South African population (aged 15-49, except in panel j), and error bars reflect 95% confidence intervals. Solid lines represent the average of the modelled associations (across the 50 posterior parameter combinations) and dashed lines reflect the 95% confidence intervals around the modelled associations. AGYW = adolescent girls and young women (ages 15-24), F = females, M = males.

#### 3.2 Calibration to HIV and STI data

A number of changes were made to the model described previously [1, 30], in order to maintain consistency with the data that we have previously used in model calibration, as well as new calibration data:

- In previous versions of MicroCOSM, the prevalence of syphilis would often run to zero due to stochastic variation, with the result that syphilis would appear to be eradicated. This is unrealistic because in reality people in the simulated sub-population have sexual contact with people in other populations, and this can lead to STIs being re-introduced into the sub-population, even if they are temporarily eliminated. To prevent permanent elimination, we changed the model, assuming that at the start of each year one randomly chosen sex worker acquires syphilis from outside of the simulated population (and similarly for gonorrhoea, chlamydia, trichomoniasis and genital herpes). Figure S17 shows that the model estimates of STI prevalence in 2017 are roughly consistent with estimates from the Spectrum STI model for the same year [180], although the Spectrum STI model suggests a greater sex differential in chlamydia prevalence.

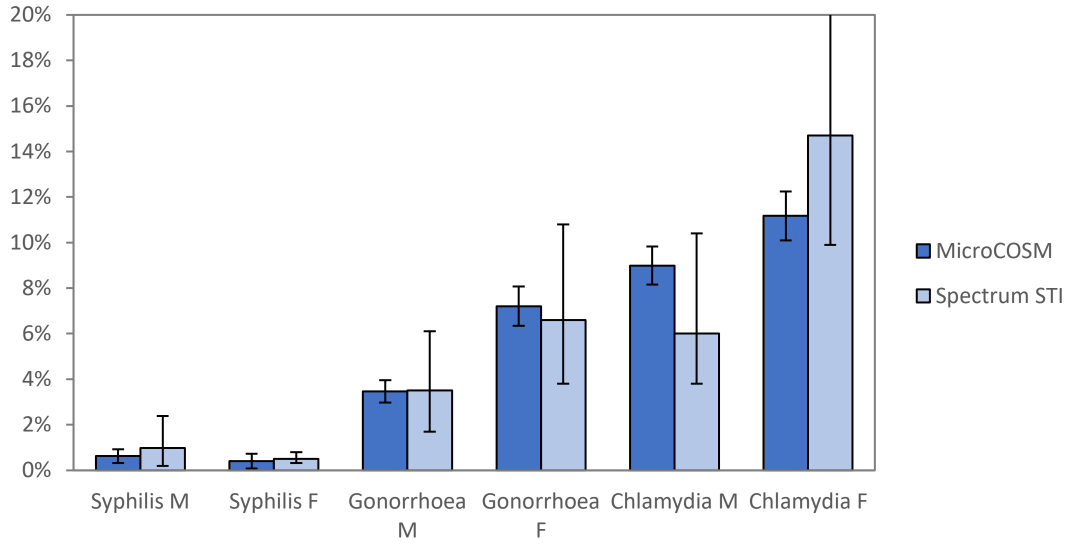

Figure S17: STI prevalence in 15-49 year olds in 2017

- Average heterosexual HIV transmission probabilities per act of unprotected sex have been reduced from the previously assumed values [30], as shown in Table S30. These changes were made in order to achieve approximate consistency with survey estimates of HIV prevalence in males, females and sex workers (Figure S18, panels a-c). No changes were made to the assumed male-to-male HIV transmission probabilities.

Table S30: Transmission probabilities per act of unprotected sex (untreated HIV)

| | Male-to-female | | | Female-to-male | | |
|---|---|---|---|---|---|---|
| | Client-FSW | Short-term | Long-term | FSW-client | Short-term | Long-term |
| Previous | 0.001020 | 0.001700 | 0.001327 | 0.000750 | 0.000750 | 0.000546 |
| New | 0.000612 | 0.001541 | 0.001128 | 0.000589 | 0.000589 | 0.000410 |

- To make the model of HIV in sex workers consistent with Thembisa version 4.8 [101], we adapted the model to assume that (a) HIV-positive women who know their HIV status are 37% less likely to enter commercial sex than HIV-positive women with the same risk characteristics who are undiagnosed; and (b) that men reduce their frequency of sex worker contact as their CD4 count declines. These represent the effects of HIV morbidity and fear of transmission to others, and these changes to the model prevent HIV prevalence in sex workers rising to implausibly high levels in the later stages of the HIV epidemic (Figure S18, panel c). The changes were previously described in section 1.4.3.
- Rates of HIV testing in the period after 2016 were updated to be consistent with recent programme data (see panel e of Figure S18).
- Adult ART initiation rates following diagnosis were updated to be consistent with those in Thembisa version 4.8 [101]. Model assumptions about ART interruption rates were also updated to be consistent with Thembisa, with ART interruption rates varying by age and sex (highest at age 20 and in men). These changes in ART assumptions led to

estimates of ART coverage that were roughly consistent with survey estimates (Figure S15, panel f).

- The relative infectivity of people on ART (the $I_5$ parameter in section 7.2 of the earlier MicroCOSM report [1]) was reduced from 0.2 to 0.08. This implies a 92% reduction in infectiousness after ART initiation (rather than the previously assumed 80% reduction). This is consistent with a systematic review of the effect of ART on HIV transmission probabilities [181]; the change was also made in order to ensure a more substantial decline in HIV incidence after the scale-up of ART (Figure S18, panel d).

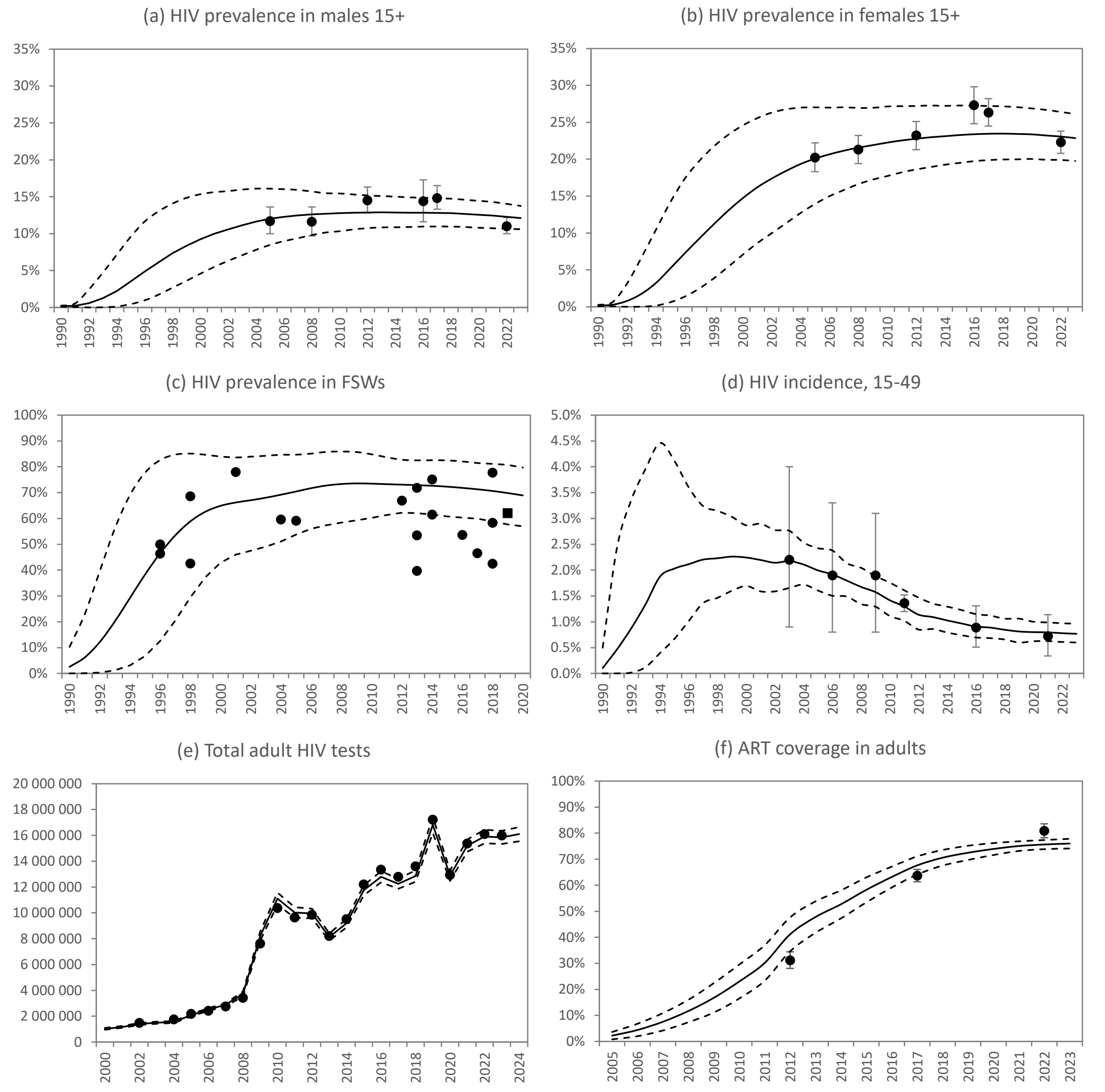

Figure S18: Model calibration to HIV data sources

In all panels, dots represent calibration targets, solid lines represent the posterior model mean and the dashed lines represent the 95% confidence interval around the model outputs (approximately corresponding to the 2.5 and 97.5 percentiles of the results from the 50 sets of results in the posterior sample).

#### 3.3 Sensitivity analysis: the value of non-cash transfers in school support

As noted in section 2.2.4, we apply a multiplier of 0.5 to the value of non-cash material support (e.g. school uniforms, textbooks) when adding this to the household income and when adjusting the school dropout rate. Because the multiplier of 0.5 is arbitrary, and there are no good data to inform this parameter, we consider here the sensitivity of the model results to alternative multiplier values. Specifically, we consider a simplified deterministic model of the rate of school dropout. Similar to the description in section 1.6, we model the relative rate of school dropout in youth who are receiving school support (with a non-cash transfer of value *x*) as

$$SC^{\sqrt{Ax/800}},$$

where *S* is the relative rate of school dropout due to the non-material component of the intervention (e.g. better attendance monitoring), *C* is the multiple by which the probability of school dropout is reduced for a R800 increase in the cash transfer value, and *A* is the multiplier described previously. We assign the same priors to the *S* and *C* parameters as described in section 1.6. A likelihood function is calculated, based on the six estimates of the effect of school support interventions on school dropout (Table S28). The likelihood function is calculated in the same was as described in section 2.2 (except that we omit the stochastic error variance term, since this simplified model is deterministic). We consider 10 possible values of *A* (0.1, 0.2, 0.3, …, 1.0). For each value of *A*, we (1) randomly sample 1000 parameters combinations from the prior distributions for *S* and *C*, (2) calculate the likelihood for each parameter combination, (3) resample 1000 parameter combinations, with replacement, from the original sample, using the likelihood values as sample weights, and (4) use these resamples as approximations to the posterior distribution [176].

Figure S19 summarizes the posterior distributions obtained for the different values of *A*. In general, the posterior means are not very sensitive to the choice of *A*: the posterior mean of *S* increases slightly as *A* increases, while the posterior mean of *C* decreases slightly as *A* increases. When considering the product of *S* and *C* (as a crude approximation to the combined effect of material and non-material support, for an ‘average’ intervention), there is virtually no change in the posterior mean across different values of *A*.

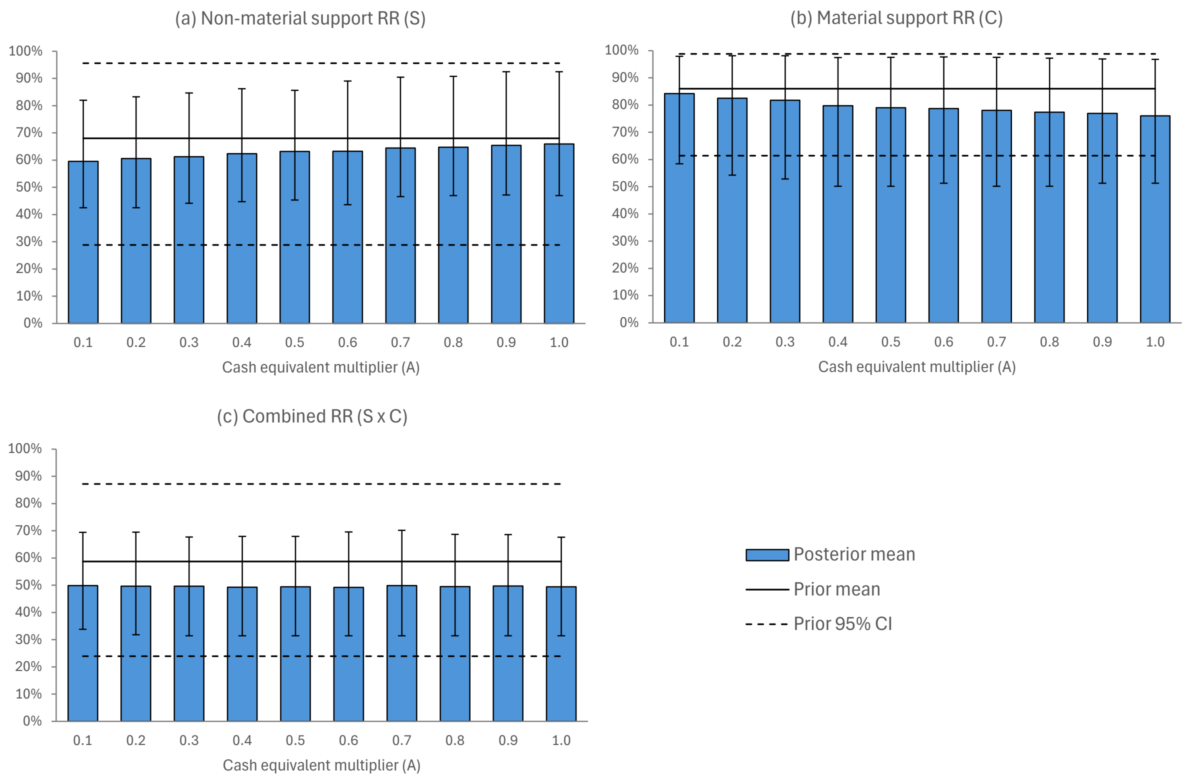

Figure S19: Effect of multiplier on estimates of school support effectiveness

It is worth noting that these posterior estimates (for $A = 0.5$) differ slightly from those in the main analysis (Table S29), which is primarily because the full likelihood is calculated for several outcomes (not only school dropout). Although school dropout is the outcome that most significantly affects the posterior likelihood, other outcomes also influence the likelihood, to the extent that the effect of school support on those outcomes is mediated by its effect on school dropout.
